# Nowcasting epidemic trends using hospital- and community-based virologic test data

**DOI:** 10.1101/2024.11.01.24316580

**Authors:** Tse Yang Lim, Sanjat Kanjilal, Shira Doron, Jessica Penney, Meredith Haddix, Tae Hee Koo, Phoebe Danza, Rebecca Fisher, Yonatan H. Grad, James A. Hay

## Abstract

Population viral loads measured by RT-qPCR cycle threshold (Ct) values are an alternative to case counts and hospitalizations for tracking epidemic trends, but their strengths, limitations and statistical power under various real-world conditions have not been explored. Here, we used SARS-CoV-2 RT-qPCR results from hospital testing in Massachusetts, USA, municipal testing in California, USA, and a combination of theory and simulation analysis to quantify biological and logistical factors impacting Ct-based epidemic nowcasting accuracy. We found that changes to peak viral load, viral growth and clearance rates, and sampling approach and delays all affect the relationship between growth rates and Ct values. We fitted generalized additive models to predict the growth rate and direction of SARS-CoV-2 incidence using time-varying Ct value distributions and assessed nowcasting accuracy over two-week windows. The model predicted epidemic growth rates and direction well from ideal synthetic data (growth rate RMSE of 0.0192; epidemic direction AUC of 0.926) but showed modest accuracy with real-world data (RMSE of 0.039-0.052; AUC of 0.72-0.78). Predictions were robust to testing regimes and sample sizes, and trimming outliers improved performance. Our results elucidate the possibilities and limitations of Ct value-based epidemic surveillance, highlighting where they may complement traditional incidence metrics.

## Introduction

Epidemic monitoring and outbreak surveillance are vital public health functions, providing early warning of emerging threats, informing healthcare capacity planning and transmission control policies, and helping to evaluate the effectiveness of interventions^1–4^. A common approach to epidemic monitoring, exemplified during the COVID-19 pandemic, is to track the incidence of reported positive diagnostic tests, clinical cases^5,6^, or deaths^7^. These data can inform key statistics such as the epidemic growth rate or effective reproductive number ^8–11^, and they are fundamental to nowcasting and forecasting an epidemic’s trajectory^12–14^. However, these data streams can be substantially lagged, biased, and incomplete due to testing delays, capacity limitations, cost, and changing test-seeking behavior^15,16^. Thus, there has been growing interest in alternative data sources, such as wastewater surveillance^17,18^, internet search trends^19^, and digital contact tracing^20^, that do not depend on large-scale testing of individuals.

One novel data source for epidemic monitoring described during the COVID-19 pandemic is the population-level distribution of viral loads among infected individuals, approximated using cycle threshold (Ct) values from reverse-transcription quantitative polymerase chain reaction (RT-qPCR) testing^21–24^. For certain acute respiratory viruses such as SARS-CoV-2, a low Ct value (high viral load) typically suggests that an individual was sampled early in their infection, whereas a high Ct value (low viral load) measurement suggests sampling later in infection^25–27^. A population-level sample of predominantly low Ct values (high viral loads) indicates that most sampled infections are recent onset, corresponding to a growing epidemic, whereas a sample of predominantly high Ct values (low viral loads) corresponds to a declining epidemic consisting of mostly late infections and post-infectious viral persistence^21^. Whereas count-based surveillance assumes that the number of reported positive tests tracks the true incidence of new infections, estimating epidemic growth rates using viral loads uses information in the distribution of Ct values from a representative sample of infected individuals.

Multiple studies have reported on the feasibility of using population-level Ct values to track SARS-CoV-2 epidemic trends, the majority demonstrating a negative correlation between average Ct values and either the effective reproduction number (*Rt*), case counts, or hospitalizations^28–38^ . Some have gone further to predict *Rt* or epidemic growth rates from Ct value statistics using linear regression^28,30,35,36,39^, autoregressive integrated moving average (ARIMA) models^33,37^, or machine-learning algorithms^40^. Other studies used the *virosolver* R package, which models the convolution of viral kinetics and incidence to predict population-level viral load distributions, to estimate infection incidence using Ct values from cross-sectional^21,31,32,35,36,40^. However, to our knowledge only one study has explored the use of Ct-based statistics for nowcasting epidemics (i.e., regularly updating growth rate estimates on a rolling basis) as opposed to retrospectively predicting entire epidemic waves^43^. In addition, these studies are generally limited to single settings and datasets, short time periods, and do not consider factors that may influence the accuracy of Ct-based inferences of epidemic growth rates, such as changes in viral kinetics and sampling schemes (e.g., symptom-based vs. random).

Here, we present an in-depth investigation of the real-world feasibility of using SARS-CoV-2 Ct values to nowcast epidemic trajectories, using a combination of theory, synthetic data, and multiple real-world datasets representing different sampling strategies and settings. We start with a theoretical exploration of how biological and logistical factors affect the non-linear relationship between population-level Ct value distributions and epidemic growth rates, using analytical convolutions of simple epidemic curves with viral load trajectories. We next establish how this relationship can be modeled to nowcast growth rates using reported Ct values. We then benchmarked nowcasting model performance using synthetic datasets, before applying the same models to three real SARS-CoV-2 RT-qPCR testing datasets, collected across multiple geographic areas in the United States and under different population sampling strategies, to assess and inform the use of this approach in real-time estimation of epidemic growth compared to using positive test counts alone. Our analyses demonstrate the possibilities and limitations of Ct-based epidemic growth rate surveillance.

## Results

### Theoretical relationship between epidemic growth rates and Ct value statistics

To understand how biological and practical factors might affect Ct-based nowcasting performance, we first examined their impact on the theoretical relationship between Ct value statistics and epidemic growth rates. We implemented analytical convolutions of population-level infection incidence from a Susceptible-Infected-Recovered model assuming *R*_0_=1.5 with various within-host viral kinetics models to generate mean Ct values observed through random or symptom-driven testing over an epidemic (**Figure S1; Supplementary Text 1**).

Figure 1 demonstrates the impact of viral kinetics properties (peak viral load, symmetry of viral kinetics, individual-level variance) and sampling approaches (distribution of delays between infection and sampling) on the predictability of growth rates using mean Ct values. These results demonstrate several key ideas. First, in most scenarios, there is a monotonic relationship between mean Ct value and growth rate, providing a basis for predicting the latter from the former. Second, the precision of growth rates estimated from Ct values depends on the shape and gradient of this relationship. Because there is uncertainty in the true population mean Ct value from a finite number of observations, a steeper gradient will correspond to a wider range of compatible growth rates for a given set of measured Ct values. Third, biological and logistical factors influence the shape of the relationship and hence prediction accuracy, but in different ways. For example, moving from a highly skewed viral kinetics curve to a less skewed one decreases the precision of the estimated growth rate, whereas moving to a fully symmetric curve results in a non-monotonic relationship and thus bi-modal estimates. Overall, this analysis demonstrates that although a relationship between mean Ct value and epidemic growth rate is always present, the strength of this relationship, and thus the statistical power of a Ct-based inference approach, varies depending on the underlying viral kinetics and sampling approach.

**Figure 1.**
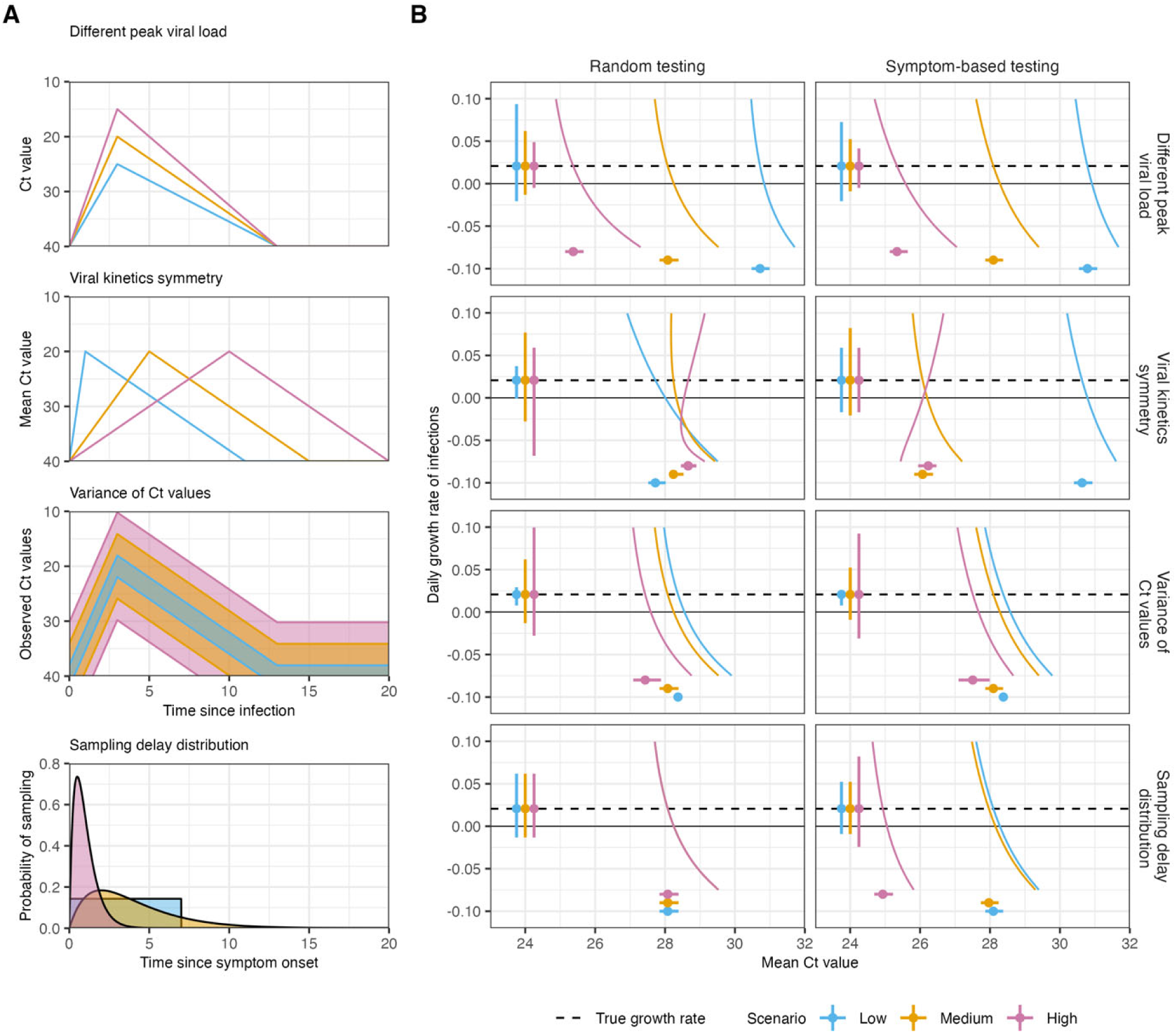
The analytical relationship between population mean Ct value and epidemic growth rate under a range of model assumptions and the impact on growth rate predictability. (**A**) Varying viral kinetics and sampling delay distribution assumptions. Top two rows show assumed mean Ct value over time-since-infection. Third row shows multiple assumed 95% quantiles of observed Ct values. Bottom row shows varying assumed distributions of delays between symptom onset and testing. Scenarios are colored to match (B). (**B**) Curved colored lines show the analytical relationship between mean Ct value and daily growth rate of new infections during a single wave epidemic, observed through random testing (first column) or following symptom onset (second column). The horizontal, colored lines show the 95% confidence intervals for the population mean Ct value, assuming the true mean corresponds to a growth rate of 0.02 (horizontal dashed line), a standard deviation of 3 (varied between 1, 3 and 5 in the bottom row), and sample size of 500. The vertical, colored lines show the range of epidemic growth rate values corresponding to the 95% CI of the mean Ct value.

### Modeling and nowcasting epidemic growth rates

We next examined how the Ct value-growth rate relationship can be used for epidemic nowcasting, applying the same set of basic models across several synthetic and real-world datasets. We used generalized additive models (GAM) with smoothing splines to predict growth rates of cases as a non-linear function of daily mean and skewness of observed Ct values (see **Supplementary Text 2**). We also fit GAMs to predict the epidemic direction (i.e., whether incidence is growing or declining) using a logit link function and with the same explanatory variables and smoothing splines.

With each dataset, we assessed in-sample fits of model-predicted vs. observed growth rates and directions across the entire dataset, based on root mean squared error (RMSE) for predicted growth rate and the area under the receiver operating characteristic curve (AUC) for predicted epidemic direction. For all datasets, model residuals were normally distributed around zero (example shown in **Figure S2**). We then refit the models using separate training and testing subsets of the data. To approximate a realistic application of the Ct value-based approach in an ongoing epidemic, we fit the models using only the first 16 weeks of data and then performed rolling nowcasts with a two-week time horizon, using the fitted model to estimate the epidemic growth rate and direction daily over the next two weeks based on the Ct values reported during that time. At the end of each two-week window, we re-fit the model using all Ct values and incidence data up to that time point, then nowcast the next two-week window, and so on. As a sensitivity analysis, we also compared RMSE and AUC with a fixed train-test split date on 31 Dec 2021 (**Table S2**).

### Nowcasting performance on synthetic datasets

We first applied these GAMs to several synthetic Ct value datasets, generated as numerical implementations of the analytical convolutions presented in the previous section. These synthetic datasets combined a range of viral kinetics models and sampling regimes with population-level reported SARS-CoV-2 incidence curves for Massachusetts, USA to produce linelists of individual-level test results and Ct value observations (see **Supplementary Text 1**; **Figure S3&4)**. Using synthetic datasets in this way allowed us to incorporate or exclude the effects of confounding factors on observed population-level Ct value distributions, in addition to the effect of the epidemic trajectory itself (see **Table S1**). The simulations generated approximately 22,000 positive tests under asymptomatic screening (mean of 20 observations per day, range 1-250) and 250,000 positive tests under symptom-driven testing (mean of 230 observations per day, range of 3-2700), over 1000 days.

We simulated two main synthetic datasets: 1) an ideal scenario where a strong Ct value/growth rate relationship is expected, assuming a) highly asymmetric viral kinetics, with a very short growth phase and long clearance phase; b) low variation in observed viral load/Ct value for a given time-since-infection; and c) a uniform probability of testing an individual at any time up to 7 days post symptom onset or tested once at random during the simulation regardless of infection status, and 2) a realistic baseline scenario assuming a) increased symmetry in viral kinetics, with a growth and clearance phase duration based on ancestral SARS-CoV-2 (**Figure S5**); b) moderate variation in observed viral load/Ct value for a given time-since-infection; and c) individuals are tested after a low-variance, gamma-distributed delay following symptom onset or once at random during the simulation (**Figure S6**). We also performed sensitivity analyses on datasets with variations of these three factors (**Figure S6**), as well as a dataset incorporating viral kinetics across epidemic waves to represent differences in viral variants (**Figure S7**). The standardized mean Ct value within each simulation and testing strategy (subtracting the overall mean Ct and dividing by the standard deviation) showed the same trend over time for all simulations, excepting the scenario assuming a symmetric viral kinetics model (**Figure S8&9**).

Both ideal and realistic synthetic datasets had a negative correlation between the 7-day rolling average epidemic growth rate of cases and 7-day rolling average mean Ct value (Figure 2A**; Figure S4B**). Ct values observed through symptom-based testing were typically lower and exhibited less variation than those observed through random testing. These results suggest that a strong relationship between mean Ct value and epidemic growth rates should be identifiable, at least in theory.

**Figure 2.**
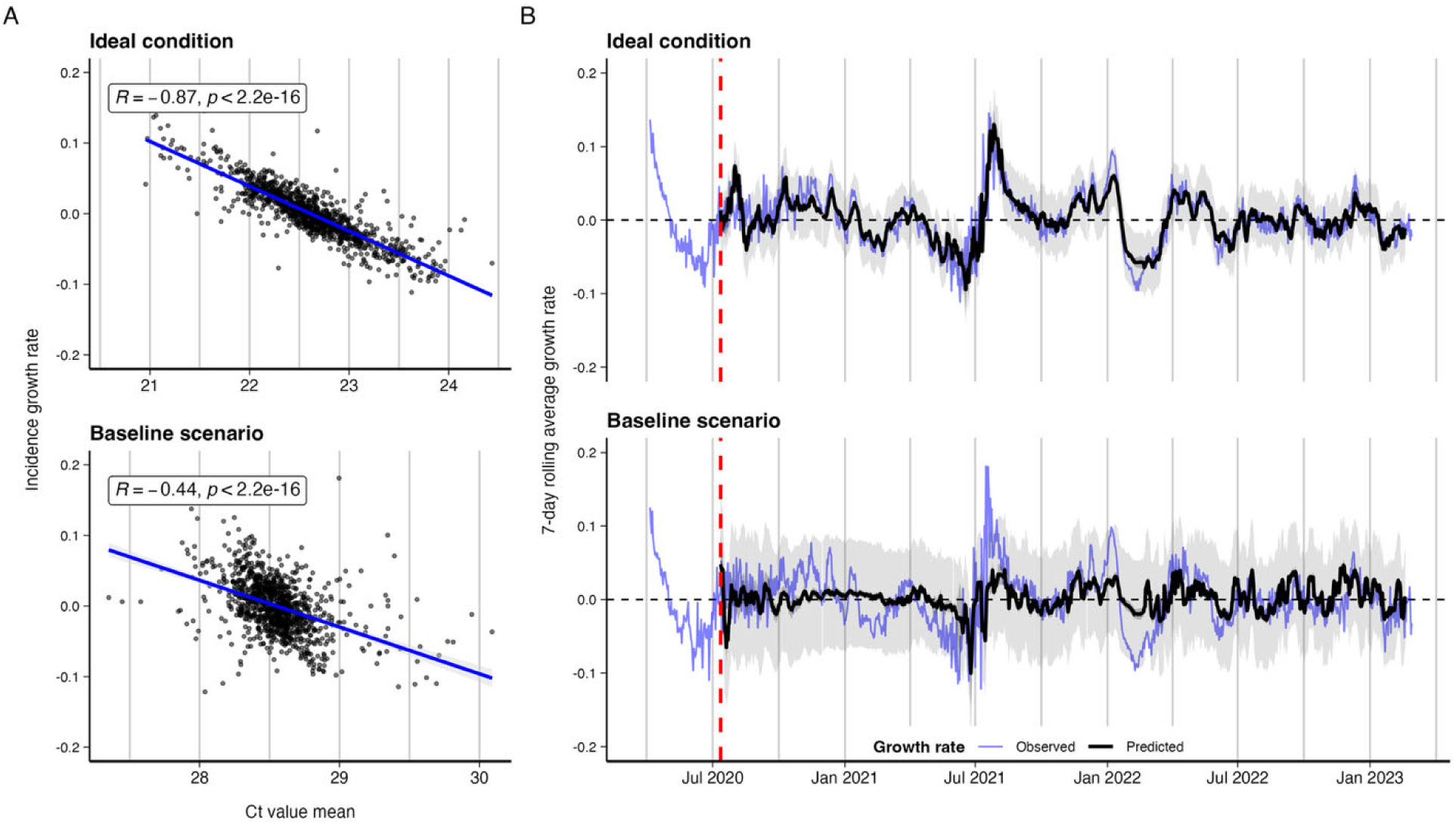
(**A**) Scatterplot showing 7-day rolling mean Ct value against 7-day rolling mean incidence growth rate from the two main synthetic datasets. Inset text shows Pearson correlation coefficient. Blue line shows fitted linear regression line with 95% CI. (**B**) Model-predicted (black) vs. observed (blue) log incidence growth rates for the two main synthetic datasets, with model-predicted 95% confidence intervals (dark shading) and 95% prediction intervals (light shading). Predictions are from 2-week rolling nowcasts, concatenated into a single time series. Vertical dashed line denotes the end of the initial training period.

With the ideal synthetic dataset, the estimates from the GAMs closely tracked observed growth rates using Ct value means and skew, with an in-sample RMSE of 0.0181—approximately 10% of the range in observed log incidence growth rates (**Figure S10&11**; **Table 1**)—as well as accurately predicting epidemic direction (in-sample AUC = 0.936). Nowcast accuracy over the two-week windows were slightly worse than the in-sample predictive performance (mean across all nowcast windows, RMSE = 0.0192, AUC = 0.926) but still accurately tracked the epidemic over the full time period (Figure 2B). In comparison, a null model using the mean growth rate over the preceding two weeks as the prediction for the following two-week window gave an RMSE of 0.0336 and AUC of 0.770.

**Table 1.** Predictive performance of GAMs using synthetic datasets, predicting per-day growth rates from daily Ct value statistics.

| **Dataset** | **RMSE, in-sample** | **RMSE, now-cast** | **AUC, in-sample** | **AUC, now-cast** |
| --- | --- | --- | --- | --- |
| *Ideal condition* | 0.0181 | 0.0192 | 0.936 | 0.926 |
| *Baseline scenario* | 0.0339 | 0.0348 | 0.728 | 0.683 |
| *Realistic kinetics* | 0.0309 | 0.0333 | 0.798 | 0.752 |
| *Realistic variation* | 0.0231 | 0.0251 | 0.863 | 0.832 |
| *Realistic sampling* | 0.0195 | 0.0220 | 0.920 | 0.893 |
| *Symmetric kinetics* | 0.0326 | 0.0355 | 0.742 | 0.682 |
| *Previous 2-week mean growth rate carried forward* | NA | 0.0336 | NA | 0.770 |

Model predictive performance was worse when using the realistic baseline synthetic dataset, with an in-sample RMSE of 0.0339 and AUC of 0.728 (**Figure S10&11**; **Table 1).** Of the three factors examined individually, the symmetry of the viral kinetics model had the largest impact on model performance, resulting in the greatest increase in RMSE and decrease in AUC (**Figure 20-12**, **Table 1**). When these models were applied to nowcasting growth rates in two-week increments, the greatest performance reduction again occurred when using either a realistic or symmetric viral kinetics model (**Table 1**). Nowcasting predictive performance for the realistic baseline scenario was similar to the null model using the mean growth rate of the preceding two weeks as the prediction for the following two-week window. In the sensitivity analysis assuming different variant waves, nowcasting performance was poor, as unaccounted for changes to the Ct/growth rate relationship due to changes to within-host viral kinetics resulted in biased predictions (**Figure S13**).

### Prediction accuracy is increased with higher sample sizes in the training set

Despite the clear theoretical relationship between mean Ct value and the epidemic growth rate shown in Figure 1 and **Figure S4**, prediction accuracy using synthetic Ct values under realistic assumptions was relatively poor. To understand why, we further examined the relationship between sample size and model predictive performance, evaluating outputs from a range of positive tests each day under the realistic baseline scenario collected under either random testing or symptom-based testing. We compared a) in-sample predictive performance fitting the GAM to the 7-day rolling average of the mean and skew of Ct values for each sample size and b) nowcasting predictive performance, varying both the number of samples used to train the model and subsequently collected each day for prediction during the nowcasting windows.

These simulations showed three patterns. First, in-sample predictive performance (assessed by RMSE) improved substantially as the number of daily positive samples increased from 25 to 1000 (Figure 3), reaching similar RMSE values to the ideal synthetic scenario (**Table 1)** with 100 positive samples collected at random per day or 500 samples through symptom-based testing. Second, both in-sample and nowcasting predictive performance was greater for the same sample size using Ct values collected at random compared to symptom-based testing (Figure 3**, Figure S14&15**). The lower sample size requirement under random testing reflects the shallower gradient between mean Ct value and growth rate compared to symptom-based testing, as described in the section *Theoretical relationship between epidemic growth rates and Ct value statistics*. Third, increasing the number of positive samples available to predict growth rates did not compensate for having too few samples available to train the model in the first place (**Figure S14&15**). Model predictions were only accurate if the relationship between growth rates and Ct values was accurately characterized to begin with. These findings likely explain the limited performance under the realistic baseline scenario, where sample sizes were around 20 positive tests per day under random testing and around 250 for symptom-based testing, but with wide variation depending on incidence (approximately 10x higher at peak incidence).

**Figure 3.**
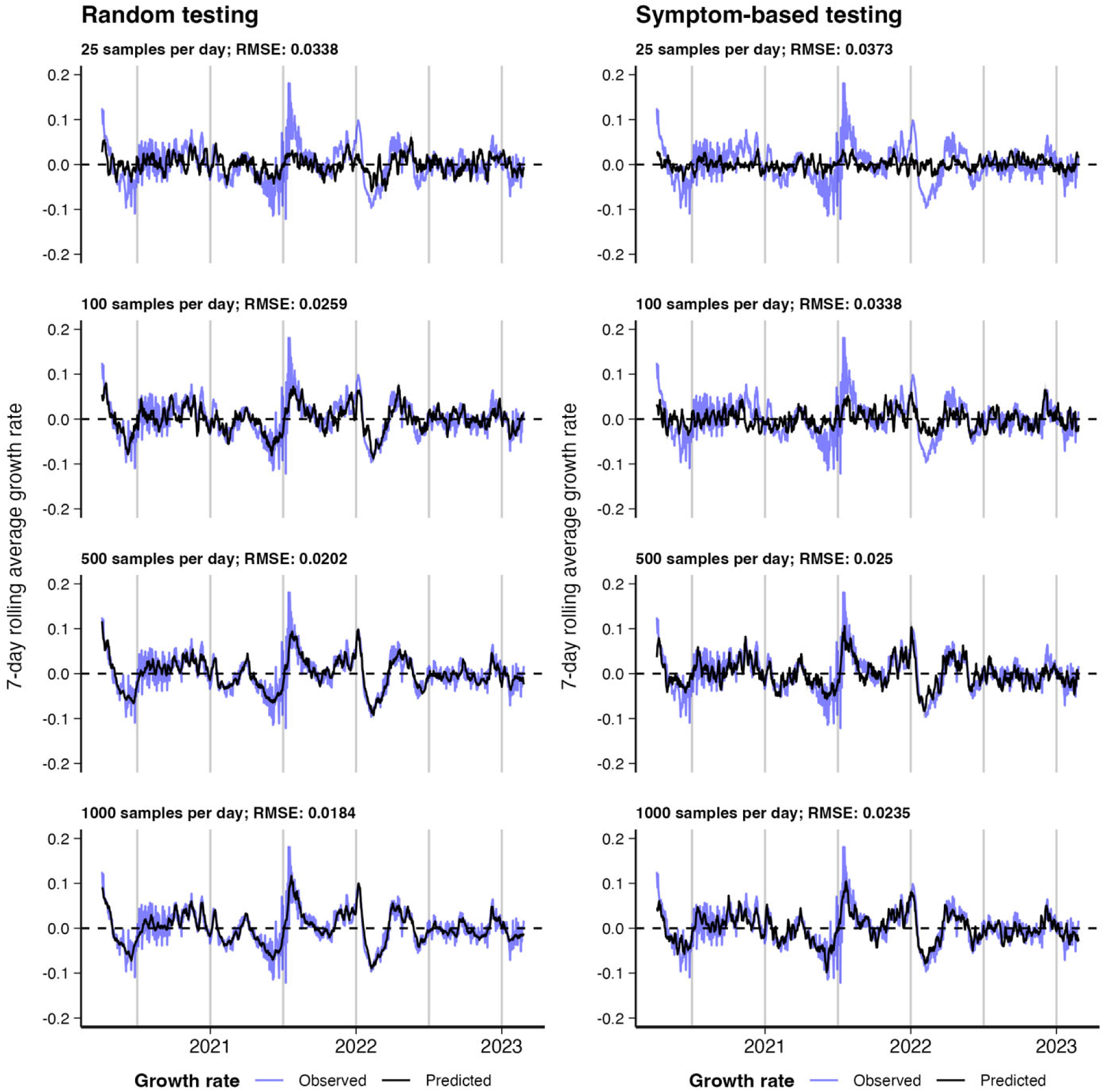
Comparison of in-sample predicted incidence growth rates against observed growth rates using simulated 7-day rolling average of the mean and skew of Ct values as a predictor. Columns distinguish simulated data obtained under a testing at random system and data obtained under a symptom-based testing system. Rows show increasing number of simulated daily observations used to train the model.

### Real-world relationship between observed Ct-value statistics and epidemic trajectories

Having established a baseline for model nowcasting performance using the synthetic data, we next tested the nowcasting models on two real RT-qPCR datasets: 1) routine hospital testing data from the Mass General Brigham hospital system in eastern Massachusetts (MGB), spanning Mar 2020-Feb 2023, and 2) municipal testing data from Los Angeles County, California (LAC), spanning May 2020-Jul 2021 and Jan-Sep 2022 (see **Supplementary Text 3**). The MGB data came largely from mandatory screening testing of outpatient, inpatients and emergency room admissions, while the LAC data were primarily symptom-driven voluntary testing (**Table S3**). Both datasets contained specimen collection dates and Ct values for SARS-CoV-2 positive results; LAC data also included vaccination status, symptom status, and symptom onset dates. MGB Ct values came from seven platform/assay combinations, while Ct values from LAC data came from one PCR platform with two possible assays (**Table S3**).

We limited our analysis to tests reporting Ct values, using the first available recorded Ct value for each infection episode. The final analyzed sample included 104,534 (MGB) and 279,492 (LAC) Ct values. We compared these Ct values against reported COVID-19 incidence for Massachusetts and Los Angeles County, respectively. We segment the data into four ‘variant eras’ based on the SARS-CoV-2 variant known or believed to be dominant in the U.S. during different approximate time periods, to allow for differences in viral kinetics by variant.

Ct value distributions from both MGB and LAC datasets showed substantial variation over the course of the pandemic (Figure 4A **& Figure S16A**). Reported COVID-19 incidence in both locations varied over time, as well (Figure 4B **& Figure S16B**), with large infection waves in the winters of 2020-21 and 2021-22, though the pattern of incidence was not synchronized across both settings. While absolute incidence varied widely, incidence growth rates remained largely between ±0.2 throughout the course of the pandemic (Figure 4B **& Figure S16B**).

**Figure 4.**
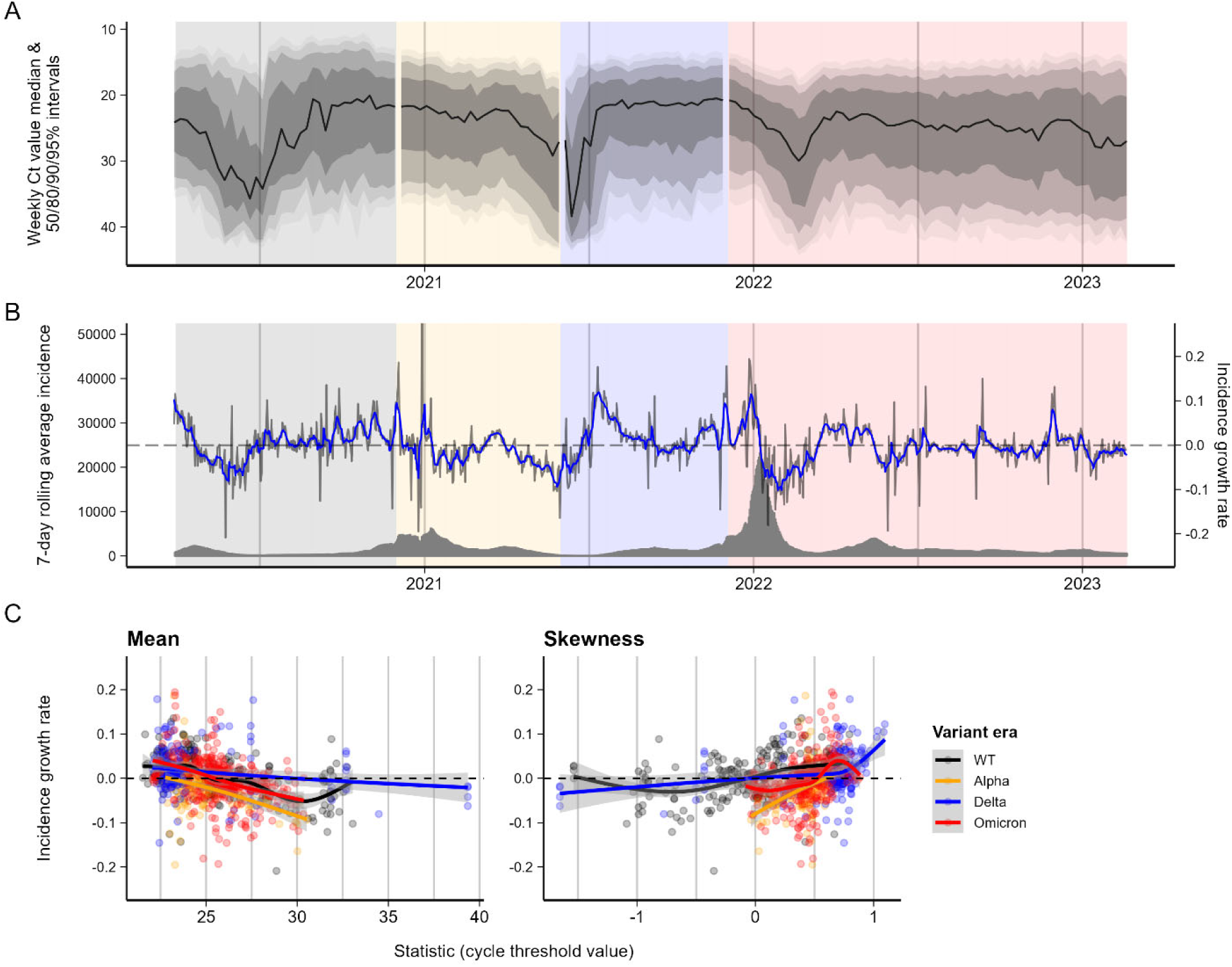
Ct values from the Mass General Brigham hospital system and corresponding reported COVID-19 incidence in Massachusetts, USA. (**A**) Weekly Ct value quantiles over time, showing weekly median Ct value and 50/80/90/95% quantiles. (**B**) 7-day rolling average reported incidence (grey bars), growth rate in 7-day rolling average reported incidence (grey line), and smoothed growth rate (blue line). Background is shaded by time periods of different variant dominance. Vertical dashed line demarcates the test-train split. **(C**) Incidence growth rate compared to smoothed daily mean and skewness of Ct value distributions. Colored lines and shaded grey regions show fitted cubic spline GAMs with 95% confidence intervals, stratified by period of variant dominance.

We found the mean and skewness of observed Ct value distributions (calculated daily over a seven-day moving window and excluding days with fewer than 10 Ct values reported) negatively correlated with the growth rate in reported incidence (Figure 4C **& Figure S16C**). Analysis of cross-correlation functions found Ct value distributions lagged incidence growth rate in the MGB data, with strongest correlations at around 19-days lag (autocorrelation function, ACF= -0.462), and led incidence growth rates for the LAC data, with strongest correlations at around 10-days lead (ACF = -0.062) (**Figure S17 & Figure S18**). However, for real-time nowcasting, we focused on the relationship between same-day Ct values and incidence (i.e., lag=0 days; Figure 4C **& Figure S16C**), which still showed significant correlation. Higher incidence growth rates corresponded with lower same-day average Ct values (Spearman’s correlation coefficient: MGB Rho = -0.43, LAC Rho = -0.22) and with positively skewed Ct distributions (MGB Rho = 0.35, LAC Rho = 0.43).

### Nowcasting epidemic growth rates using Ct values in Massachusetts, USA and Los Angeles County, USA

We next re-trained the same GAM models used with synthetic data to the MGB and LAC dataset using smooth functions of mean and skewness of Ct values to predict log incidence growth rates, with corresponding logistic models to predict epidemic direction. Model predictions were compared against observed values first in-sample across the entire dataset, over a rolling two-week prediction window with an expanding training period encompassing all preceding weeks, and with a single fixed train-test split date on 31 Dec 2021.

In both datasets, this simple model achieved in-sample prediction accuracy for incidence growth rate slightly worse than performance on the realistic synthetic data, with relatively small absolute errors (MGB RMSE = 0.0451; LAC RMSE = 0.0335, see **Table *2***, **Figure S19-S22**, and **Table S4**). Corresponding logistic regression models successfully discriminated growing from declining incidence (Area under the curve: MGB AUC = 0.785, LAC AUC = 0.843).

**Table 2.**
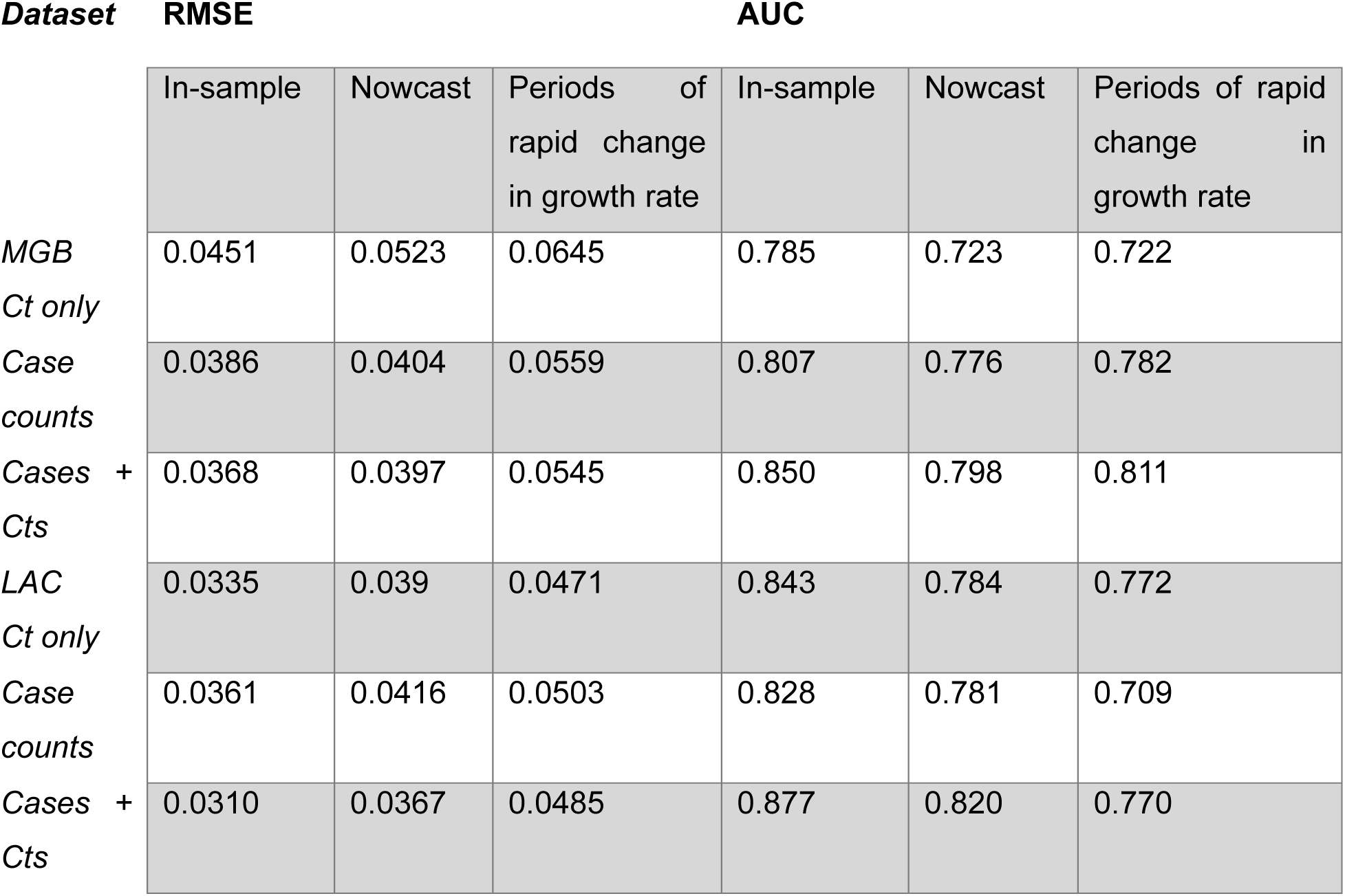
Predictive performance of the selected GAM using data from MGB and LAC, predicting per-day growth rates from daily Ct value statistics.

The models were able to nowcast growth rates, in two-week increments with models periodically refitted to more recent data, with accuracy slightly worse than in-sample model fits (MGB RMSE = 0.0523, LAC RMSE = 0.039) (Figure 5A **& Figure S23A**). This level of nowcast accuracy was likewise slightly worse than nowcasting performance with realistic synthetic data. While average prediction error was relatively small, comparable to in-sample model error and to prediction error with realistic synthetic data, accuracy was highly variable from one two-week window to the next (Figure 5B **& Figure S23B**). Nowcast accuracy was comparable to model performance over a fixed multi-month prediction window, slightly better for one dataset and worse for the other (MGB RMSE = 0.047, LAC RMSE = 0.0458; **Table S2**). Nowcast predictions of epidemic direction were slightly worse than in-sample ones (MGB AUC = 0.723, LAC AUC = 0.784) and outperformed the directional predictions for realistic synthetic data. In addition, over all two-week nowcast windows combined, model-predicted growth rates correlated moderately well with observed ones (Spearman’s Rho: MGB Rho = 0.398, LAC Rho = 0.556).

**Figure 5.**
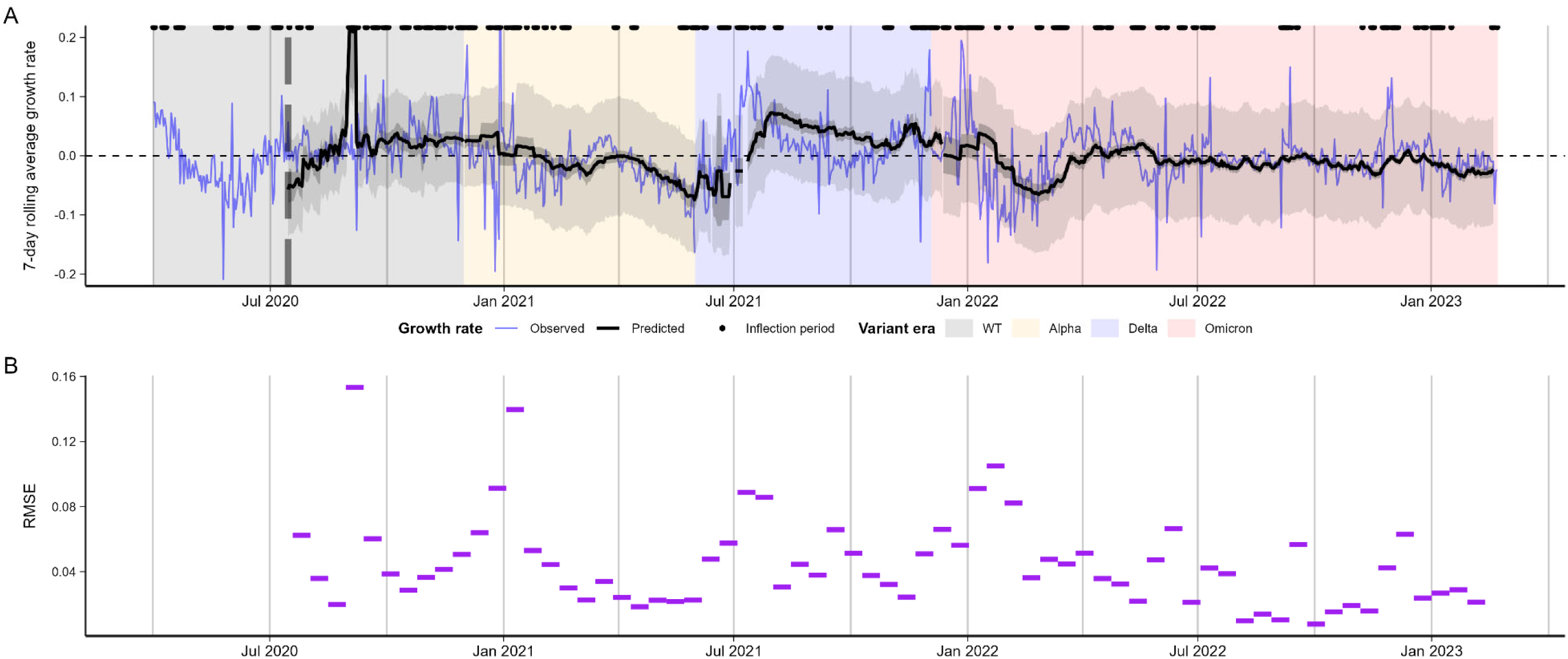
**(A)** Model-predicted (black) vs. observed (blue) log incidence growth rates for MGB data, with 95% confidence intervals (dark shading) and 95% prediction intervals (light shading). **(B)** RMSE of predicted vs. observed log incidence growth rates for each 2-week nowcasting window. “Inflection periods” refer to times when the absolute smoothed log incidence growth rate exceeded 0.025 over a one-week period, marked with points above each subplot.

### Nowcasting state- or county-level epidemic growth rates combining Ct values and number of new positive tests

To understand what information Ct values add over simply counting new positive tests, we evaluated GAM and logistic models which included smoothed log incidence growth rate of positive tests within each dataset as a predictor. In all cases we aimed to predict state- or county-wide incidence trends using data from a single reporting system (i.e., MGB hospital testing or LAC community testing). These within-dataset growth rates would be expected to correlate strongly with population-wide growth rates provided they are a representative sample. We compared model predictions using only Ct value statistics, only growth rate of positive tests, or combined Ct value statistics and positive tests.

For both the MGB and LAC datasets, the models incorporating both positive tests and Ct value mean and skew had the greatest prediction accuracy (MGB RMSE = 0.0397, LAC RMSE = 0.0367), close to the accuracy achieved with realistic synthetic data, indicating that both number of positive tests and Ct value distributions were informative about incidence growth rates. However, for the MGB dataset, the model using growth rate of positive tests alone performed substantially better than the model using Ct mean and skew alone (RMSE 0.0404 vs. 0.0523). In contrast, for the LAC dataset, the model using only Ct mean and skew outperformed the model using positive test data only (RMSE 0.0390 vs. 0.0416). The same rank ordering of nowcast performance holds for epidemic direction prediction accuracy as measured by AUC.

### Sensitivity analyses

We performed several sensitivity analyses, assessing nowcasting performance under various sample restrictions and stratifications. First, we restricted model performance assessment to periods of rapid change in the epidemic trajectory (defined as days when absolute smoothed incidence growth rate exceeded 0.025 over a one-week period). Across both datasets, prediction error over periods of rapid change was greater than over the whole nowcast period (MGB RMSE = 0.0645 [rapid change] vs. 0.0523 [nowcast], LAC RMSE = 0.0471 [rapid change] vs. 0.0390 [nowcast]; see **Table *2***), while directional prediction accuracy was comparable (MGB AUC = 0.722 vs. 0.723, LAC AUC = 0.772 vs. 0.784).

Next, we assessed model performance on the same datasets stratified or restricted by covariates including 1) test setting (i.e., outpatient vs. inpatient vs. emergency room), 2) symptom status, and 3) immune history (accounting for both vaccination and past infection) (**Figure S24**). The relationship between Ct values and growth rate sometimes differed when subsetting or stratifying by these variables (**Figure S25**), but including these stratifications in the model did not always improve predictive performance. Restricting to outpatient tests only improved prediction error compared to baseline (nowcast RMSE = 0.0494 vs. 0.0523 base), whereas incorporating symptom status or immune history slightly worsened prediction error (nowcast RMSE = 0.0454 for symptom-stratified, 0.0415 for asymptomatic/no symptom status only, 0.0401 for immunologically naïve only, vs. 0.039 base, see **Table S6**).

### Nowcasting performance with variable sample size and outlier removal

Finally, we assessed the sensitivity of nowcasting performance to sample size both by randomly downsampling the MGB dataset (100 random draws) and by analyzing a third, smaller dataset from Tufts Medical Center—a hospital from the same region of Massachusetts—using the same response variable (i.e., log incidence growth rates for Massachusetts) but with approximately 10% of the total sample size of the MGB data (see **Supplementary Text 4**; **Figure S26&27**). In most cases, prediction accuracy for incidence growth rate was comparable with the downsampled datasets and the equivalent full datasets (Figure 1; see also **Table S5**). Only with 10% of the full dataset (but not with the Tufts dataset) did nowcasting accuracy degrade appreciably, though with a lot of variation between draws; with 50-75% downsampling or a daily maximum of 25 positive samples, accuracy improved slightly compared to baseline. Likewise, directional prediction accuracy was generally similar between downsampled and full datasets, with substantially worse accuracy only for the 10% and 25% downsamples. Improved accuracy may reflect reduced influence of outliers – downsampling the full dataset tends to exclude the days with smallest sample sizes, which are otherwise given equal weight in model training to days with more observations, while sub-sampling each day’s observations reduces the impact of outliers on each day’s observed Ct value distribution. To test this, we examined model performance with trimming of outlier Ct values from each day’s observed data. Trimming outliers reduced prediction error with 2.5%, 5%, and 10% trims (Figure 1, **Table S5**), while 2.5% and 5% trims also improved directional prediction accuracy.

**Figure 1.**
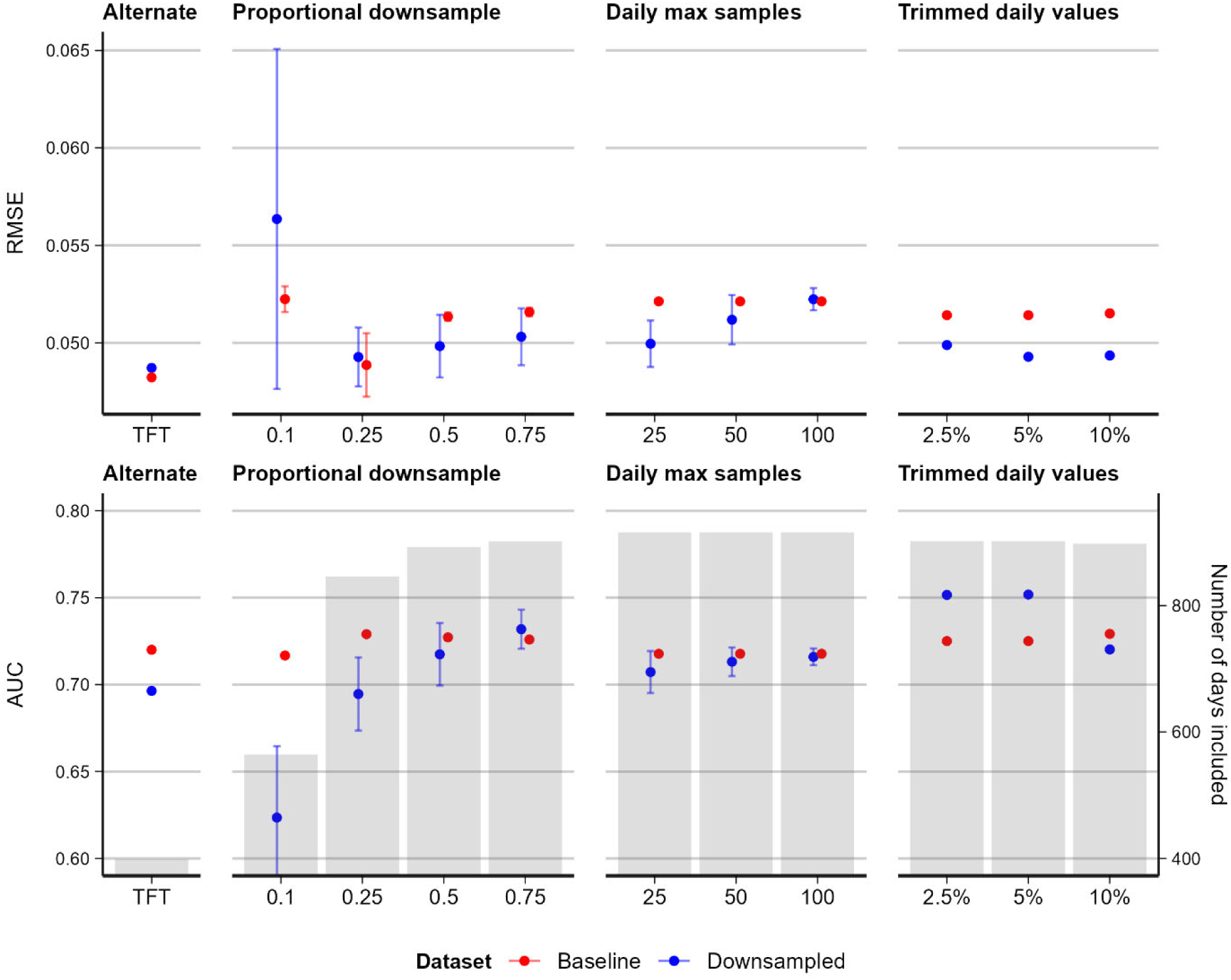
Model performance for downsampled MGB and full Tufts datasets. Baseline comparison metrics are re-calculated for only the days included in each downsampled dataset’s nowcast. For proportional and daily max downsampling, both downsampled and baseline performance are averaged over 100 random draws (and their corresponding days included). Trim percentages indicate quantiles trimmed from each end of daily Ct value distributions (i.e., 5% trim yields the 5-95 percentile range of Ct values). Grey bars indicate the number of days of data included for the various downsampling analyses; the random downsampling can reduce some days below the 10-sample inclusion threshold, hence the variation.

We also repeated these analyses on the models using within-dataset positive test counts instead of, or in combination with, Ct value distributions. In both instances, but particularly for the models using only within-dataset positive tests, reducing sample size substantially worsened model predictive performance for both growth rate and epidemic direction (**Figures S28&29**). Greater reductions resulted in greater performance degradation, indicating that models relying on the growth rates of case counts in available data are sensitive to sample size. Similarly, the models relying on only case counts within the Tufts dataset performed substantially worse than models using the full dataset.

## Discussion

Our results show that despite a clear theoretical basis for the use of Ct values collected through routine surveillance to nowcast epidemic growth rates,^41–43^ predictive performance depends on a range of factors including viral kinetics, variation in viral load for a given time-since-infection, sampling strategy, and number of positive samples. Under real-world conditions, simple generalized additive models using the mean and skewness of recorded Ct values could nowcast (log) incidence growth rates with prediction errors (RMSE) of approximately 0.04-0.05; slightly worse than a realistic simulated scenario (RMSE ∼0.03) and substantially worse than an ideal simulated scenario (RMSE ∼0.02). Across both real-world settings (Massachusetts and Los Angeles County), growth rates generally varied between approximately ±0.2, so this level of accuracy in modelled estimates, while not highly precise, is nonetheless informative. These same models also identified whether incidence is growing or shrinking with AUC greater than 0.7, substantially better than chance. Nowcast accuracy over two-week time horizons is slightly worse than in-sample model fits, especially early in the emergence of new dominant viral variants, whose effect cannot yet be accurately estimated. During periods of rapid change in incidence growth rate (e.g., just as a new outbreak wave is developing), nowcast accuracy for growth rate is slightly worse, possibly due to larger absolute growth rates during such periods. Crucially, however, directional predictions remain moderately accurate during those times (AUC > 0.7), supporting the use of Ct-value based nowcasts for situational awareness.

Our results corroborate findings from other settings, where Ct values have been used successfully to infer epidemic growth rates or reproduction numbers^28–38^. Our analysis builds on these studies with one of the largest empirical tests of this nowcasting approach to date using data from two locations in the USA over a three-year period. Epidemic growth rates and directions were nowcasted with reasonable accuracy using both datasets, despite showing different Ct value trends and capturing different populations and testing approaches, highlighting the generalizability of this approach. Furthermore, these data covered a long-time window and included periods of different variant dominance and population immunity, suggesting Ct values could continue to augment infectious disease surveillance as SARS-CoV-2 epidemiology continues to change.

Compared to nowcasting using growth rates in reported case counts within the real-world datasets we examined, Ct-value based nowcasts yielded worse accuracy in one setting (MGB data) and better accuracy in another (LAC data). Insofar as a testing dataset reflects a representative surveillance sample of a broader population, incidence growth rates within the dataset should accurately predict population-level incidence growth rates; the less representative the sample, the less reliable this relationship becomes. The MGB dataset, which included primarily routine screening results from all hospital outpatients, may have been closer to a representative sample of the broader population than the LAC dataset’s voluntary and primarily symptom-driven testing, which would explain the latter’s lower nowcasting accuracy vs. Ct-value based nowcasting. Our analyses of reduced sample sizes corroborate this idea – with smaller sample sizes (which would result in greater error), nowcasting accuracy of models using case counts degraded noticeably, whereas accuracy of Ct-value models was robust to smaller samples. These contrasts highlight one of the strengths of the Ct value-based approach: it does not depend on sampling positive cases proportional to incidence in the underlying population, and thus could be particularly useful when surveillance sampling is impractical (or when smaller datasets are readily available).

While the relationship between sampled viral loads, viral kinetics, and epidemic dynamics can be described mathematically under ideal conditions, in practice several issues complicate its application as an epidemic monitoring tool. Measured Ct values are determined by a combination of biological factors, such as individual-level heterogeneity, immunological history and infecting variant ^45^, and practical factors such as whether individuals are tested at a random point in their infection or around the time of peak viral load (prompted by symptom onset)^46^, demography of the tested population^47^, sample type^30,33,34^, and RT-qPCR platform^22,31,36,48^. Our synthetic data analyses help disambiguate some of these confounding factors by elucidating their effects on the underlying relationship between Ct distributions and growth rates, and comparing resultant impacts on predictive performance. For example, increasing the amount of variation in Ct values for a given time-since-infection increased the RMSE of our model predictions, and simulating a viral kinetics trajectory with similar growth and clearance durations resulted in a weaker relationship between mean Ct value and epidemic growth rate.

Predictive performance was slightly worse in the real datasets compared to a ‘realistic’ synthetic dataset. One key contributor in our real datasets is that Ct values were generated using multiple RT-qPCR assays and/or platforms and were not standardized, limiting the comparison of Ct values across platforms and assays (see Methods)^47–49^. Additionally, the data-generating model for our ‘realistic’ synthetic dataset did not incorporate the impact of vaccination or past infection which affect individual viral load trajectories^42,50^, potentially contributing to the differences in performance between models with empirical vs. synthetic data. Real-life implementations of Ct value nowcasting could potentially include covariates to account for these factors. Ultimately, predictions may become biased when the relationship between observed Ct values and the epidemic growth rate differ between training and test data. For example, if there are strong, time-varying correlations between symptom status, symptom onset, testing delay, and a time-varying mixture of symptomatic vs. asymptomatic samples, then the prediction model may become mis-specified over time if not updated.

Our synthetic data analysis also highlights the importance of sampling delays. Fundamentally, the relationship between population-level epidemic dynamics and viral load distributions arises because individuals’ viral loads reflect times since infection^22,31,36,50^, and hence cross-sectional distributions of viral loads (or Ct values) reflect the distribution of times-since-infection among currently infected individuals, similar to the relationship between incidence and prevalence. This relationship holds if individuals have a uniform probability of sampling any time after infection; such random cross-sectional samples are rare^22,31,36,51^ but are approximated in our datasets by routine screening of hospital outpatients. However, a more realistic sampling delay distribution – such as if individuals tend to be tested shortly after suspected exposure or developing symptoms – biases the probability of sampling over time since infection and dilutes the signal of infection age. Symptom-driven testing, where individuals are tested due to recent symptom onset beginning at around the same time as peak viral load, is the most common source of data used for epidemiological surveillance, which reduces any epidemic signal in the population-level Ct distribution. Changes in public health recommendations around testing and screening algorithms, such as recommendations around pre-travel testing or hospital admissions screening, may therefore change the relationship between population Ct values and epidemic dynamics, which may bias Ct-based epidemiological estimates, if not accounted for.

PCR platform differences and nonrandom sampling regimes are both addressable challenges, at least in principle. Ct value data could be calibrated across platforms and assays using standardized samples. Random surveillance sampling could reduce the bias in testing delay found with symptom-driven testing. True random sampling may be important, as voluntary testing by asymptomatic individuals may still show some bias in testing delays (**Table S6**). When we approximated these changes by subsetting one of our datasets to only results from outpatient screening tests, which were largely collected and analyzed the same way (**Figure S25**), we found small improvements in model predictive performance compared to using the full, mixed dataset (**Table S6**). While random surveillance sampling at low prevalence may yield very few infections detected, nowcasting accuracy was not severely degraded even with substantially reduced sample sizes (Figure 1), though crucially, model training requires sufficient data. Both these changes would improve the accuracy of simple Ct-based nowcasting models. Even absent such logistical solutions, however, we found the simple statistical heuristic of trimming outliers (2.5-5%) from daily observed Ct values improves nowcasting accuracy (Figure 1).

Beyond confounding factors, it is plausible that the growth rate of reported COVID-19 cases may not be the most accurate benchmark against which to compare Ct value distributions. Reporting rates and delays may also change over time, causing deviations between reported case counts and true infection incidence. Alternative benchmarks, such as growth rate in hospitalizations, mortality, or wastewater viral loads, may therefore yield stronger relationships (possibly with some time-shifting); investigating these relationships would be a fruitful avenue for further research. In addition, geographically aggregated incidence may mask heterogenous outbreak trajectories at finer scale, e.g., city or even neighborhood level. Such finer-scale incidence data may yield cleaner relationships with Ct value distributions, especially if matched to the catchment areas for the Ct value data collection process.

Another challenge for modeling Ct value dynamics is the choice of mathematical model to capture the relationship between observed Ct values and underlying epidemic growth rates. With random sampling, the link between epidemic dynamics and observed viral loads can be described precisely based on the convolution of the infection incidence curve and viral kinetics curve^51^. In contrast, viral loads observed through non-random or convenience samples, such as symptom-driven testing, arise from complex data generating processes which are difficult to describe mathematically, and thus past studies, including ours, tend to favor regression models or machine learning algorithms to estimate epidemic dynamics from observed Ct values^52^. Our experience is that the convolution framework presented in ^21^ can more reliably estimate incidence from Ct values, as it precisely describes the relationship between incidence, cross-sectional viral load distributions and viral kinetics, but it requires external data to parameterize a viral kinetics model and becomes computationally infeasible as the data-generating process becomes more complex. Thus, future work should also focus on more complex statistical methods that accounts for the time-series nature of the data^37^, the non-linear and potentially non-monotonic relationship between Ct values and growth rates, and combine multiple data streams to provide more accurate predictions of epidemic dynamics^54^. Future work should also consider the theoretical findings presented here: growth rate estimation is inherently limited by how reliably moments of the Ct value distribution are measured (sample size and variance), as well as the strength of the relationship between Ct value metrics and epidemic growth rates.

Tracking epidemic growth rates in near-real-time remains an important challenge for public health surveillance. Our analyses show that simple Ct-based models can track SARS-CoV-2 epidemic growth rates, highlighting their potential use in augmenting infectious disease surveillance systems. Ultimately, their greatest strength lies in their speed and simplicity. The models presented here are conceptually straightforward and computationally lightweight, easy to implement even in resource-constrained settings, and, unlike wastewater testing, are reliant only on data already routinely collected as part of screening or diagnostic testing. Our analyses show that they retain their accuracy even with varying sample sizes or during periods of rapid change in epidemic trajectories, such as during the transition from the end of one epidemic wave to the start of the next one and thus could provide rapid situational awareness as outbreak waves emerge. Further research could examine how Ct-based estimates of epidemic trajectories complement other, orthogonal indicators such as wastewater surveillance, as well as potential applications to different viral pathogens with well-characterized viral kinetics such as influenza or RSV^55^.

## Methods

### Study settings & data sources

#### Massachusetts

Massachusetts Ct value data comes primarily from testing in 16 hospitals in the Mass General Brigham hospital system. The full dataset comprises 2,671,041 SARS-CoV-2 test results. Limiting to results reporting Ct values and first reported Ct values for each confirmed case yields the final sample of 104,534 Ct values used in this analysis (**Table S3**), of which the earliest specimens were collected on 31 Mar 2020. Approximately 75% of samples were from routine outpatient screening. Specimens included nasal and nasopharyngeal swabs, processed using seven different RT-qPCR platform/assay combinations; Ct values were pooled across platforms/assays.

We did not have access to information on patients’ vaccination or infection history, infecting variant, or symptom status. More details on the dataset and sample restriction are in **Supplementary Text 3**.

Daily confirmed case counts for Massachusetts were obtained from the Massachusetts Department of Public Health COVID-19 dashboard^55^.

We also analyzed a secondary dataset of Ct values from Tufts Medical Center in Boston, Massachusetts for comparison. The full dataset comprised 84,848 test results (10,338 positive) with collection dates ranging from 18 Feb 2021 to 31 Oct 2022; the final sample included 10,214 Ct values (see **Supplementary Text 3**). **Figure S27** summarizes the reported Ct value distributions over time and compares these to reported COVID-19 incidence.

### Los Angeles County

LAC Ct value data comes from municipal COVID-19 testing sites operated by the LAC Department of Public Health and Department of Health Services, comprising approximately 10% of all municipal testing conducted in LAC during the sample period. The full dataset comprises 330,034 SARS-CoV-2 positive test results, with specimens collected over two time periods – 21 May 2020 to 27 Jul 2021, and 30 Dec 2021 to 29 Sep 2022. (Note: data were unavailable for the intervening period.) Limiting to the first reported Ct value for each infection episode yields the final sample of 279,492 Ct values used in this analysis. Specimens were collected through nasal, nasopharyngeal, and oral swabs, and analyzed by Fulgent Genetics using two RT-qPCR assays (see SUPPLEMENT). Symptom status was reported for approximately 75% of the sample, of which in turn approximately 75% (56% of the full sample) are reported as symptomatic for COVID-19 (**Table S3**). For symptomatic cases, most specimens were collected 1-10 days after symptom onset (modal delay of 3 days). The sample also included vaccination status, with approximately 24% of results coming from vaccinated (partially, fully, or boosted) individuals (**Table S3**).

Daily confirmed case counts were obtained from the LAC DPH COVID-19 dashboard^56^.

### Synthetic datasets

We built on a previously published model to simulate realistic Ct value distributions that would be expected under testing and sampling schemes similar to real-world data^55^. The simulation combines 1) an individual-level viral kinetics model describing the expectation and distribution of Ct values over time following infection, using previously published longitudinal SARS-CoV-2 testing data (**Figure S5, Table S7**)^50^; 2) a population-level epidemic curve with approximately 2 million infections, with infection times distributed based on the reported incidence of COVID-19 cases in Massachusetts between 5 March 2020 and 25 Feb 2023; and 3) a surveillance model combining random testing (i.e., symptom-independent) and symptom-based testing. Different scenarios were captured by changing parameters for the viral kinetics model or sampling delay distribution (**Figure S6**). Full details of the simulation framework are given in **Supplementary Text 1**.

### Statistical methods

We calculated daily incidence-based epidemic growth rates as the natural log-transformed ratio of 7-day moving average new reported cases for each day to the 7-day moving average for the preceding day:

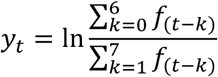

where 𝑦_t_ is incidence growth rate and 𝑓_t_ is daily incidence at time 𝑡. We defined epidemic direction as growing when 𝑦_t_ > 0 and declining when 𝑦_t_ ≤ 0.

### Classifying time periods of rapid incidence change

To identify periods of rapid change in incidence growth rate, we first smoothed the daily incidence growth rate (as defined above) using a centered 7-day moving average:

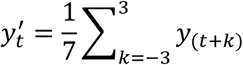

We then identified times when the absolute change in smoothed log incidence growth rate 𝑦’ equals or exceeds 0.025 over a one-week period, denoting the midpoint days of those weeks as periods of rapid change. That is, time 𝑡 is defined as having rapid change in incidence if and only if j𝑦’_(t+3)_ − 𝑦’_(t-3)_ j ≥ 0.025.

### Growth rate & epidemic direction models

We modeled incidence growth rate using a generalized additive model (GAM) incorporating the mean and skewness of Ct values:

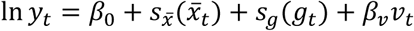

Where 𝑠_x̅_ and 𝑠_g_ are smoothing functions fitted using cubic regression splines^56^, and 𝑥̅_t_ and 𝑔_t_ are the 7-day rolling averages at time 𝑡 of the daily mean and skewness respectively of Ct values from samples collected or over the window from time 𝑡 to 𝑡− 6, excluding days with fewer than 10 Ct values reported. 𝑣_t_ is a categorical variable identifying the SARS-CoV-2 variant known or believed to be dominant in the U.S. during different approximate time periods. For our datasets, we designated four such variants / time periods: wild type (up to 30 Nov 2020), Alpha (01 Dec 2020 to 31 May 2020), Delta (01 Jun 2020 to 03 Dec 2021), and Omicron (04 Dec 2021 onwards). We used this rough approximation rather than relying on more direct and detailed observations, e.g. sequencing data linked to our datasets, to better represent a realistic use case for the Ct-based method such as a small municipal public health department. In such cases, resources for extensive sequencing may not be available, necessitating reliance on broader national trends. When encountering new variant[s] in a nowcasting or testing period not present in training data, our models use a realistic decision rule of making predictions based on the last known variant from training data. Details on the choice of model are in **Supplementary Text 2**.

We also modeled epidemic direction using the same set of GAMs used for incidence growth rate, but with a logit link function to predict binary outcome data (growing or shrinking epidemic):

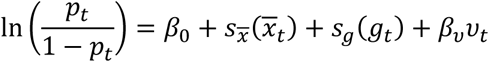

Where 𝑝_t_ is the probability that the epidemic is growing at time *t,* and declining otherwise. This is implemented by setting ‘family=binomial()’ in ‘mgcv:gam’.

### Evaluating model performance

To evaluate the performance of Ct-based nowcasting models, we conducted two model validation tests. First, we fitted the main models to each dataset using only data up to 31 Dec 2021 (training set), then used the fitted model to predict incidence growth rates and epidemic direction for the remainder of each dataset (test set from 01 Jan 2022 onwards), based on reported Ct values. We assessed prediction performance using RMSE between model-predicted and observed incidence growth rates and AUC for directional predictions from the logistic regression model.

Next, we conducted an expanding nowcast test, approximating a realistic application of this approach. For each dataset, we trained the main models on the first 16 weeks of available data, using the models thus fitted to predict incidence growth rates and epidemic direction over the following 2-week period using only reported Ct value statistics. We then re-fit the models on an expanded training window incorporating those two weeks of incidence data (i.e., up to 18 weeks) and predict the *subsequent* 2-week period, repeating this re-fitting and prediction procedure in 2-week increments up to the end of each dataset. A 2-week window was chosen to demonstrate how models could be updated on a rolling basis. In practice, the appropriate window would depend on multiple factors such as computational capacity and frequency of data updates. Shorter windows could improve predictive performance, by allowing faster detection of changes in the Ct-growth rate relationship. We report performance as RMSE or AUC across all 2-week prediction periods concatenated into a single prediction time series for each dataset and model, while detailed period-by-period performance is reported in the online repository at ^55^.

### Model performance comparison to within-dataset case count models

As a comparison, we also modeled incidence growth rate using a GAM incorporating the growth rate of case counts reported within each dataset:

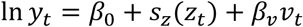

Where 𝑠_z_ is a smoothing functions fitted using cubic regression splines, and 𝑧_t_ is the natural log-transformed ratio of 7-day moving average new cases reported within the dataset for each day to the 7-day moving average for the preceding day; in other words, 𝑧_t_ is equivalent to 𝑦_t_ for cases reported within the sample dataset rather than the broader population. We also tested a combined model incorporating both within-dataset growth rate and Ct value mean and skewness:

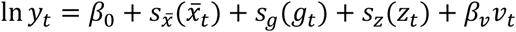

We modeled epidemic direction using the analogous logit link functions for the same models, as described above.

### Impact of reduced sample size and outliers on Ct-based growth rate estimation

As a sensitivity analysis, we repeated the rolling nowcast analyses using versions of the MGB dataset downsampled in two ways: 1) by randomly drawing 10/25/50/75% of the total test results available, or 2) by limiting the maximum number of positive test results for each day to 25/50/100, discarding any additional tests. We then reassessed nowcasting performance on each of these downsampled datasets, repeated with 100 random downsampling draws for each size. We also compared nowcasting performance using a third, smaller dataset from Tufts Medical Center, which uses the same response variable data as the MGB dataset (i.e., log incidence growth rates for Massachusetts) but has approximately 10% of the total sample size.

To assess the impact of outliers, we also trimmed daily observed Ct value distributions by 2.5/5/10% (yielding 95/90/80% ranges) before calculating Ct value distribution statistics, using the trimmed data for both training and nowcasting. Further details on downsampling and trimming are in **Supplementary Text 4**.

We also repeated these analyses using the models incorporating only within-dataset case count growth rate, as well as the combined models incorporating within-dataset case count growth rate along with Ct value mean and skewness, as described in the previous section.

## Data & code availability

Data and analysis code are available online at https://github.com/gradlab/ct-nowcasting [NOTE: we will update this to a Zenodo DOI before publication].

## Data Availability

All data produced will be available online at https://github.com/gradlab/ct-nowcasting once data sharing approvals have been finalized.

## Acknowledgements & financial disclosures

JAH is supported by a Wellcome Trust Early Career Award (grant 225001/Z/22/Z). This work was supported in part by the Francis P. Tally, MD, Fellowship in the Division of Geographic Medicine and Infectious Disease (JAP). This project has been funded in part by contract 200-2016-91779 with the Centers for Disease Control and Prevention (CDC). Disclaimer: The findings, conclusions, and views expressed are those of the authors and do not necessarily represent the official position of the CDC. The authors also thank Jason Cheng and Hanlin (Harry) Gao of Fulgent Genetics for assistance with data for the analysis.

## Author contributions

All authors meet *Nature* journal authorship requirements. TYL, YHG, and JAH conceptualized the project. TYL and JAH designed the analyses, developed the code, and created the visualizations. TYL, SK, JP, MH, THK, and PD prepared data. SK, SD, RF, and YHG provided resources and contributed to analysis design and interpretation. YHG provided primary supervision and funding support. TYL and JAH wrote the first draft. All authors provided critical review and revision of the text and approved the final version.

