## Supplementary information for "Nowcasting epidemic trends using hospital- and community-based virologic test data"

**Table S1.** Factors confounding Ct value inference

| Factor | Explanation | Scenario | | | | | |
|---|---|---|---|---|---|---|---|
| | | **1** | **2** | **3** | **4** | **5** | **6** |
| *Biological factors* | | | | | | | |
| Inter-individual variation in viral kinetics | Individuals' viral loads and viral load trajectories vary substantially, even accounting for immune history, demographic factors, etc. | 0 | 0 | X | 0 | X | 0 |
| Symmetry of viral load trajectory | While the viral growth phase is generally much shorter than the clearance phase, any single viral load measurement could come from either phase[1]. | 0 | X | 0 | 0 | X | X |
| Impact of immune history on viral kinetics | Past exposure through previous infection, vaccination, or both may result in faster viral clearance[2,3]. | 0 | 0 | 0 | 0 | 0 | 0 |
| Impact of demographic factors on viral kinetics | Older individuals generally have higher viral loads and slower viral clearance than younger ones[2,4]. | 0 | 0 | 0 | 0 | 0 | 0 |
| Impact of viral variant | Different SARS-CoV-2 variants may be associated with different viral load trajectories[2,3]. | 0 | 0 | 0 | 0 | 0 | 0 |
| *Logistical factors* | | | | | | | |
| PCR platform / assay | Ct values are not typically standardized across different PCR platforms and assays[5]; differences in individual testing protocol (e.g. location swabbed) could contribute further differences in measured viral loads. | 0 | 0 | 0 | 0 | 0 | 0 |
| Testing behavior and sampling regime or delay distribution | Cross-sectional Ct value distributions reflect the convolution of the distribution of true infection ages and the sampling delay distribution; if sampling delays are highly clustered (e.g. mostly 3-5 days after infection), observed Ct distributions will reflect primarily individual-level random variation rather than informative variation in infection ages[6]. Sampling regime (e.g. representative random sampling, contact-tracing based sampling, voluntary testing, hospital outpatient screening) would influence the sampling delay distribution – random sampling theoretically results in a uniform delay distribution, while e.g. symptom-driven voluntary testing results in highly clustered sampling delays. | 0 | 0 | 0 | X | X | 0 |
| Synthetic data scenarios are numbered as follows: 1) Ideal condition, 2) realistic asymmetry in viral kinetics, 3) moderate individual-level variation, 4) clustered sampling delay distribution, 5) highly symmetric viral kinetics, 6) realistic baseline | | | | | | | |

**Table S2.** Comparison of the 24 different models used to model the relationship between daily reported Ct value statistics and epidemic growth rates, fitted to the realistic baseline synthetic dataset. We tested various combinations of three Ct-value statistics (mean, standard deviation and skewness), incorporation of a variant 'era' interaction term (intercept only or intercept and coefficient), and different functional forms of the model (log-linear regression or cubic regression splines).

| | | | RMSE | | | AUC | | |
|---|---|---|---|---|---|---|---|---|
| **Spline** | **Model** | **AIC** | **In-s.** | **Now.** | **Inf.** | **In-s.** | **Now.** | **Inf.** |
| None | Mean (no variant era) | -4151 | 0.034 | 0.0349 | 0.0434 | 0.743 | 0.701 | 0.66 |
| None | Mean + variant era | -4175 | 0.0335 | 0.0368 | 0.0473 | 0.752 | 0.662 | 0.608 |
| None | Mean * variant era | -4230 | 0.0326 | 0.0374 | 0.0509 | 0.763 | 0.697 | 0.575 |
| None | Mean + st.dev. (no variant era) | -4159 | 0.0338 | 0.0354 | 0.0442 | 0.75 | 0.696 | 0.651 |
| None | Mean + st.dev. + variant era | -4186 | 0.0333 | 0.0373 | 0.0481 | 0.763 | 0.661 | 0.621 |
| None | (Mean + st.dev.) * variant era | -4256 | 0.032 | 0.0385 | 0.0527 | 0.78 | 0.689 | 0.582 |
| None | Mean + skew (no variant era) | -4149 | 0.034 | 0.0351 | 0.0434 | 0.743 | 0.695 | 0.66 |
| None | Mean + skew + variant era | -4174 | 0.0335 | 0.037 | 0.0474 | 0.752 | 0.654 | 0.609 |
| None | (Mean + skew) * variant era | -4227 | 0.0325 | 0.0386 | 0.0524 | 0.764 | 0.685 | 0.576 |
| None | Mean + s.d. + skew (no variant era) | -4157 | 0.0338 | 0.0358 | 0.0446 | 0.751 | 0.689 | 0.654 |
| None | Mean + s.d. + skew + variant era | -4184 | 0.0333 | 0.0376 | 0.0485 | 0.763 | 0.654 | 0.621 |
| None | (Mean + s.d. + skew) * variant era | -4255 | 0.0319 | 0.0401 | 0.0536 | 0.783 | 0.68 | 0.586 |
| Cubic | Mean (no variant era) | -4229 | 0.0326 | 0.0356 | 0.0466 | 0.755 | 0.715 | 0.668 |
| Cubic | Mean + variant era | -4258 | 0.0321 | 0.0406 | 0.0631 | 0.78 | 0.706 | 0.624 |
| Cubic | Mean * variant era | -4296 | 0.0313 | 0.565 | 1.33 | 0.796 | 0.71 | 0.591 |
| Cubic | Mean + st.dev. (no variant era) | -4238 | 0.0324 | 0.036 | 0.0476 | 0.763 | 0.709 | 0.662 |
| Cubic | Mean + st.dev. + variant era | -4272 | 0.0318 | 0.0407 | 0.0631 | 0.789 | 0.705 | 0.629 |
| Cubic | (Mean + st.dev.) * variant era | -4344 | 0.0304 | 0.564 | 1.33 | 0.811 | 0.694 | 0.636 |
| Cubic | Mean + skew (no variant era) | -4232 | 0.0325 | 0.0366 | 0.0479 | 0.755 | 0.706 | 0.664 |
| Cubic | Mean + skew + variant era | -4261 | 0.0319 | 0.0413 | 0.0636 | 0.78 | 0.7 | 0.622 |
| Cubic | (Mean + skew) * variant era | -4339 | 0.0303 | 0.567 | 1.33 | 0.8 | 0.686 | 0.583 |
| Cubic | Mean + s.d. + skew (no variant era) | -4240 | 0.0323 | 0.0371 | 0.0491 | 0.762 | 0.705 | 0.67 |
| Cubic | Mean + s.d. + skew + variant era | -4274 | 0.0317 | 0.0415 | 0.0639 | 0.789 | 0.699 | 0.628 |
| Cubic | (Mean + s.d. + skew) * variant era | -4379 | 0.0296 | 0.0543 | 0.0981 | 0.814 | 0.676 | 0.643 |
| Key: In-s. = in-sample; Now. = nowcast; Inf. = inflection point | | | | | | | | |

**Table S3.** Summary characteristics of the SARS-CoV-2 testing datasets.

| | **MGB** | **LAC** | **Tufts** |
|---|---|---|---|
| Sample size | 2,671,041 total<br>161,273 positive<br>104,534 included | 330,034 positive<br>279,463 included | 84,848 total<br>10,338 positive<br>10,214 included |
| Dates | Mar 2020-Jan 2023 (1022 days) | May 2020-Jul 2021, Jan-Sep 2022 (680 days) | Feb 2021-Oct 2022 (496 days) |
| Testing modality | Hospital outpatient, inpatient, ER | Voluntary outpatient testing | Hospital outpatient, inpatient, ER |
| Platforms/assays | 7 platforms: Broad in-house assay; Cepheid SARS-CoV-2; Cepheid multiplex SARS-CoV-2/influenza/RSV; Hologic Fusion; Roche Cobas SARS-CoV-2; Roche Cobas multiplex SARS-CoV-2/influenza; Roche Liat multiplex SARS-CoV-2/influenza (see Figure S12) | Fulgent Genetics platform using ThermoFisher QuantStudio™ 6 and 7 PCR system, with LOINC 94531-1 (primarily to Nov 2020) and LOINC 94533-7 (primarily after Nov 2020) | Alinity single-plex and Alinity multiplex |
| Symptom status known? | No | Yes (approx. 55% symptomatic from Sep 2020 onward) | Yes (approx. 65% symptomatic) |
| Vaccination status known? | No | Yes (approx. 25% of all included results; >70% of 2022 results) | No |

| Dataset | AIC | BIC | RMSE | | | | Spearman's Rho | | | | 95% PrI coverage | | | | AUC | | | |
|---|---|---|---|---|---|---|---|---|---|---|---|---|---|---|---|---|---|---|
| | | | **In-s.** | **Now.** | **Inf.** | **FS** | **In-s.** | **Now.** | **Inf.** | **FS** | **In-s.** | **Now.** | **Inf.** | **FS** | **In-s.** | **Now.** | **Inf.** | **FS** |
| MGB | -3493 | -3433 | 0.0451 | 0.0523 | 0.0645 | 0.047 | 0.523 | 0.398 | 0.333 | 0.44 | 0.949 | 0.929 | 0.873 | 0.938 | 0.785 | 0.723 | 0.722 | 0.754 |
| LAC | -2660 | -2585 | 0.0335 | 0.039 | 0.0471 | 0.0458 | 0.649 | 0.556 | 0.394 | 0.573 | 0.953 | 0.912 | 0.837 | 0.888 | 0.843 | 0.784 | 0.772 | 0.724 |
| TFT | -1691 | -1649 | 0.0415 | 0.0497 | 0.0591 | 0.0695 | 0.455 | 0.266 | 0.149 | 0.554 | 0.944 | 0.864 | 0.791 | 0.801 | 0.754 | 0.685 | 0.584 | 0.796 |
| Key: In-s. = in-sample; Now. = nowcast; Inf. = inflection point; FS = fixed train-test split | | | | | | | | | | | | | | | | | | |

**Table S4.** Summary of the performance of the chosen model in predicting epidemic growth rates using Ct values for MGB, LAC, and Tufts datasets, including in-sample fits, nowcast performance, inflection period performance, and fit over the testing period with a single fixed train-test split. Metrics reported are RMSE of predicted vs. observed log incidence growth rates, Spearman's rank-order correlation coefficient for predicted vs. observed growth rates, proportion of observed growth rates falling within the 95% prediction interval, and AUC for epidemic direction predictions.

**Table S5.** Summary of model performance metrics for the downsampled MGB and external comparison (Tufts) datasets, for nowcast performance and comparable baseline nowcast performance. Metrics reported are RMSE of predicted vs. observed log incidence growth rates, Spearman's rank-order correlation coefficient for predicted vs. observed growth rates, proportion of observed growth rates falling within the 95% prediction interval, and AUC for epidemic direction predictions.

| **Dataset** | **Days included** | **RMSE** | | **Spearman's Rho** | | **95% PrI coverage** | | **AUC** | |
|---|---|---|---|---|---|---|---|---|---|
| | | **Now.** | **Comp.** | **Now.** | **Comp.** | **Now.** | **Comp.** | **Now.** | **Comp.** |
| Tufts | 413 | 0.0497 | 0.0489 | 0.266 | 0.348 | 0.864 | 0.937 | 0.685 | 0.719 |
| 10% downsample | 582.54 | 0.0564 | 0.0525 | 0.137 | 0.321 | 0.927 | 0.932 | 0.615 | 0.714 |
| 25% downsample | 860.02 | 0.0494 | 0.0492 | 0.323 | 0.389 | 0.939 | 0.942 | 0.698 | 0.729 |
| 50% downsample | 909.84 | 0.0501 | 0.0516 | 0.373 | 0.4 | 0.936 | 0.936 | 0.718 | 0.729 |
| 75% downsample | 927.64 | 0.0506 | 0.0518 | 0.399 | 0.41 | 0.935 | 0.935 | 0.736 | 0.731 |
| 25/day max samples | 944 | 0.0525 | 0.0523 | 0.39 | 0.398 | 0.93 | 0.931 | 0.721 | 0.723 |
| 50/day max samples | 944 | 0.0502 | 0.0523 | 0.375 | 0.398 | 0.935 | 0.931 | 0.714 | 0.723 |
| 100/day max samples | 944 | 0.0515 | 0.0523 | 0.384 | 0.398 | 0.932 | 0.931 | 0.718 | 0.723 |
| 2.5% trimmed | 930 | 0.0502 | 0.0516 | 0.431 | 0.416 | 0.933 | 0.935 | 0.75 | 0.73 |
| 5% trimmed | 930 | 0.0496 | 0.0516 | 0.436 | 0.416 | 0.938 | 0.935 | 0.748 | 0.73 |
| 10% trimmed | 921 | 0.0496 | 0.0517 | 0.397 | 0.413 | 0.933 | 0.936 | 0.726 | 0.735 |
| Key: Now. = nowcast; Comp. = comparison data | | | | | | | | | |

| | Model | AIC | BIC | RMSE | | | | Spearman's Rho | | | | 95% PrI coverage | | | | AUC | | | |
|---|---|---|---|---|---|---|---|---|---|---|---|---|---|---|---|---|---|---|---|
| | | | | In-s. | Now. | Inf. | FS | In-s. | Now. | Inf. | FS | In-s. | Now. | Inf. | FS | In-s. | Now. | Inf. | FS |
| MGB | Base model | -3493 | -3433 | 0.0451 | 0.0523 | 0.0645 | 0.047 | 0.523 | 0.398 | 0.333 | 0.44 | 0.949 | 0.929 | 0.873 | 0.933 | 0.785 | 0.723 | 0.722 | 0.754 |
| | Outpatient only | -3432 | -3358 | 0.0439 | 0.0494 | 0.0653 | 0.041 | 0.546 | 0.336 | 0.234 | 0.462 | 0.947 | 0.941 | 0.903 | 0.95 | 0.804 | 0.724 | 0.696 | 0.763 |
| LAC | Base model | -2660 | -2585 | 0.0335 | 0.039 | 0.0471 | 0.0458 | 0.649 | 0.556 | 0.394 | 0.573 | 0.954 | 0.912 | 0.837 | 0.884 | 0.843 | 0.784 | 0.772 | 0.724 |
| | Symptom stratified | -2005 | -1914 | 0.0326 | 0.0454 | 0.0507 | 0.0648 | 0.728 | 0.45 | 0.369 | 0.313 | 0.949 | 0.846 | 0.744 | 0.577 | 0.907 | 0.829 | 0.765 | 0.5 |
| | Asymptomatic only | -1986 | -1916 | 0.0335 | 0.0415 | 0.0497 | 0.0487 | 0.693 | 0.55 | 0.399 | 0.535 | 0.932 | 0.866 | 0.8 | 0.856 | 0.88 | 0.809 | 0.784 | 0.5 |
| | Immunologically naive only | -2556 | -2500 | 0.0344 | 0.0401 | 0.0474 | 0.055 | 0.62 | 0.489 | 0.42 | 0.31 | 0.944 | 0.896 | 0.821 | 0.776 | 0.819 | 0.761 | 0.754 | 0.475 |

Key: In-s. = in-sample; Now. = nowcast; Inf. = inflection point; FS = fixed train-test split

**Table S6.** Summary of sensitivity analyses comparing performance of the base model in predicting epidemic growth rates using Ct values for LAC against models using only asymptomatic / unknown symptom status individuals, and using only immunologically naïve (no known vaccination or previous SARS-CoV-2 infection) individuals, including in-sample fits, nowcast performance, inflection period performance, and fit over the testing period with a single fixed train-test split. Metrics reported are RMSE of predicted vs. observed log incidence growth rates, Spearman's rank-order correlation coefficient for predicted vs. observed growth rates, proportion of observed growth rates falling within the 95% prediction interval, and AUC for epidemic direction predictions.

**Table S7**. Viral kinetics model parameters, descriptions and values used in each of the 6 synthetic datasets. Cells are shaded grey where assumed values differ to estimated values. Values in “Baseline scenario” are the maximum posterior probability estimates from fitting the model. Bottom row shows the sampling delay distribution used for each scenario.

| Parameter | Description | Ideal condition | Realistic kinetics | Clustered sampling | Realistic variation | Symmetric kinetics | Baseline scenario (estimated value) |
|---|---|---|---|---|---|---|---|
| $t_p$ | Days to peak viral load | 1.00 days | 2.41 days | 1.00 days | 1.00 days | 7.00 days | 2.41 days |
| $c_p$ | Minimum Ct value | 15.0 | 26.1 | 15.0 | 15.0 | 15.0 | 26.1 |
| $c_s$ | Ct value at inflection point | 29.9 | 29.9 | 29.9 | 29.9 | 29.9 | 29.9 |
| $t_s$ | Days from peak to inflection point | 14.0 days | 5.20 days | 14.0 days | 14.0 days | 5.20 days | 5.20 days |
| $t_c$ | Days from inflection point to full clearance | 50.3 days | 50.3 days | 50.3 days | 50.3 days | 50.3 days | 50.3 days |
| $c_0$ | Baseline Ct value at time of infection | 43.8 | 43.8 | 43.8 | 43.8 | 43.8 | 43.8 |
| $\sigma_{obs}$ | Unmodified variance of observed Ct values for a given time-since-infection | 2.70 | 2.70 | 2.70 | 5.40 | 2.70 | 5.40 |
| $t_m$ | Days to reach minimum variance in observed Ct values | 12.1 days | 5.14 days | 12.1 days | 12.1 days | 5.14 days | 5.14 days |
| $s_m$ | Proportion reduction on variance at minimum (observation variance decreases from $\sigma_{obs}$ to $s_m\sigma_{obs}$) | 0.473 | 0.473 | 0.473 | 0.473 | 0.473 | 0.473 |
| $p_c$ | Daily probability of full clearance | 0.163 | 0.163 | 0.163 | 0.163 | 0.163 | 0.163 |
| Sampling delay distribution | | Uniform(0,7) | Uniform(0,7) | Gamma(1.8,0.6) | Uniform(0,7) | Uniform(0,7) | Gamma(1.8,0.6) |

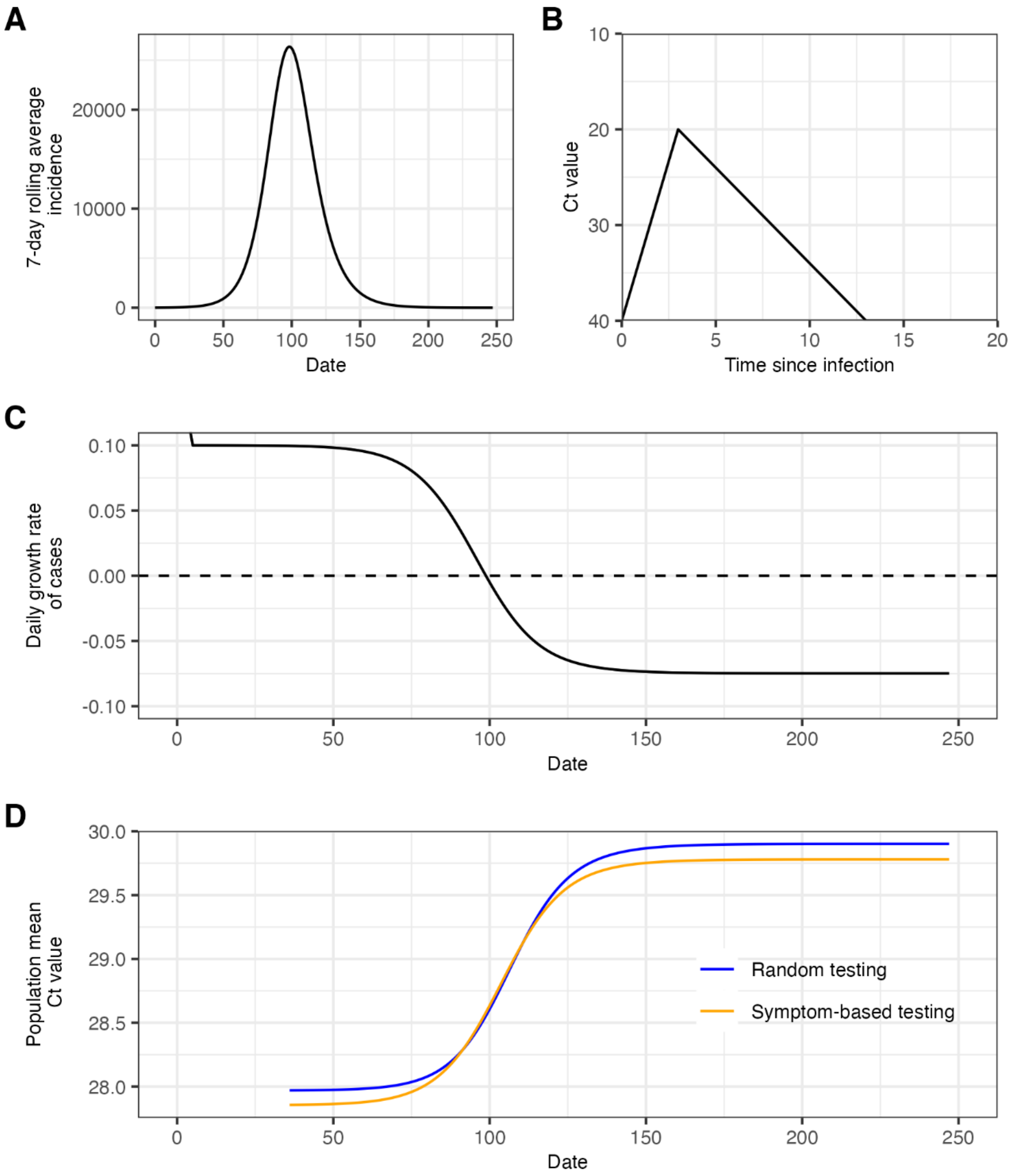


**Figure S1.** Mathematical model describing population-level mean Ct values through the convolution of infection incidence and a within-host viral kinetics model. (**A**) 7-day rolling average infection incidence generated from a deterministic SIR model with $R0$=1.5. (**B**) Viral kinetics model giving the mean Ct value over time-since-infection. (**C**) Daily log growth rate of infections calculated from (A). (**D**) Population-level mean Ct values observed through random and symptom-based testing calculated as the analytical convolution of infection incidence in (A) and viral kinetics curve in (B) (see **Supplementary Text 4, section 5**).

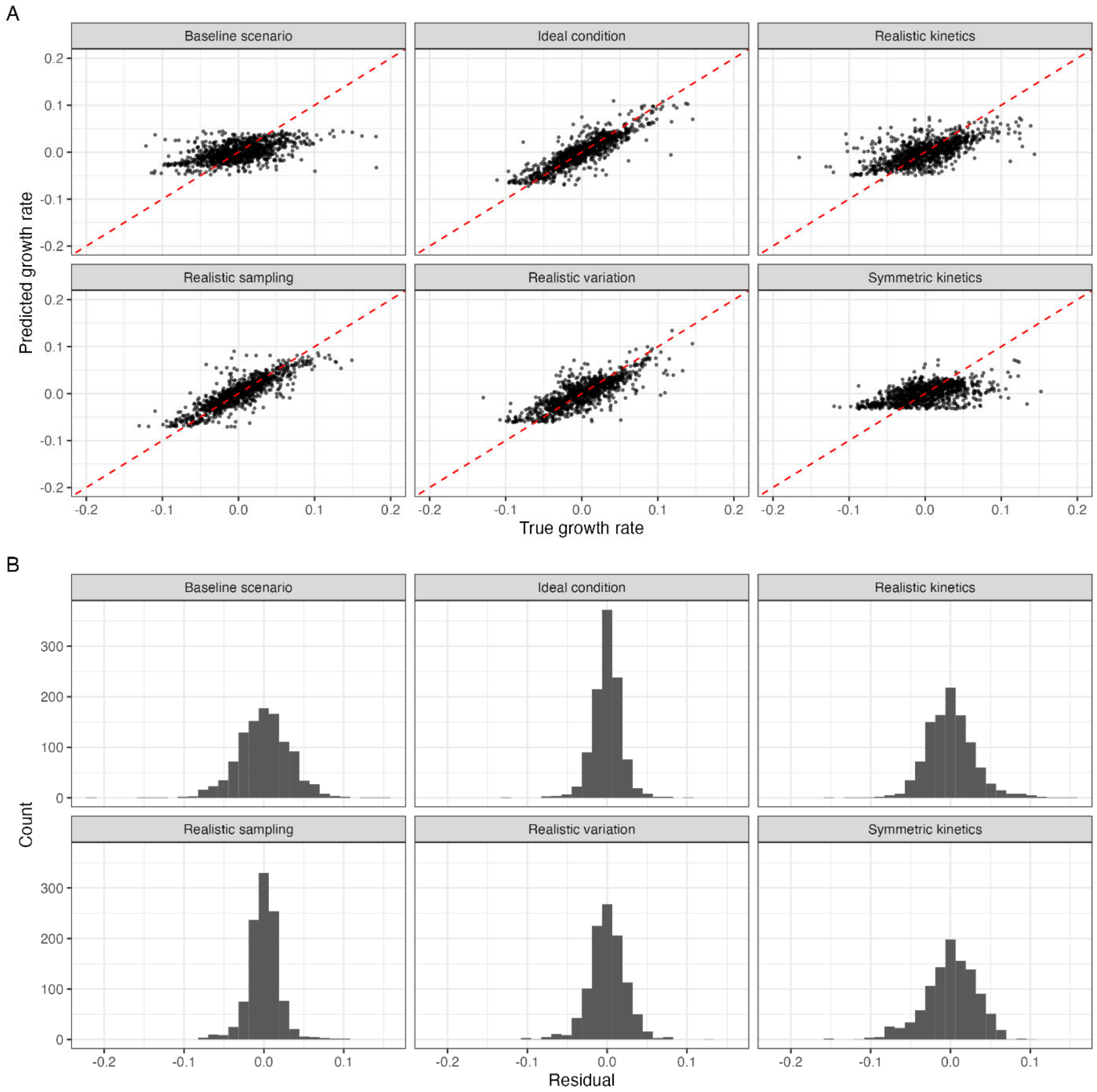


**Figure S2.** (**A**) Comparison of model-predicted incidence growth rates with true growth rates from the synthetic dataset analyses. Estimates shown are from in-sample fits for each scenario using the full time series of data to train the GAM. (**B**) Distribution of residuals between model predictions and true growth rates.

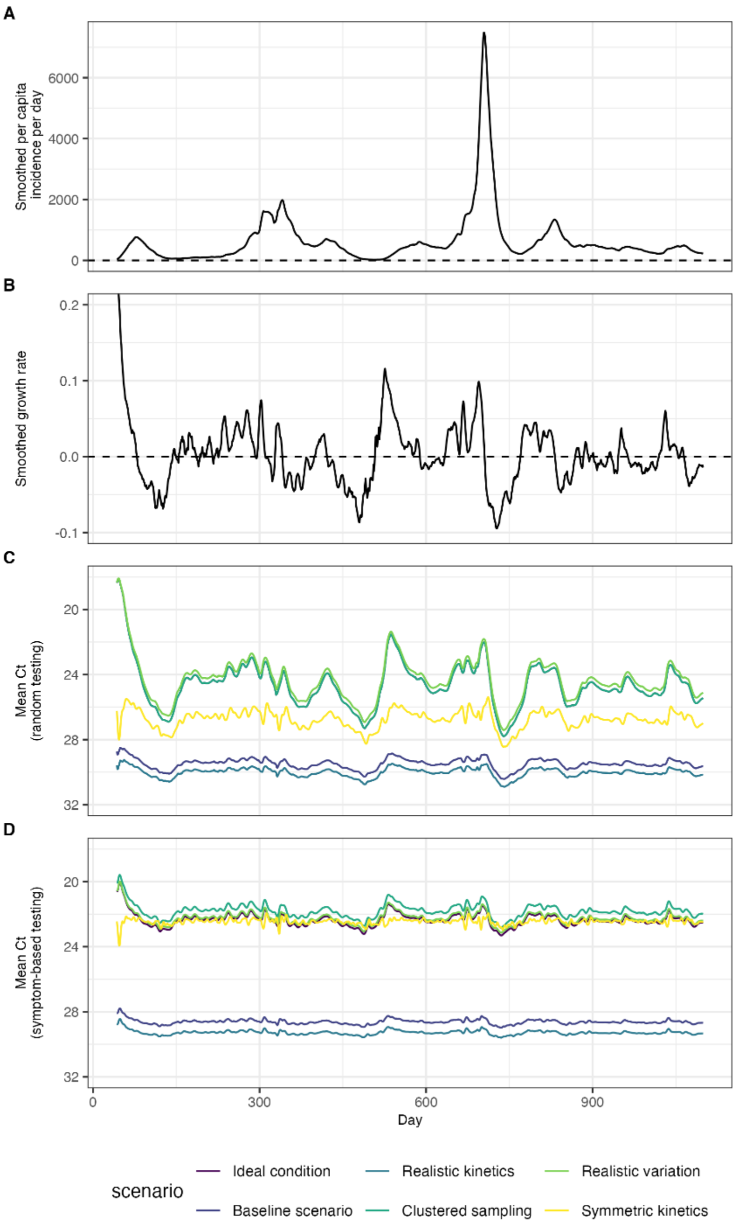


**Figure S3.** Analytical convolutions of an infection incidence curve based on COVID-19 data from Massachusetts, USA, with various model assumptions for within-host viral kinetics and sampling delays. (**A**) 7-day rolling average infection incidence used for the simulation. (**B**) 7-day rolling average growth rates of infection incidence. (**C**) Mean Ct values over time obtained under a random testing scheme based on the analytical convolution described in **Supplementary Text 1, section 5**. Each line represents a different set of model assumptions described in **Table S1.** (**D**) As in (C), but assuming symptom-based testing.

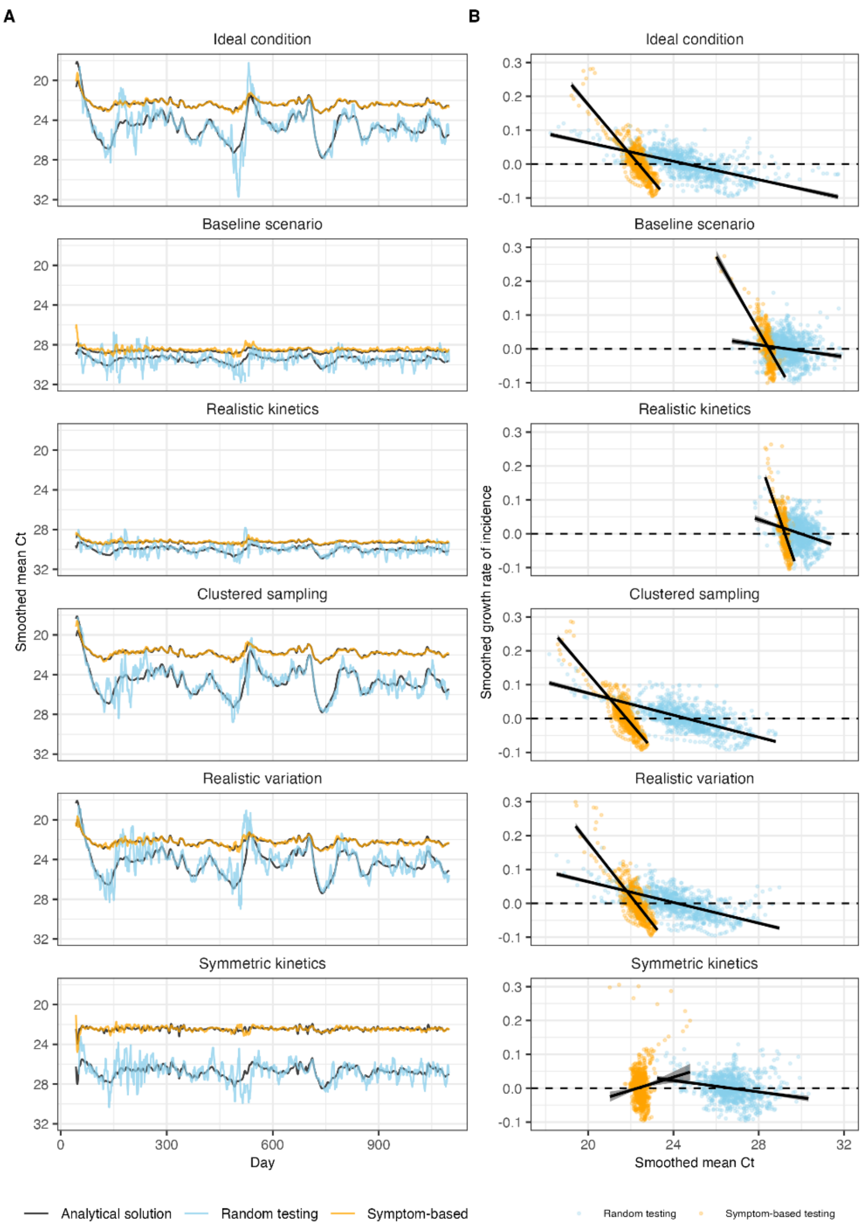


**Figure S4. (A)** Outputs of each synthetic dataset scenario showing the 7-day average mean Ct value reported through the two surveillance strategies. Black lines show the analytically predicted mean Ct value over time based on the convolution of the incidence curve and viral kinetics model, as in **Figure S3**. Colored lines show the outputs from the numerical simulation referenced in the main text. **(B)** Scatterplot showing the relationship between 7-day rolling average growth rates and 7-day rolling average mean Ct value for each scenario, stratified by surveillance strategy. Solid lines show fitted linear regression models.

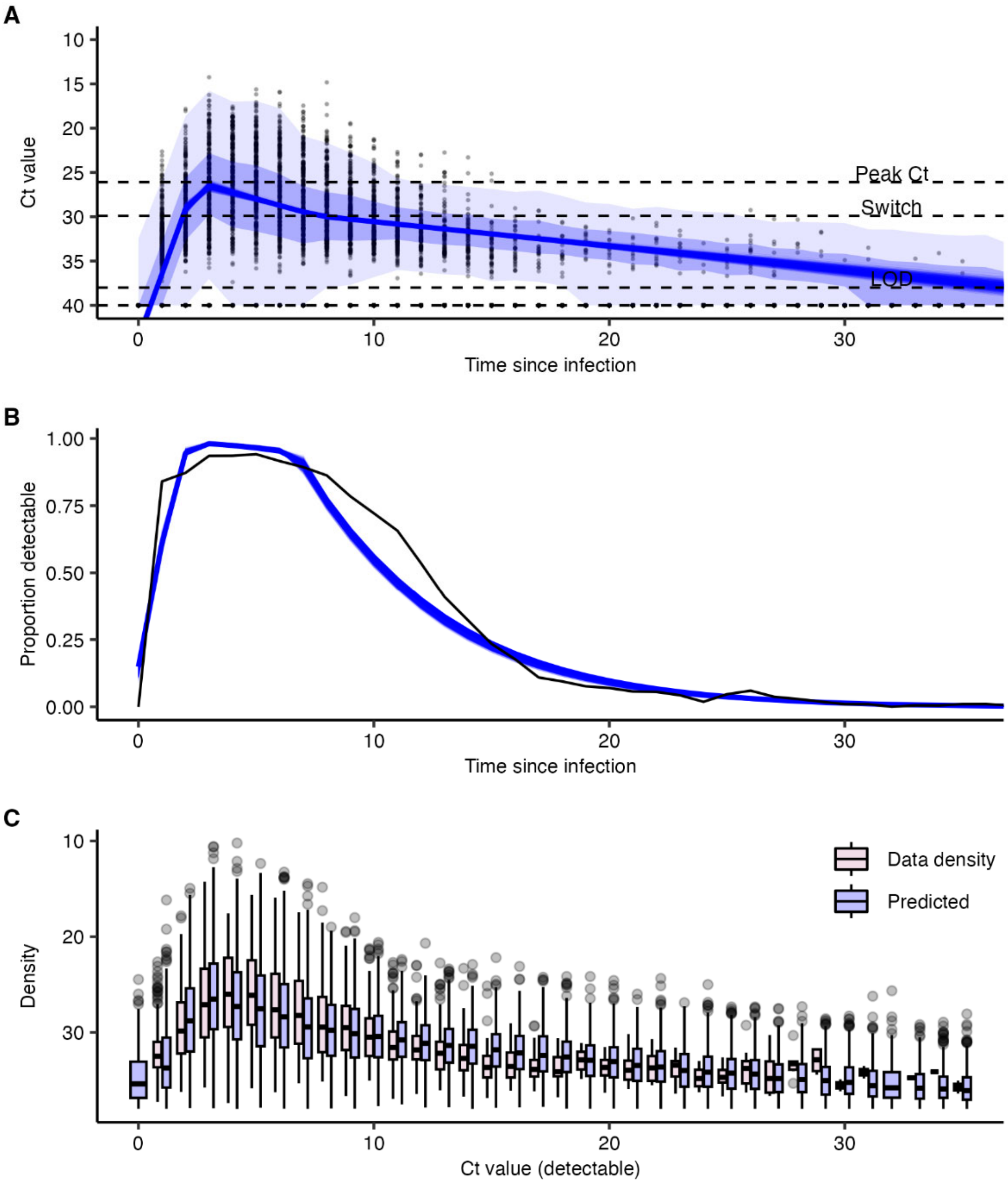


**Figure S5.** Viral kinetics model fitted to longitudinal SARS-CoV-2 RT-qPCR testing data following a previous negative test. **(A)** Solid blue lines show 1000 posterior draws of the mean Ct value over time. Shaded envelope shows 50% and 95% quantiles. Horizontal lines show control points used to parameterize the model. **(B)** Solid blue lines show 1000 posterior draws for the model-predicted proportion of positive tests over time-since infection overlaid on empirical proportion detectable. **(C)** Box plots for the posterior distribution of observed Ct values on each day post infection (blue) compared to distribution of raw data (red).

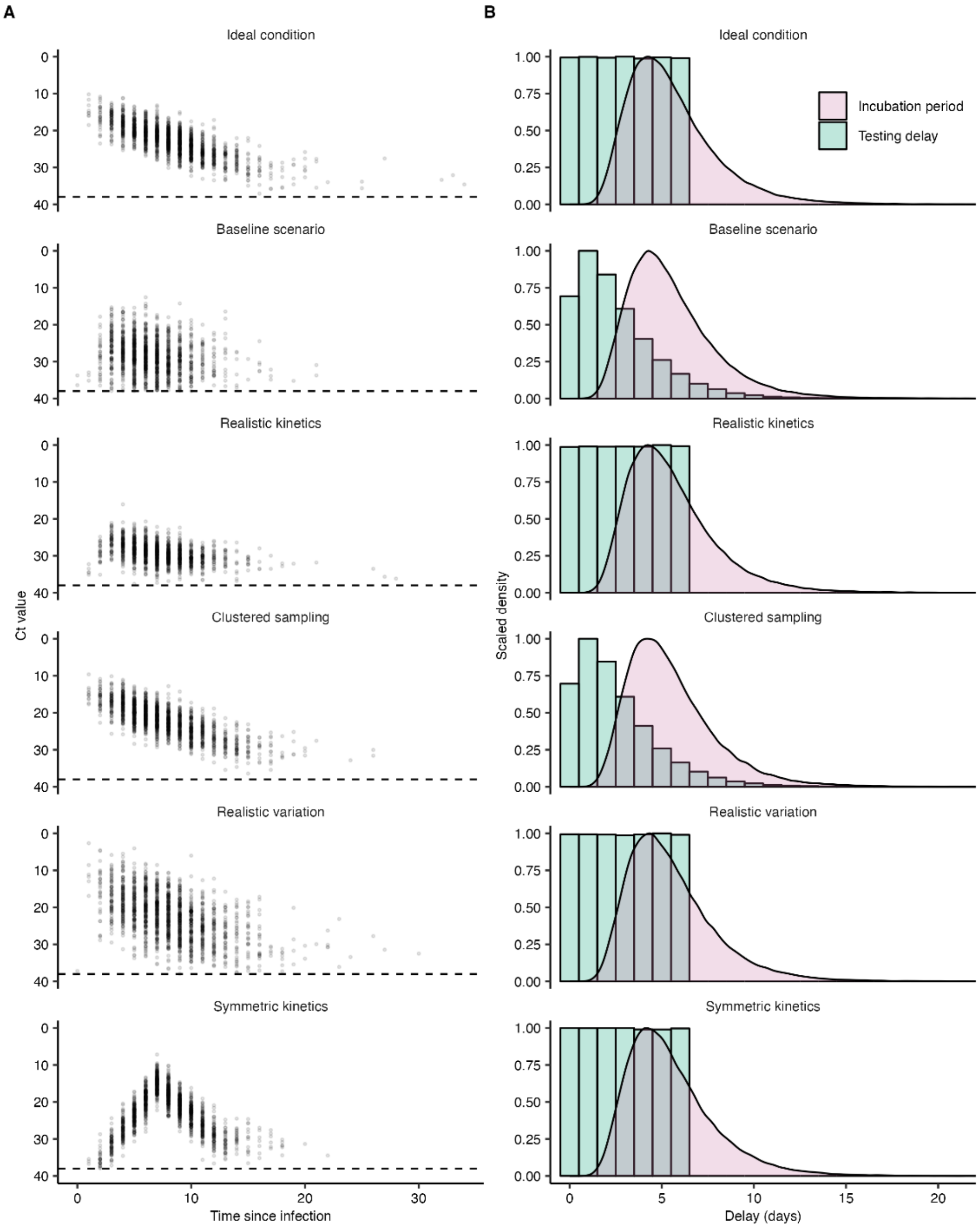


**Figure S6. (A)** Randomly simulated Ct values for the 6 main synthetic data scenarios. "Baseline condition" shows the Ct value distribution as estimated from fitting to the longitudinal testing data, whereas the other scenarios show assumed Ct value distributions after changing some parameters. **(B)** Assumed incubation period distribution (red) and sampling delay distribution (green) for each scenario.

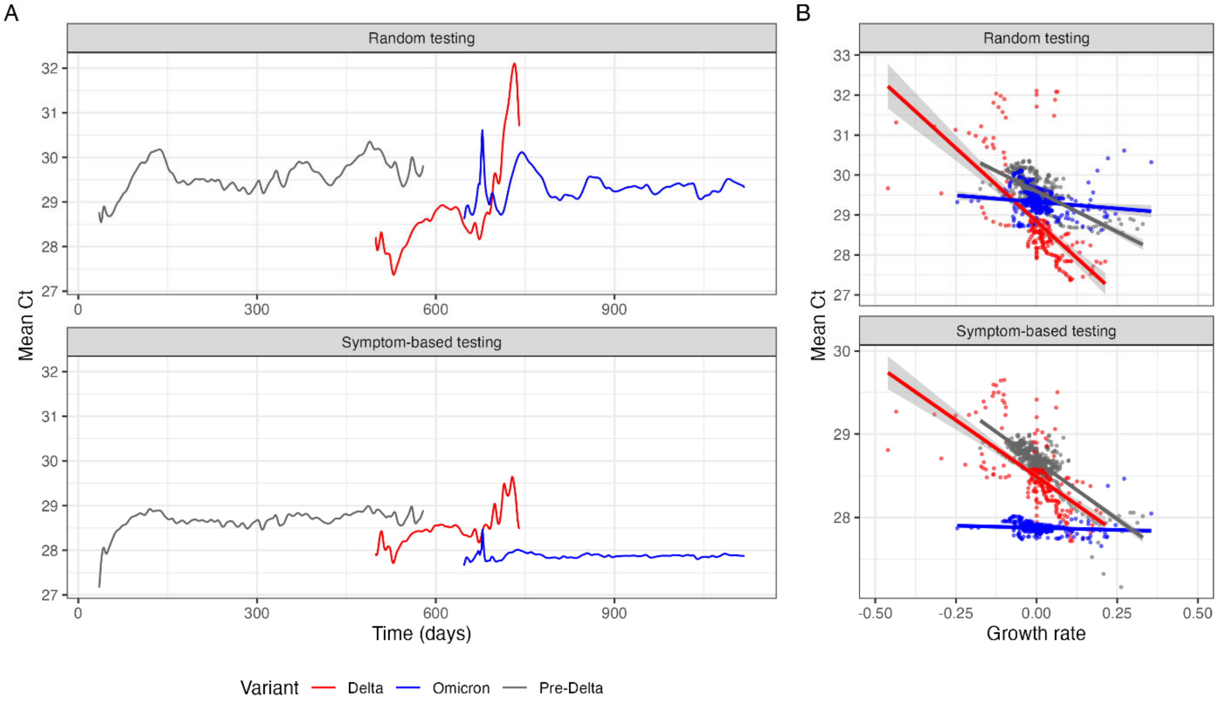


**Figure S7. (A)** Population-level mean Ct values from analytical convolutions of an infection incidence curve based on COVID-19 data from Massachusetts, USA, with within-host viral kinetics models representing different variant waves. (**B**) Scatterplot showing mean Ct value against mean incidence growth rate, stratified by variant. Solid lines and shaded region show fitted linear regression line and 95% confidence intervals. Note that while mean Ct value was negatively correlated with growth rate for all variants, the relationship differed substantially by variant.

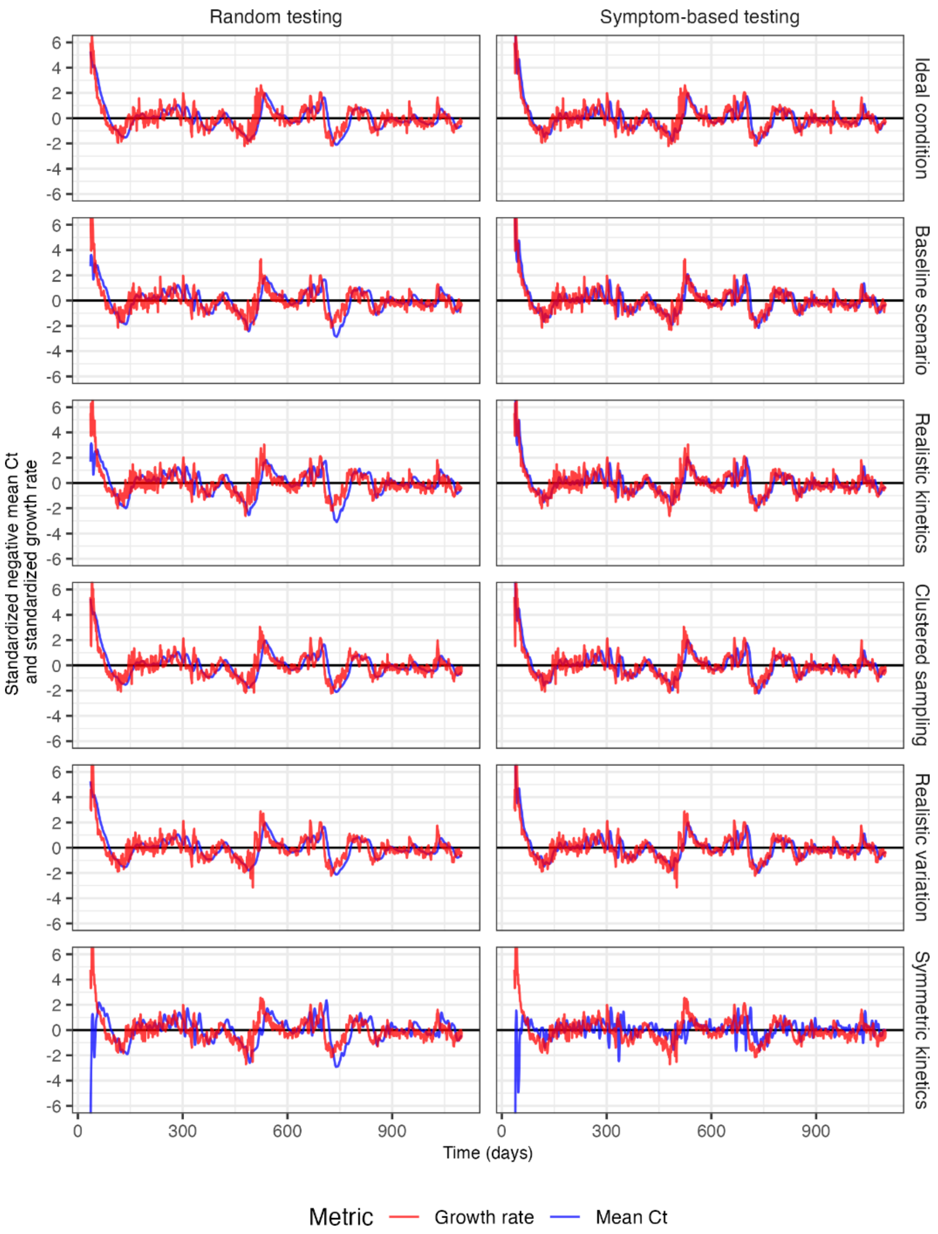


**Figure S8.** Data shown are as in **Figure S3**, but with each dataset standardized by subtracting the mean Ct value from each observation and dividing by the standard deviation.

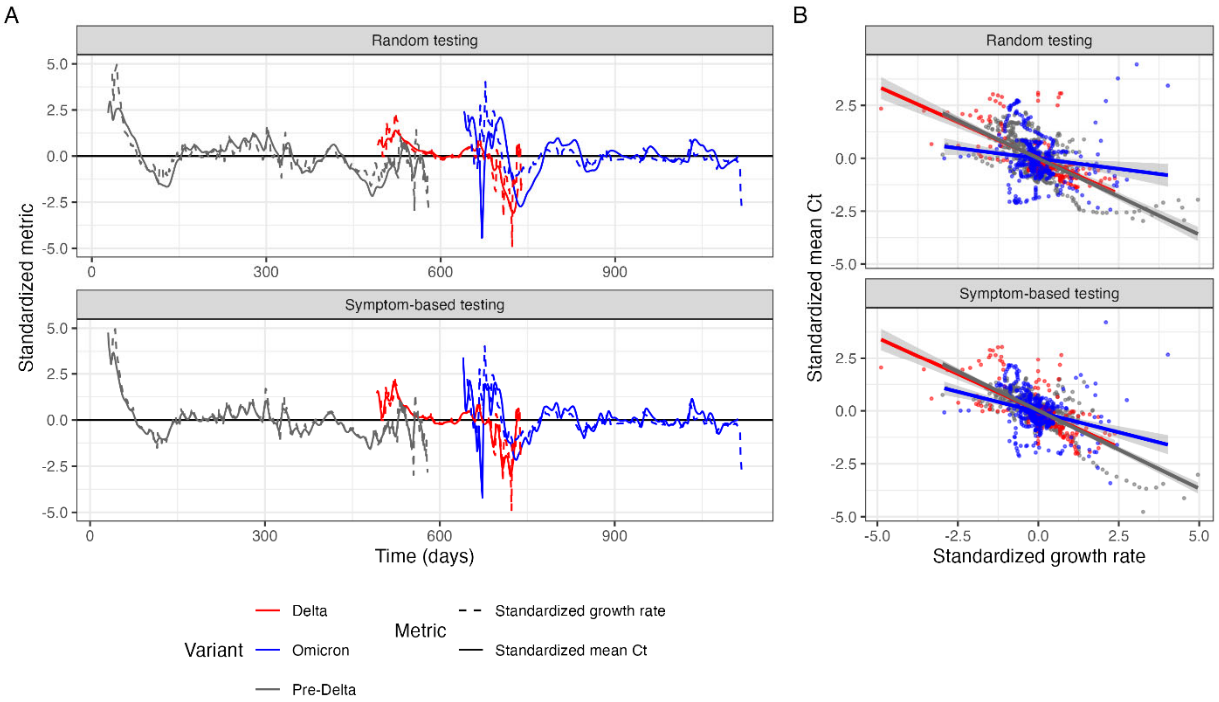


**Figure S9.** As in **Figure S7** but showing Ct values standardized for each variant (subtracting the overall mean Ct value and dividing by the standard deviation). (**A**) now also shows the standardized mean daily growth rate of infections, overlapping almost perfectly with the standardized mean daily Ct value.

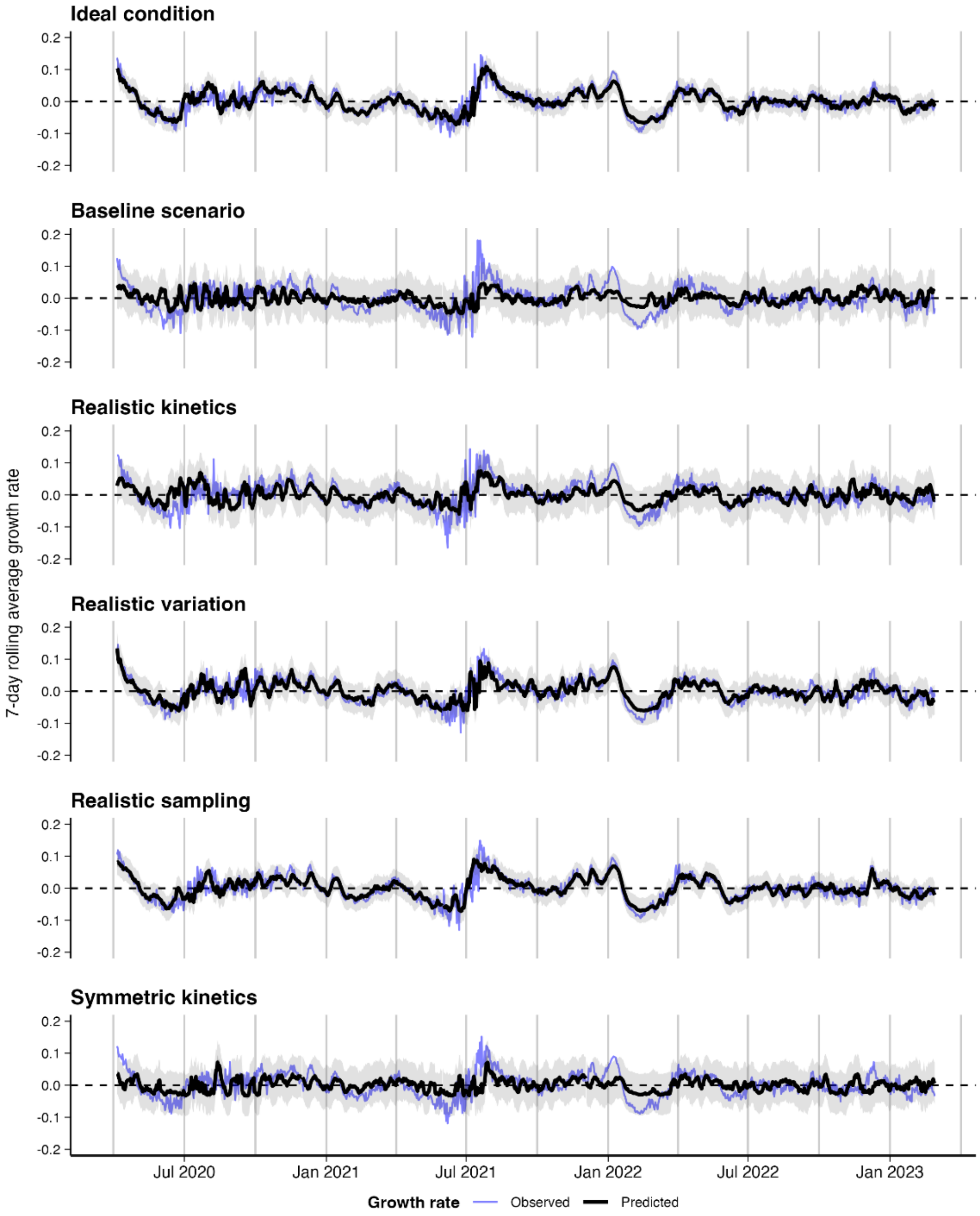


**Figure S10.** In-sample fits of the best-performing GAM model predicting epidemic growth rates over time using only reported Ct values using the 6 synthetic datasets. Blue line shows true growth rate of infections used for the simulation. Black lines and shaded region show model-predicted growth rates and 95% confidence (dark shading) / prediction (light shading) intervals.

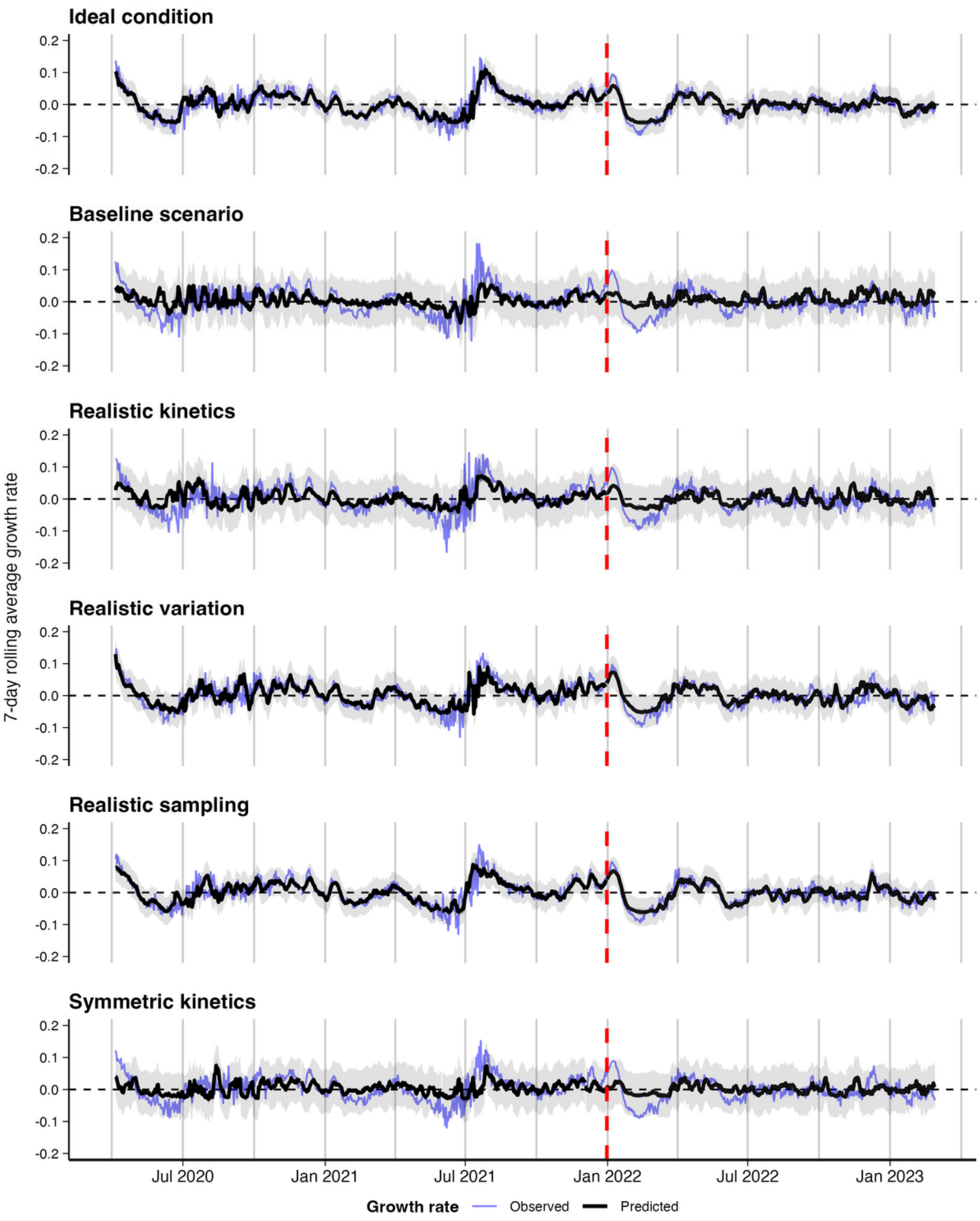


**Figure S11.** Training dataset fits (up to vertical dashed line) and test dataset predicted epidemic growth rates over time using only reported Ct values using the 6 synthetic datasets. Results shown are from the best-performing GAM model. Blue line shows true growth rate of infections used for the simulation. Black lines and shaded region show model-predicted growth rates and 95% confidence (dark shading) / prediction (light shading) intervals.

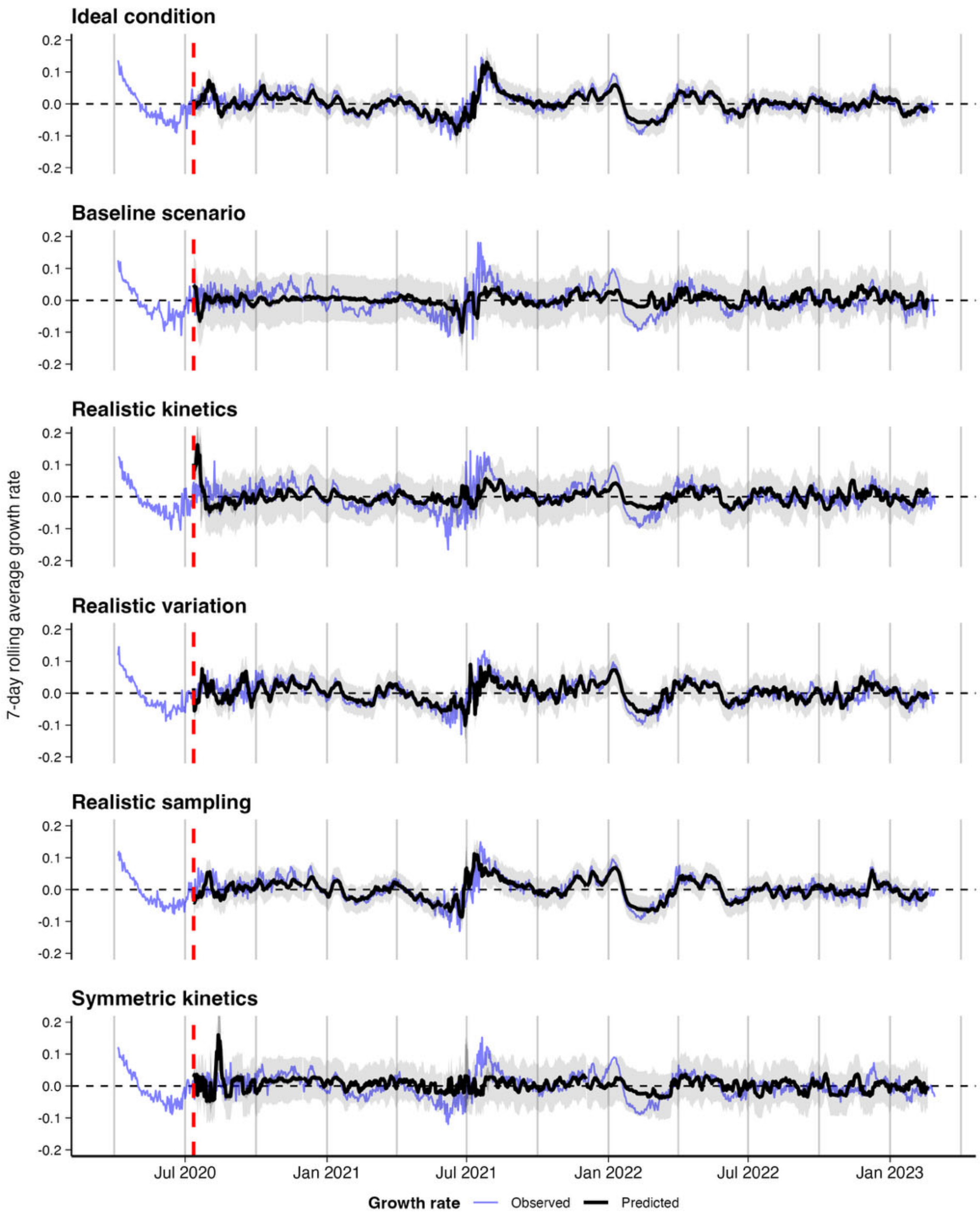


**Figure S12.** Model-predicted (black) vs. observed (blue) log incidence growth rates for the 6 synthetic datasets, with model-predicted 95% confidence intervals (dark shading) and 95% prediction intervals (light shading). Predictions are from 2-week rolling nowcasts, concatenated into a single time series. Vertical dashed line denotes the end of the initial training period.

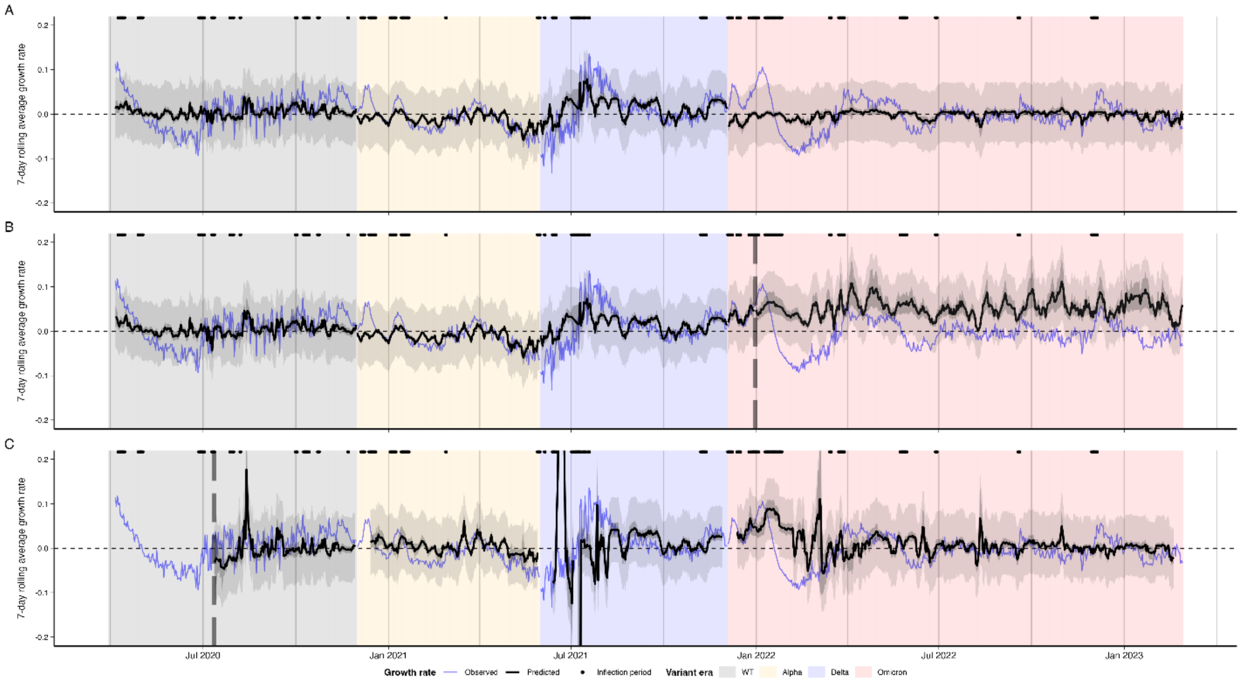


**Figure S13.** Model-predicted vs. observed log incidence growth rate for models fitted using the synthetic, variant-stratified datasets shown in **Figure S7**, showing **(A)** in-sample fits, **(B)** fit over the testing period with a single fixed train-test split shown by the vertical dashed line, and **(C)** two-week rolling nowcast fits, starting at the vertical dashed line. Nowcasting performance was noticeably worse than in-sample predictions, particularly when predictors were stratified by variant, in particular during the early weeks of each variant era.

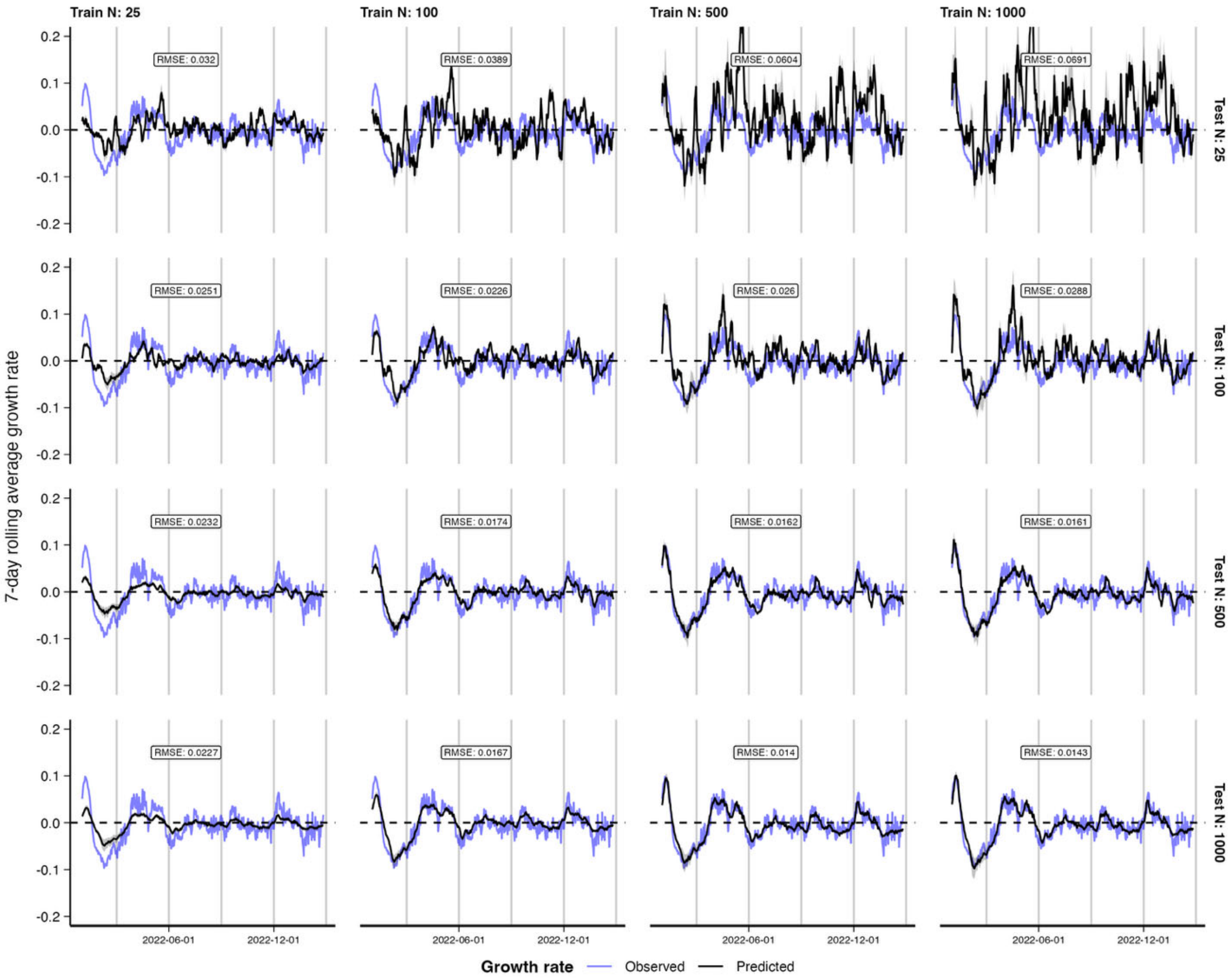


**Figure S14.** Comparison of out-of-sample predicted incidence growth rates from the train/test split analyses against observed growth rates using simulated 7-day rolling average of the mean and skew of Ct values obtained through random testing as a predictor. Data up to and including 2021-12-31 were used to train the model, and data collected thereafter were used for out-of-sample predictions. Columns distinguish different numbers of observations used to train the model. Rows show increasing number of simulated daily observations used to test the model.

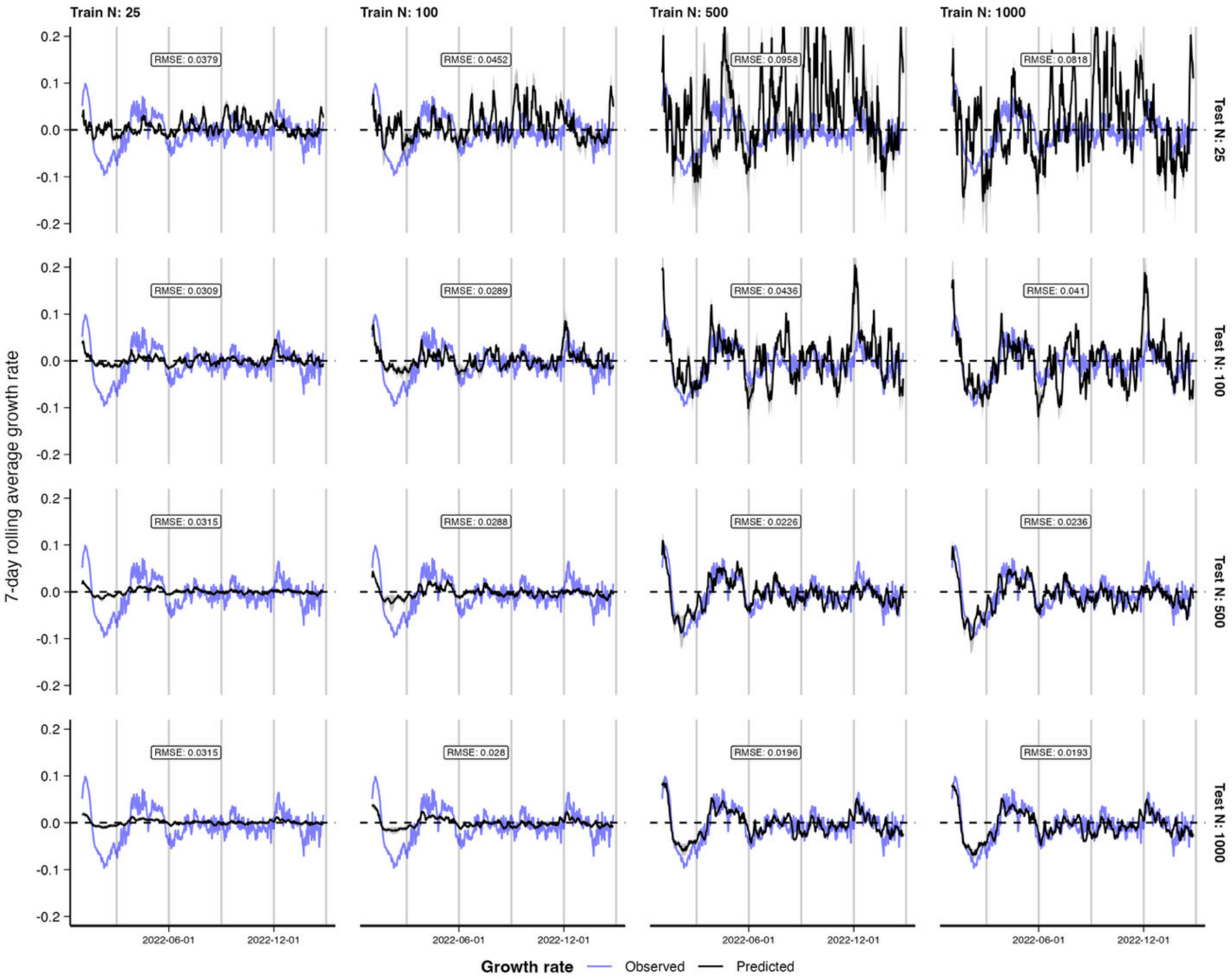


**Figure S15.** Comparison of out-of-sample predicted incidence growth rates from the train/test split analyses against observed growth rates using simulated 7-day rolling average of the mean and skew of Ct values obtained through symptom-based testing as a predictor. Data up to and including 2021-12-31 were used to train the model, and data collected thereafter were used for out-of-sample predictions. Columns distinguish different numbers of observations used to train the model. Rows show increasing number of simulated daily observations used to test the model.

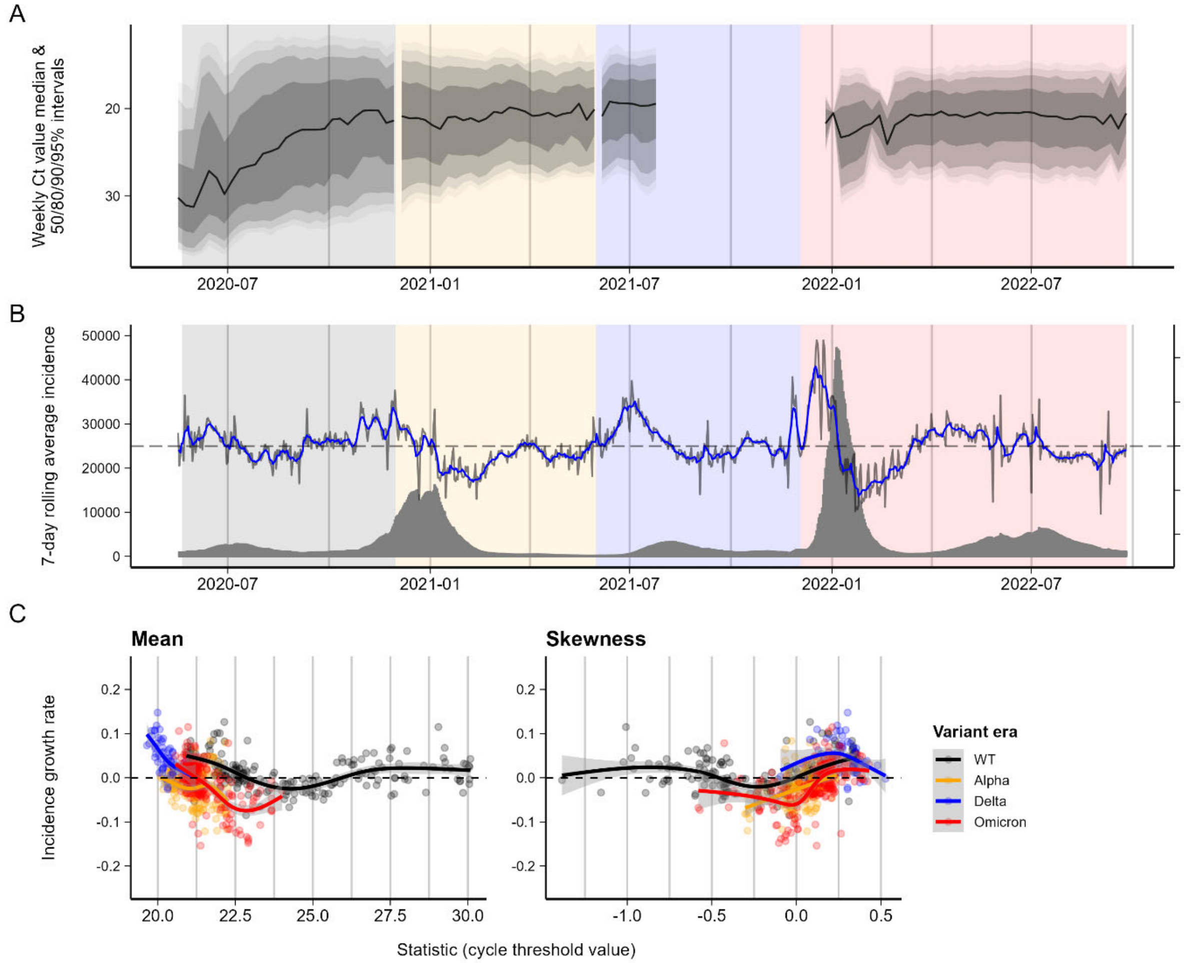


**Figure S16.** Ct values from Los Angeles County and corresponding reported COVID-19 incidence. (**A**) Weekly Ct value quantiles over time, showing weekly median Ct value and 50/80/90/95% quantiles. (**B**) 7-day rolling average reported incidence (grey bars), growth rate in 7-day rolling average reported incidence (grey line), and smoothed growth rate (blue line). Background is shaded by time periods of different variant dominance. Vertical dashed line demarcates the test-train split. (**C**) Incidence growth rate compared to smoothed daily mean and skewness of Ct value distributions. Colored lines and shaded grey regions show fitted cubic spline GAMs with 95% confidence intervals, stratified by period of variant dominance.

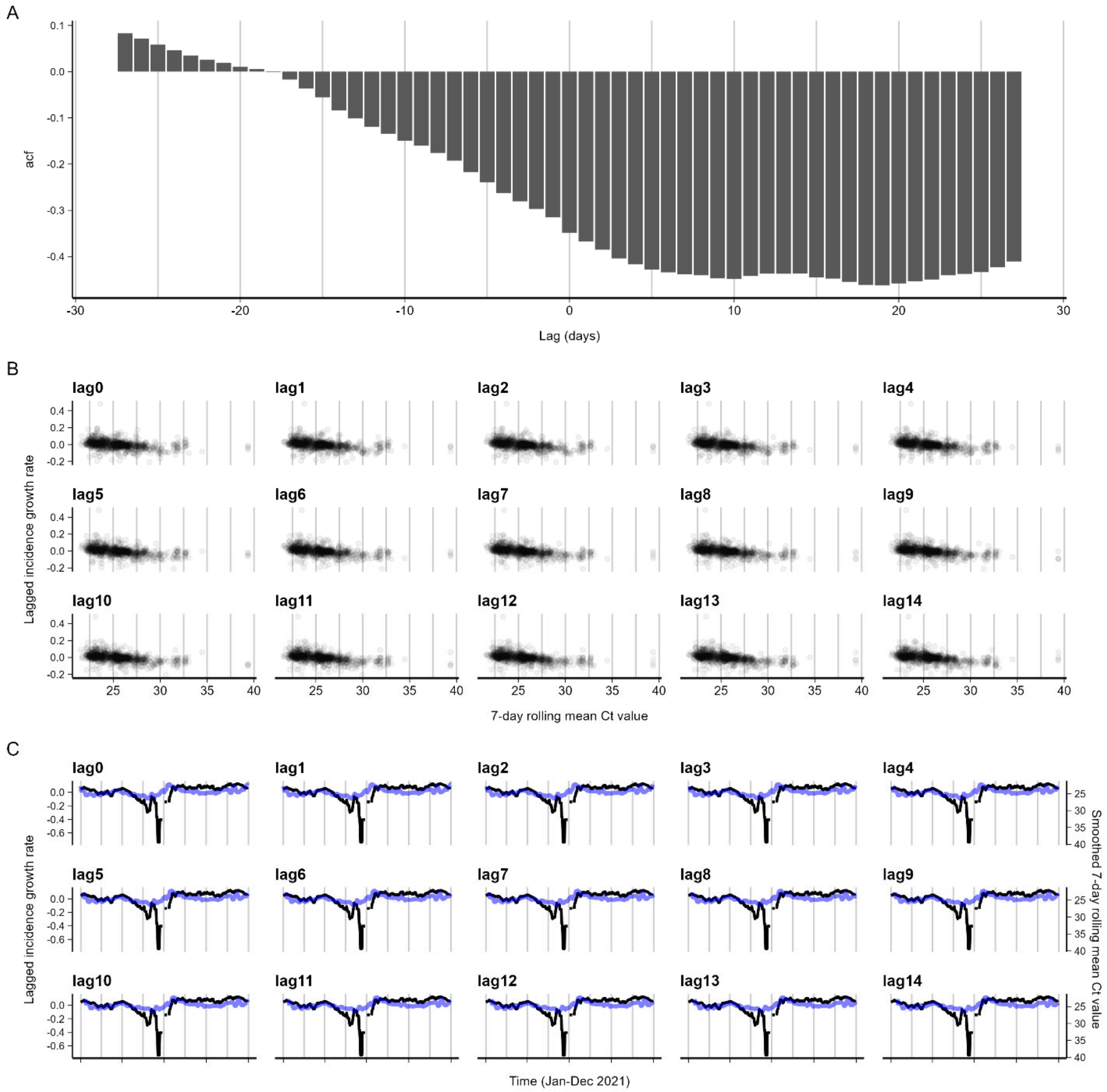


**Figure S17.** Cross-correlations between log incidence growth rates and mean Ct values at different lead/lag times, for MGB data. **(A)** correlation strength, **(B)** scatterplots of daily observed growth rate and mean Ct value at different lags, **(C)** growth rate and mean Ct value over time with different time shifts for the Ct value curve.

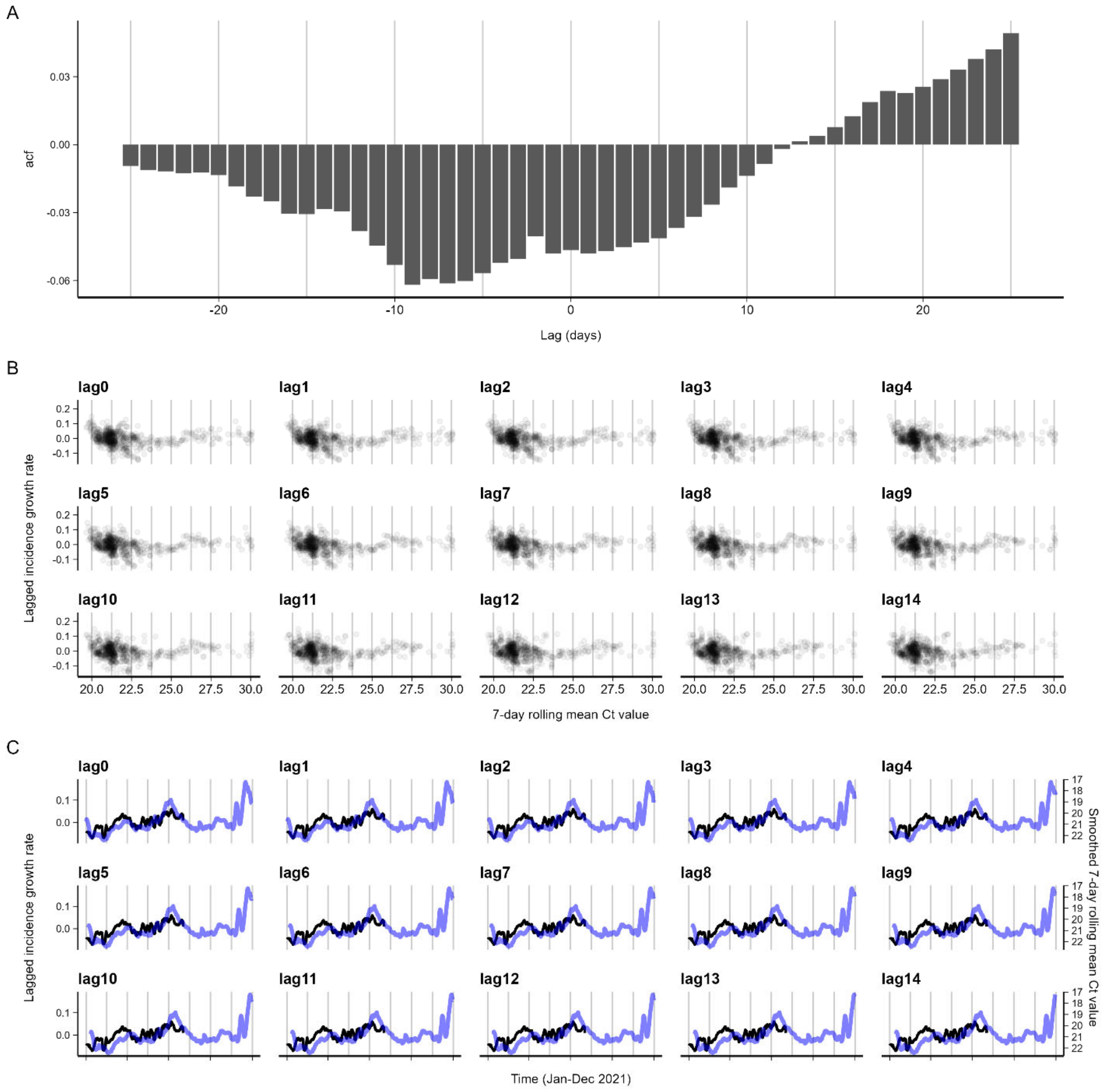


**Figure S18.** Cross-correlations between log incidence growth rates and mean Ct values at different lead/lag times, for LAC data. **(A)** correlation strength, **(B)** scatterplots of daily observed growth rate and mean Ct value, **(C)** growth rate and mean Ct value over time with different time shifts for the Ct value curve.

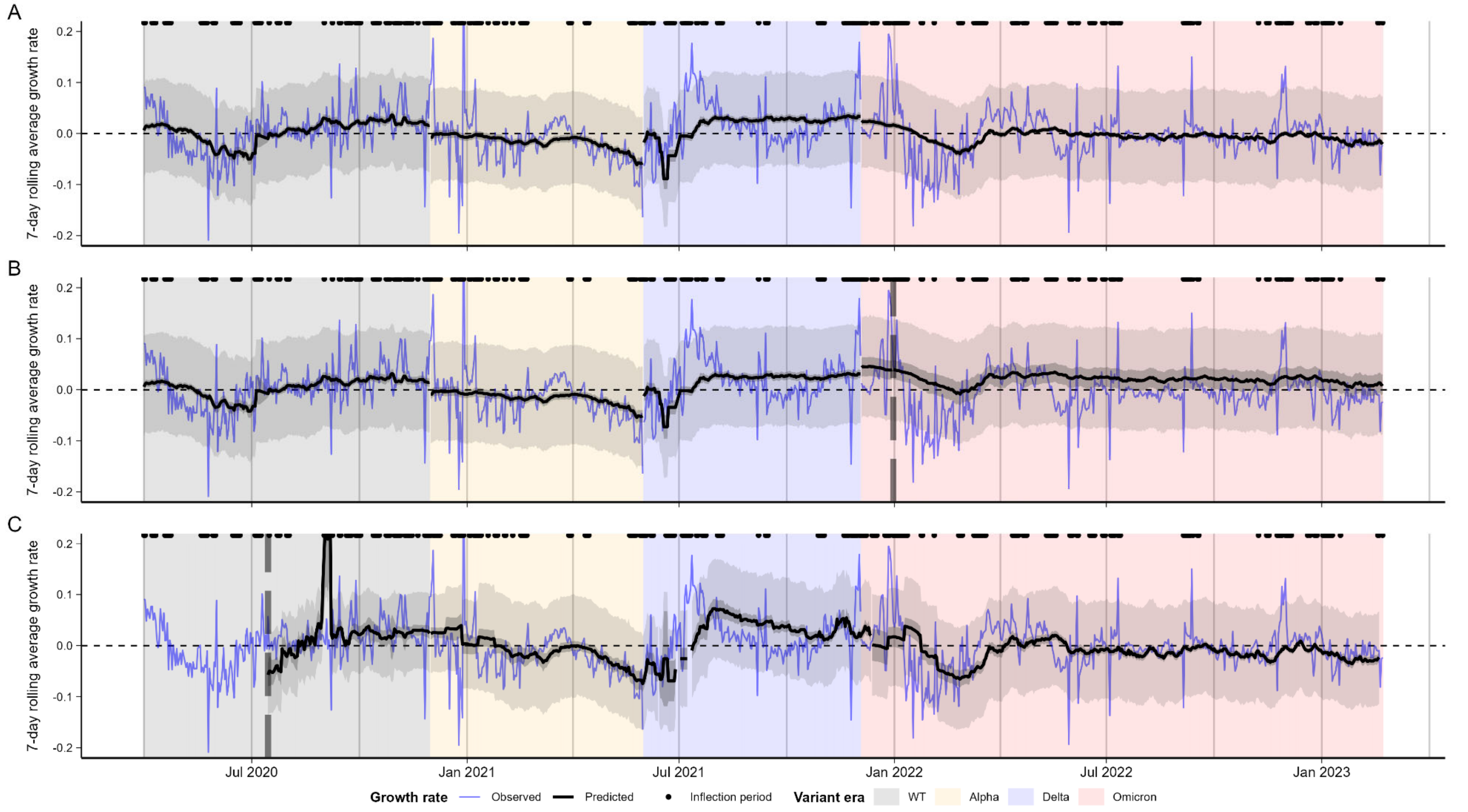


**Figure S19.** Model-predicted vs. observed log incidence growth rate for models fitted using the MGB data, showing **(A)** in-sample fits , **(B)** fit over the testing period with a single fixed train-test split shown by the vertical dashed line, and **(C)** two-week rolling nowcast fits, starting at the vertical dashed line.

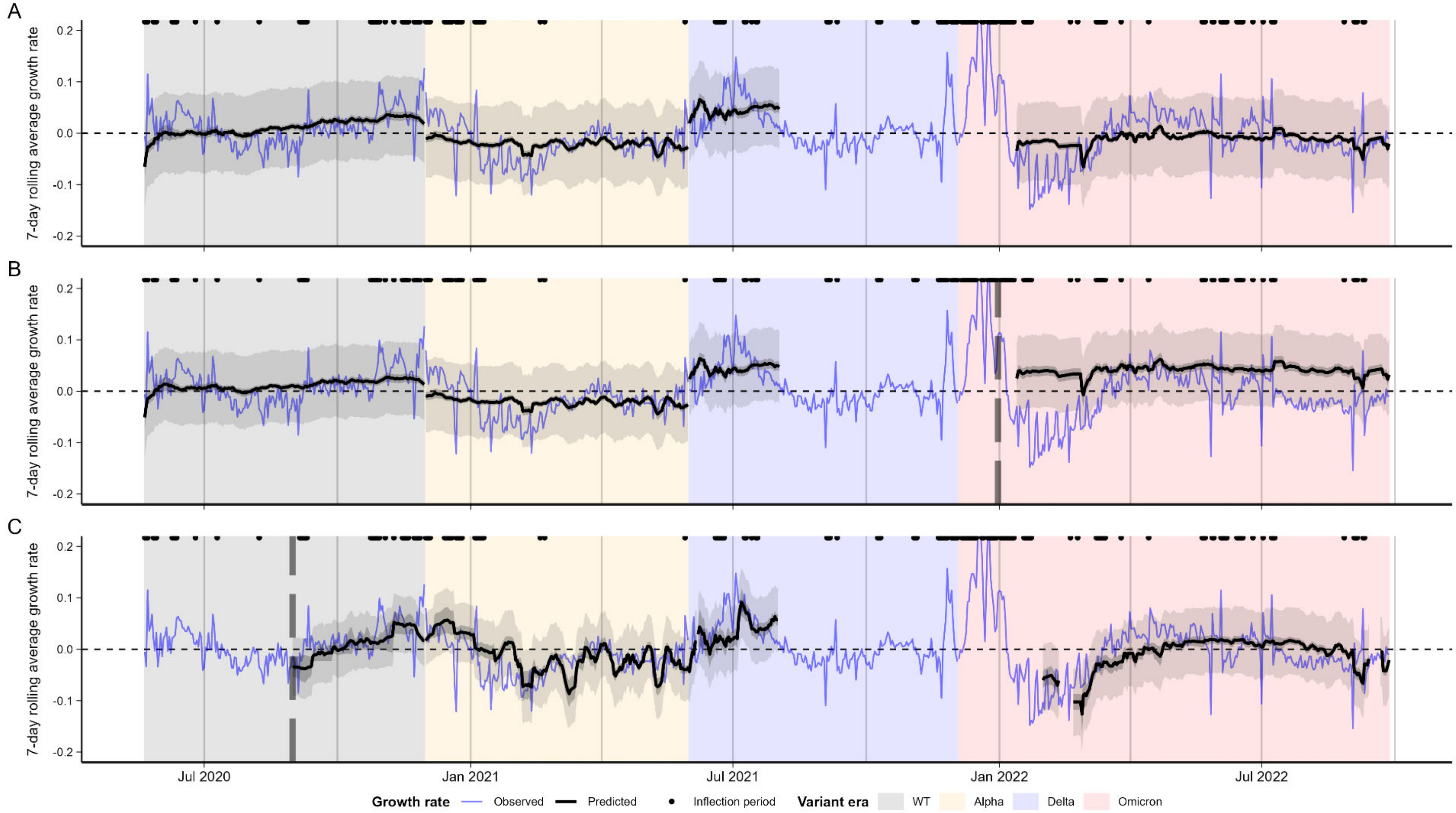


**Figure S20.** Model-predicted vs. observed log incidence growth rate for models fitted using the LAC data, showing **(A)** in-sample fits, **(B)** fit over the testing period with a single fixed train-test split shown by the vertical dashed line, and **(C)** two-week rolling nowcast fits starting at the dashed vertical line.

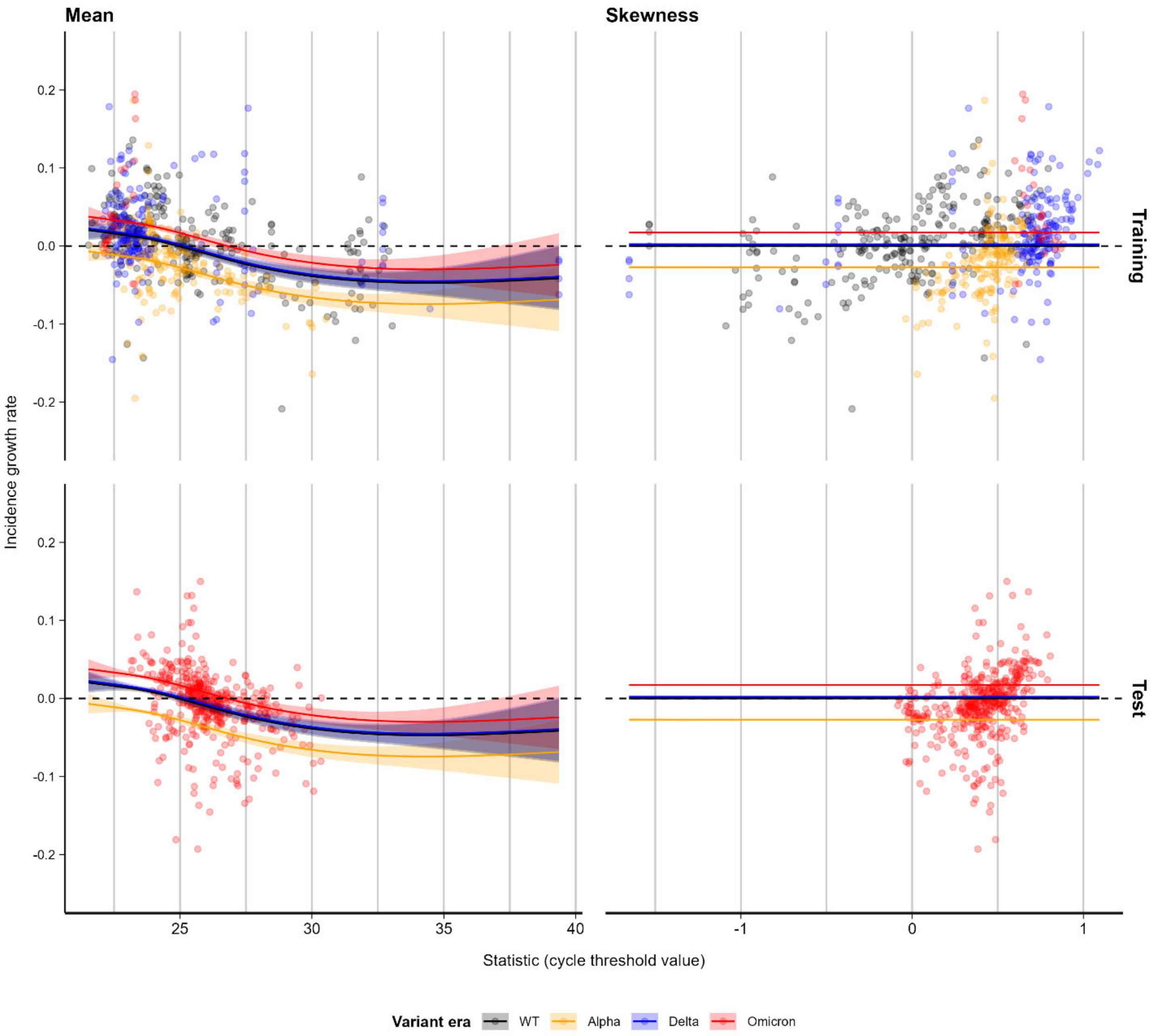


**Figure S21.** Best-fit lines (with standard errors) showing the modeled relationship between mean (left) and skewness (right) in Ct values, by variant era, against growth rate for MGB data. Upper panels show fit to training data with a fixed cutoff at 31 Dec 2021, while lower panels show the fit of the trained model to data in the subsequent test period (01 Jan 2022 onward).

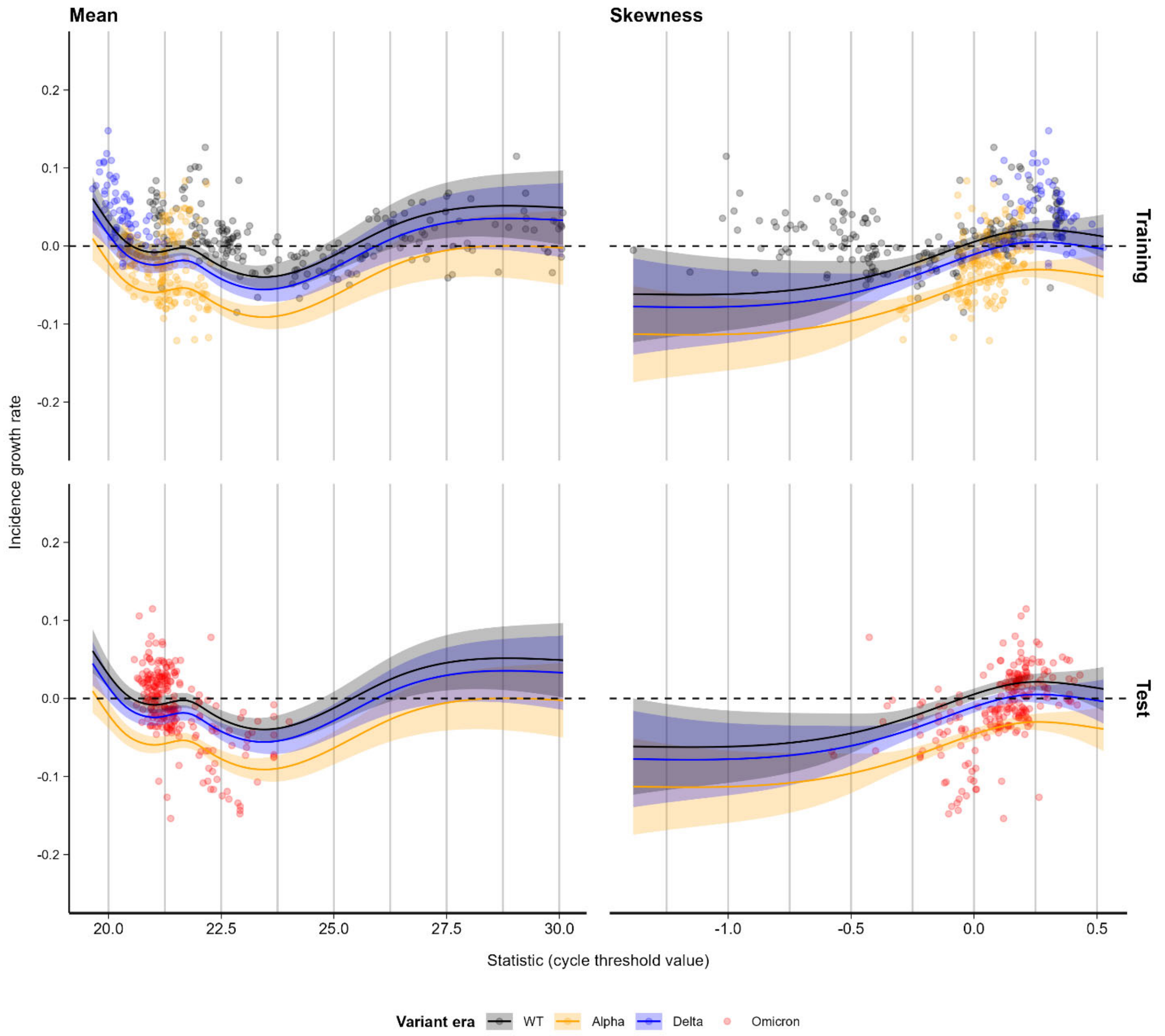


**Figure S22.** Best-fit lines (with standard errors) showing the modeled relationship between mean (left) and skewness (right) in Ct values, by variant era, against growth rate for LAC data. Upper panels show fit to training data with a fixed cutoff at 31 Dec 2021, while lower panels show the fit of the trained model to data in the subsequent test period (01 Jan 2022 onward). Note the absence of Omicron-era data from the training period.

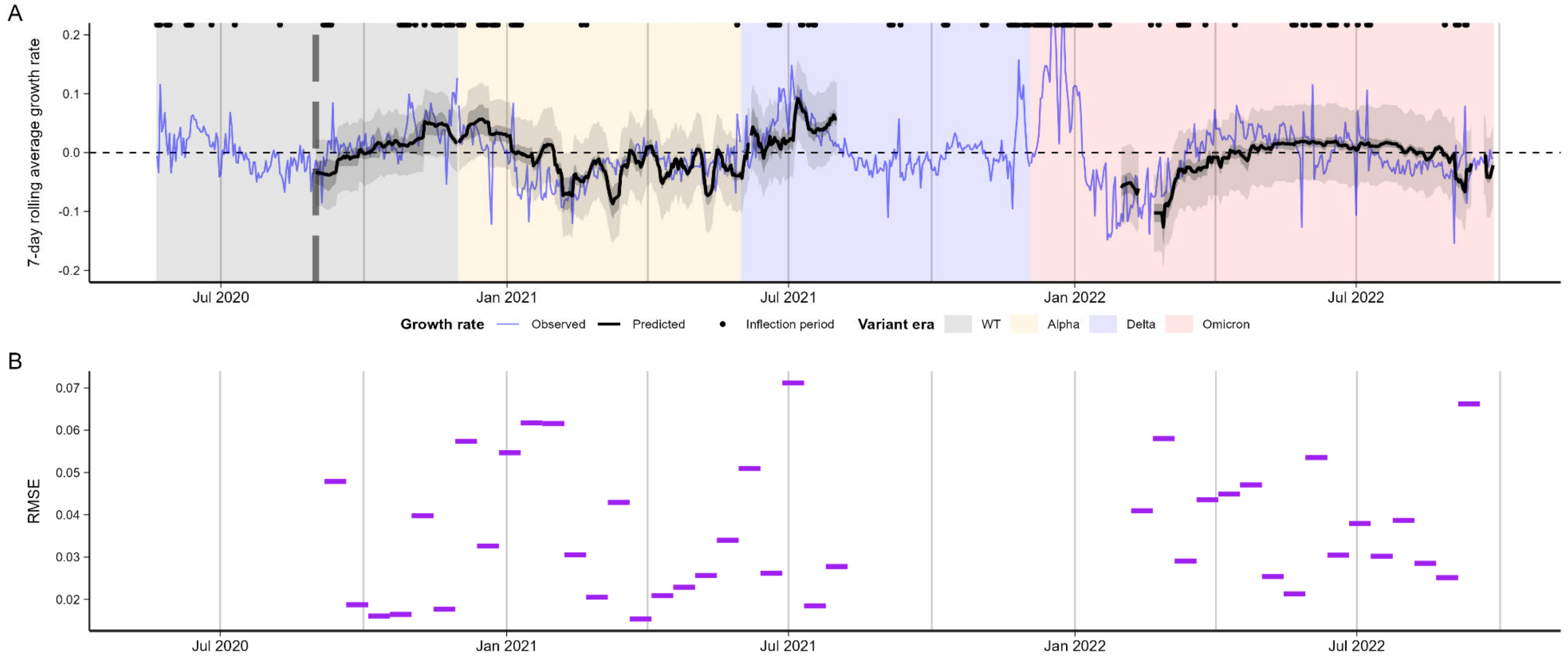


**Figure S23. (A)** Model-predicted vs. observed log incidence growth rates and RMSEs for LAC data. Model-predicted (black) vs. observed (blue) log incidence growth rates for MGB data, with 95% confidence intervals (dark shading) and 95% prediction intervals (light shading). **(B)** RMSE of predicted vs. observed log incidence growth rates for each 2-week nowcasting window. “Inflection periods” refer to times when the absolute smoothed log incidence growth rate exceeded 0.025 over a one-week period, marked with points above each subplot.

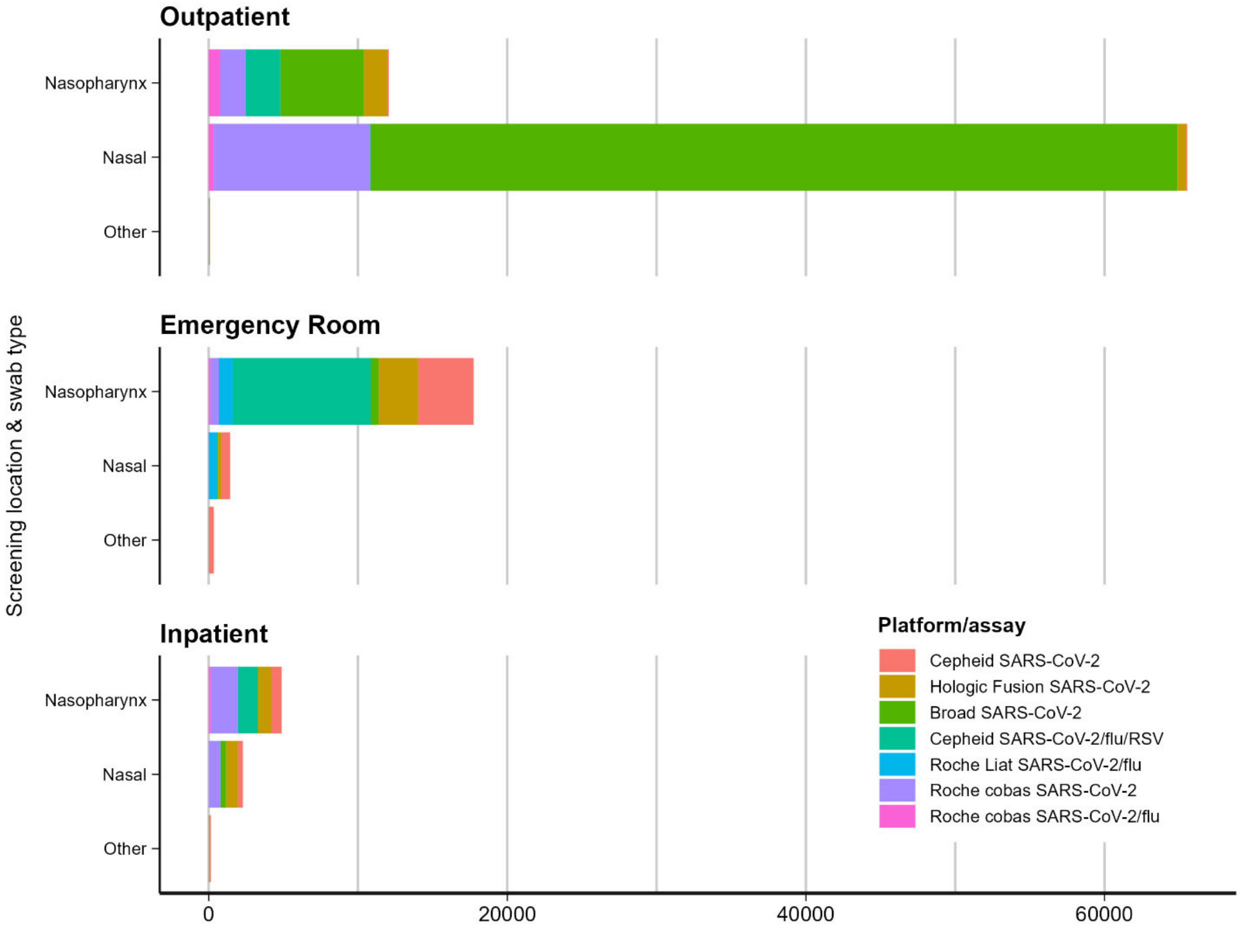


**Figure S24.** Distribution of test results included in the MGB dataset, broken down by screening location (outpatient pre-procedural screening, ER testing, inpatient testing), swab type (nasopharyngeal vs. nasal vs. other), and PCR platform / assay used for analysis.

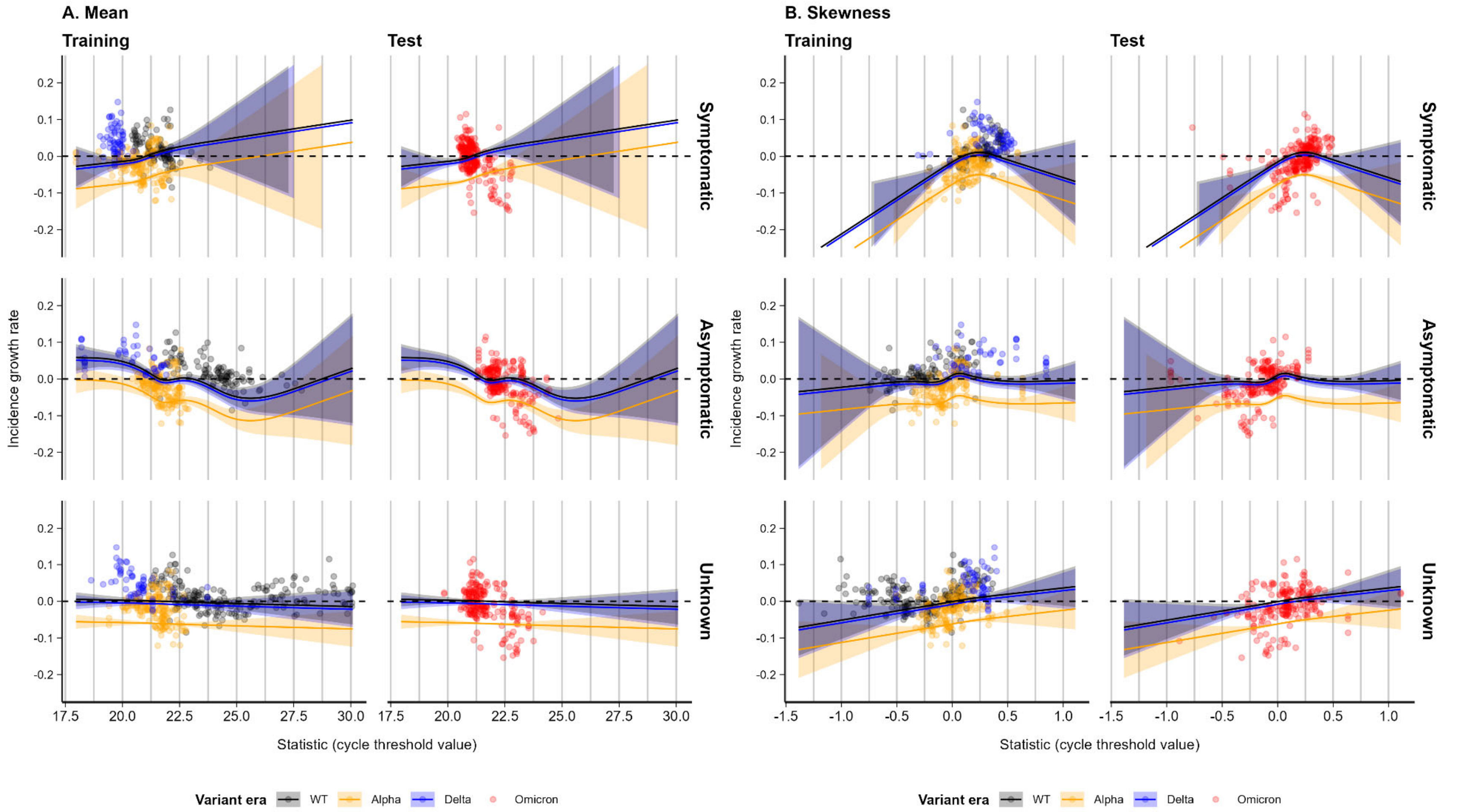


**Figure S25.** Best-fit lines (with standard errors) showing the modeled relationship between mean (left) and skewness (right) in Ct values against growth rate, stratified by reported symptom status, for LAC data, up to 31 Dec 2021 (training period) and after (test period). Note the differing relationships by symptom status stratum.

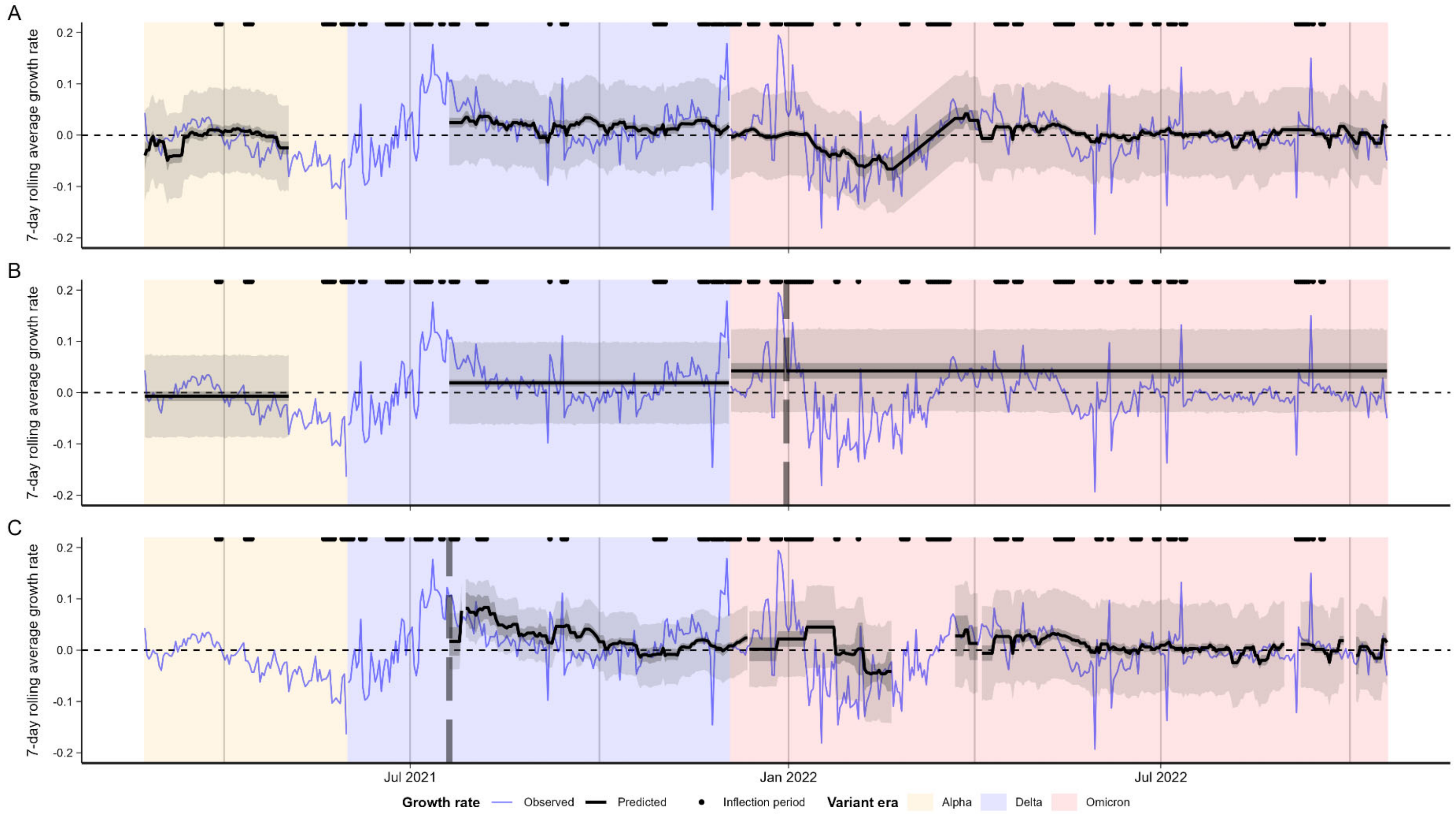


**Figure S26.** Model-predicted vs. observed log incidence growth rate for models fitted using the Tufts data, showing **(A)** in-sample fits, **(B)** fit over the testing period with a single fixed train-test split shown by the vertical dashed line, and **(C)** two-week rolling nowcast fits starting at the dashed vertical line.

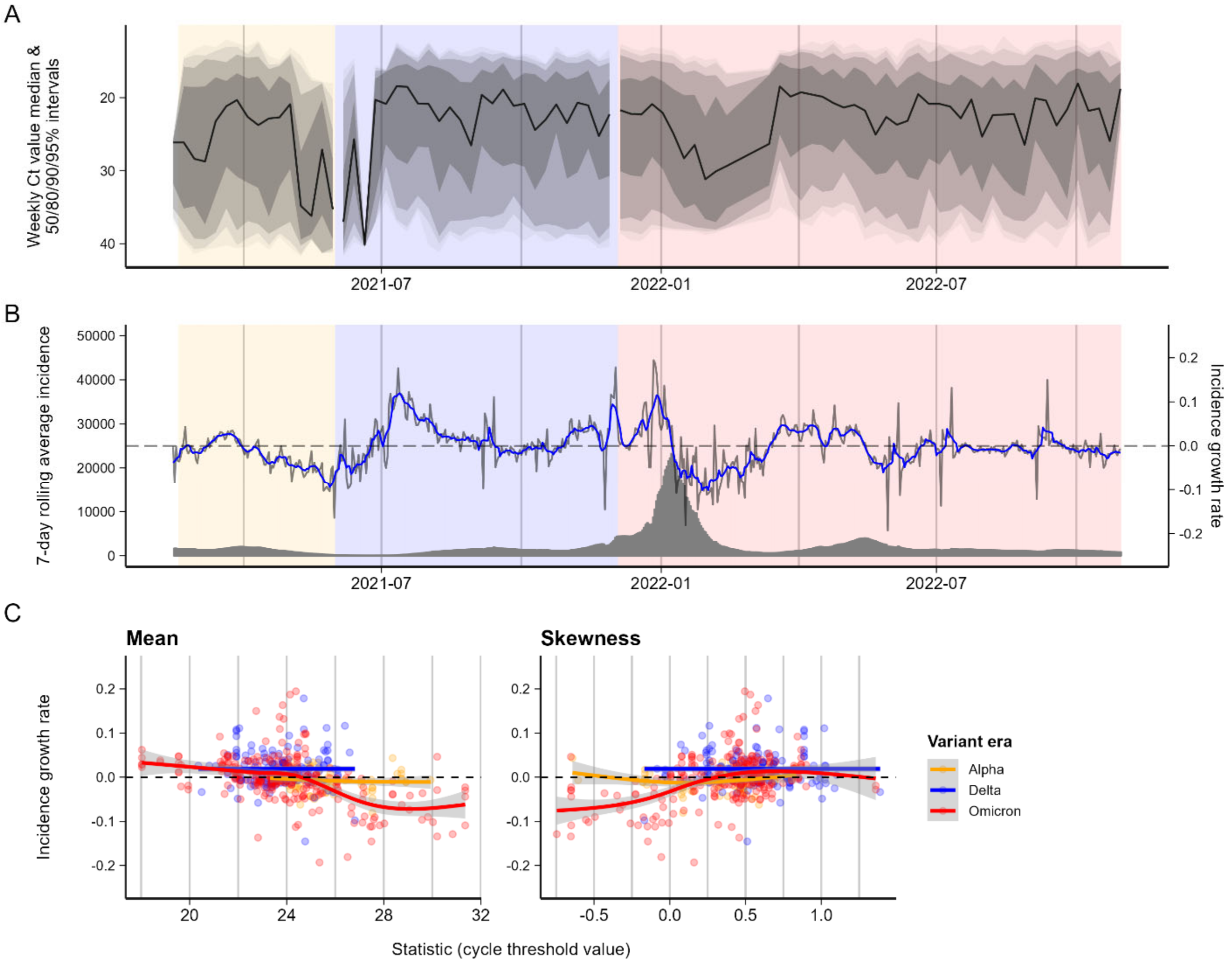


**Figure S27.** (**A**) Ct value distributions by week for the Tufts dataset. Solid line shows mean, shaded ribbons show 50% and 95% quantiles. (**B**) Incidence of COVID-19 cases and corresponding epidemic rates for Massachusetts, USA. Grey line shows growth rate of cases, whereas blue line shows 7-day rolling mean growth rate. (**C**) Relationship between Ct value means and skewness against epidemic growth rates.

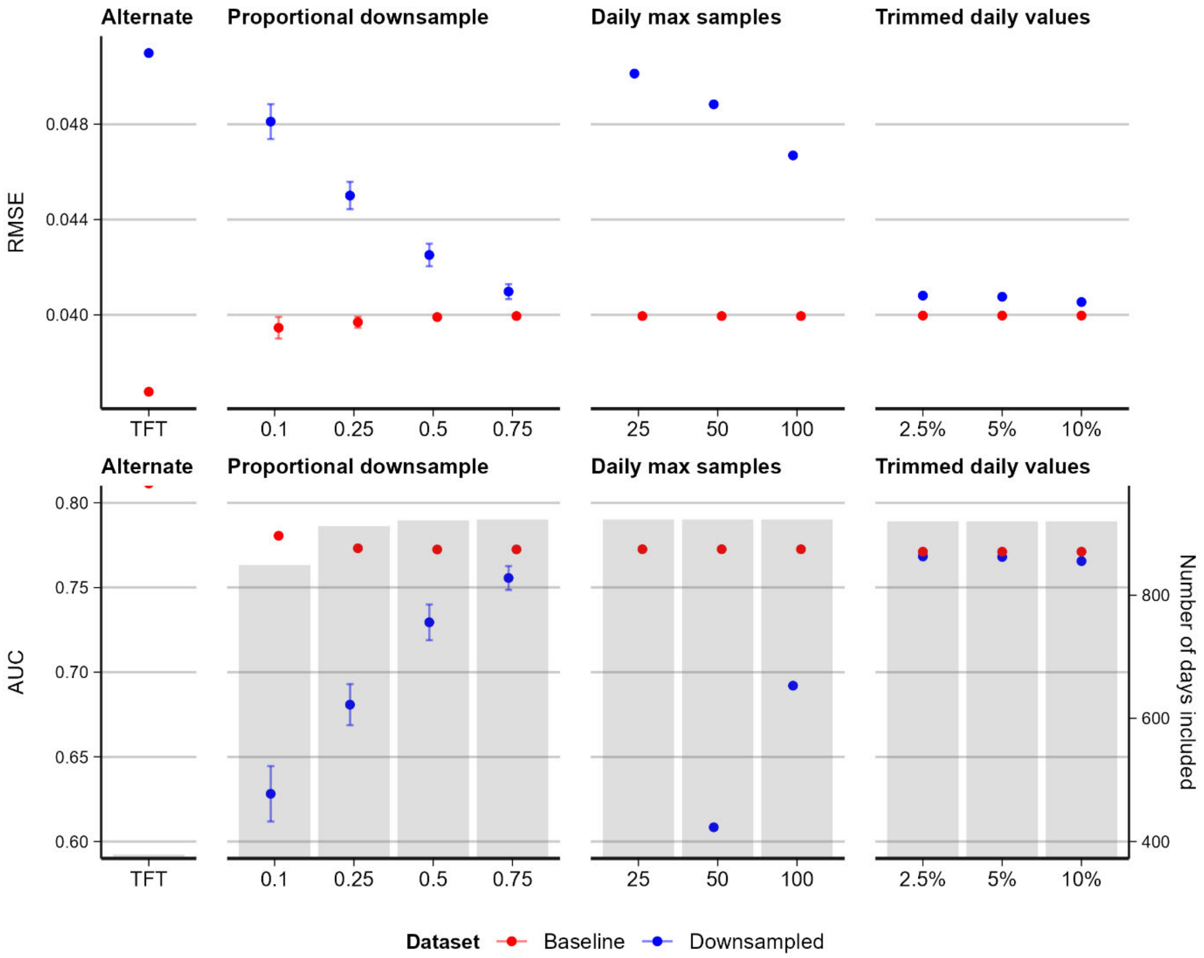


**Figure S28.** Model performance for downsampled MGB and full Tufts datasets, for models incorporating within-dataset positive test counts, but excluding Ct values. Baseline comparison metrics are re-calculated for only the days included in each downsampled dataset's nowcast. For proportional and daily max downsampling, both downsampled and baseline performance are averaged over 100 random draws (and their corresponding days included). Trim percentages indicate quantiles trimmed from each end of daily Ct value distributions (i.e., 5% trim yields the 5-95 percentile range of Ct values). Grey bars indicate the number of days of data included for the various downsampling analyses.

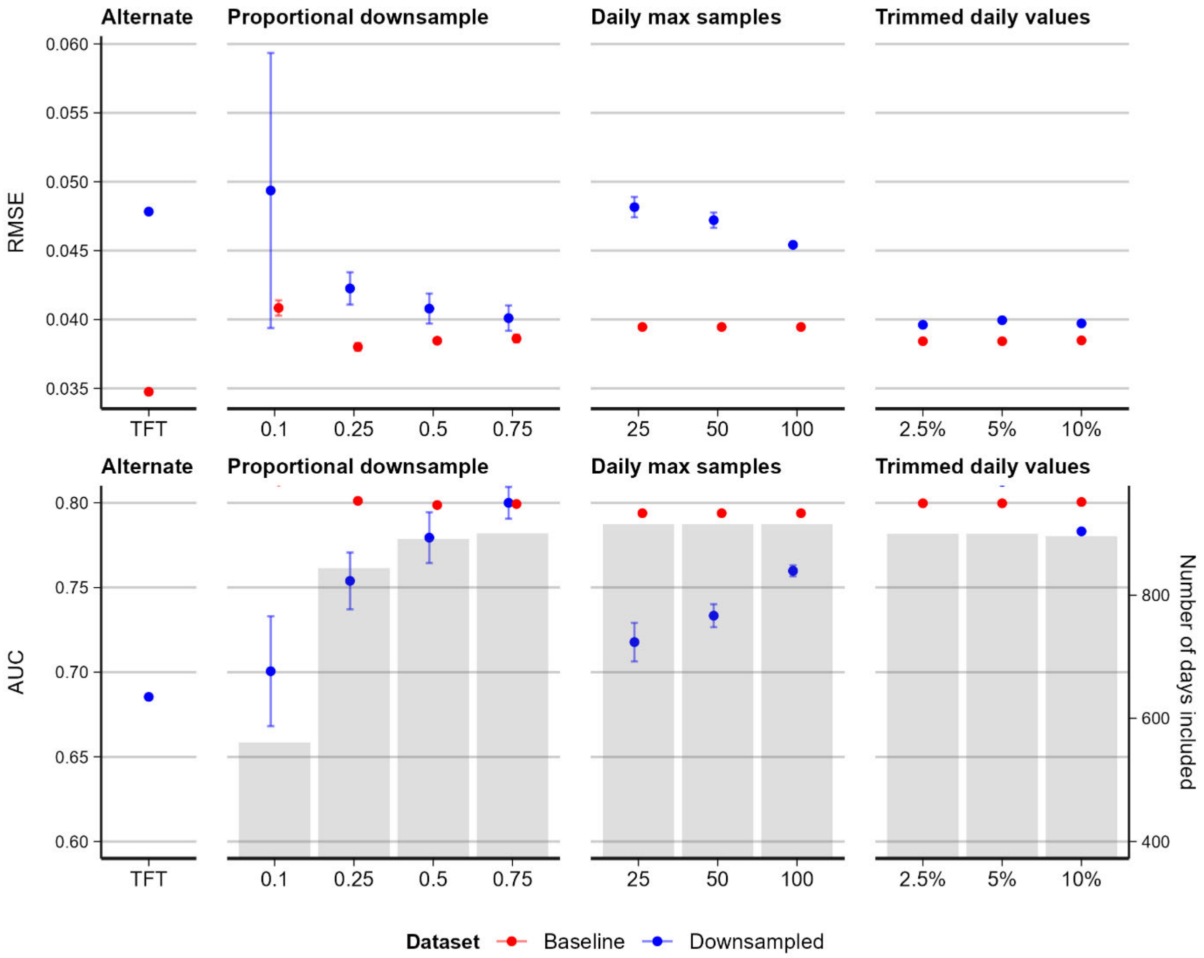


**Figure S29.** Model performance for downsampled MGB and full Tufts datasets, for models incorporating both within-dataset positive test counts and Ct value mean and skew. Baseline comparison metrics are re-calculated for only the days included in each downsampled dataset's nowcast. For proportional and daily max downsampling, both downsampled and baseline performance are averaged over 100 random draws (and their corresponding days included). Trim percentages indicate quantiles trimmed from each end of daily Ct value distributions (i.e., 5% trim yields the 5-95 percentile range of Ct values). Grey bars indicate the number of days of data included for the various downsampling analyses.

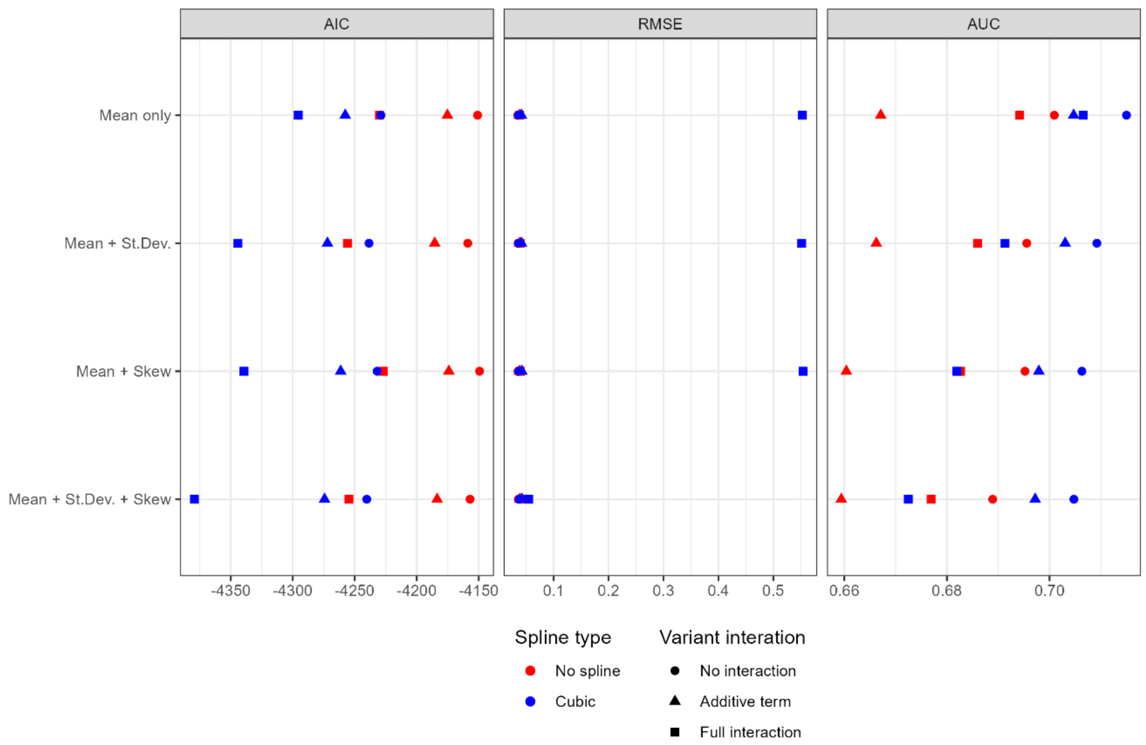


**Figure S30.** Comparison of predictive performance (RMSE and AUC) for the 24 tested models.

### Supplementary Text 1 – synthetic datasets

We built on a previously published model to simulate realistic Ct value distributions that would be expected under testing and sampling schemes similar to real-world data[6]. First, we parameterized a viral kinetics model describing the expectation and distribution of Ct values over all days following infection using previously published longitudinal SARS-CoV-2 testing data (**Figure S5, Table S7**)[2]. This is a piecewise linear model governed by a set of control points determining the time from infection to peak viral load, time from peak viral load to an inflection point at a high Ct value, and a longer-term clearance rate with a daily probability of full clearance. Second, we simulated approximately 2 million infections with infection times distributed based on the reported incidence of COVID-19 cases in Massachusetts between 5 March 2020 and 25 Feb 2023. Third, we simulated a surveillance system as a mixture of random testing (i.e., symptom-independent) and symptom-based testing (individuals are tested with a random delay following a randomly generated incubation period). Combining these three simulation steps gave a synthetic dataset of Ct values for a mixture of asymptomatic and symptomatic individuals tested at various times post infection and over a multi-wave SARS-CoV-2 epidemic (**Figure S1**). Different scenarios were captured by changing the parameters used either for the viral kinetics model or sampling delay distribution (**Figure S6**).

#### *1. Viral kinetics model*

We adapted a previously published viral kinetics model describing the mean and distribution of Ct values over time-since-infection[6]. Our simulations do not need to track individual infections, and thus unlike other published viral kinetics model[2,3,7,8], our model describes only the mean and variance of Ct values for all infections given time-since-infection rather than modeling each individual's viral trajectory. Model parameters and interpretation are shown in **Table S6**.

We used a piecewise linear model of the form:

$$f(t) = \begin{cases} c_0, & t \leq 0 \\ \mu t + c_0 , & 0 < t \leq t_p \\ c_p - \omega_1(t - t_p) + c_0 , & t_p < t \leq t_p + t_s \\ c_s - \omega_2\left(t - t_p - t_s\right) + c_0, & t > t_p + t_s \end{cases}$$

Where $c_0$ is the true baseline Ct value at time of infection; $\mu = \frac{c_p - c_0}{t_p}$ is the Ct value growth rate; $c_p$ is the minimum Ct value; $t_p$ is the time from infection to minimum Ct value; $\omega_1 = \frac{c_p - c_s}{t_s}$ is the initial clearance rate; $c_s$ is the Ct value at which waning switches to a second, slower clearance rate; $\omega_2 = \frac{c_p - c_0}{t_c}$ is the second, slower clearance rate; $t_c$ is the time taken to decay from $c_s$ to $c_0$.

We model the distribution of observed Ct values around the mean Ct value ($f(t)$) to capture three observations:

1. Ct values are highly varied on a given day post infection.
2. The variance of the Ct distribution is not necessarily constant over time.
3. Most individuals clear their infections quickly, but a small proportion remain detectable at a very high Ct value for many days after infection.

The distribution of observed Ct values on a given day post infection is modeled as a truncated Normal distribution:

$C(t) \sim N(f(t), \sigma(t))_0^{38}$

Where $N$ is the normal distribution. We assumed the distribution was truncated between 0 and 38 based on the distribution of observed Ct values in the NBA dataset. $f(t)$ is the mean of the Normal distribution and $\sigma(t)$ is the time-varying variance given by:

$$\sigma(t) = \begin{cases} \sigma_{obs}, & t < t_p + t_s \\ \sigma_{obs}(1 - \frac{1 - s_m}{t_m}(t - t_p - t_{s)}, & t_p + t < t \leq t_p + t_s + t_m \\ s_m \sigma_{obs}, & t > t_p + t_s + t_m \end{cases}$$

$\sigma_{obs}$ gives the variance. The second term describes a gradually decreasing variance during the second clearance phase, declining at a constant rate over duration $t_m$ before reaching a minimum of $s_m \sigma_{obs}$.

In addition, the probability of a sample having a detectable sample on a given day following infection is the product of two probabilities: the probability of having a Ct value less than the limit of detection, given by the cumulative density of the Normal distribution; and the probability of having not cleared the infection by that day:

$$\phi(t) = P[C(t) < c_0](1 - p_c)^{t - t_p + t_s}$$

The first part of the equation gives the cumulative density of the Normal distribution. The second part describes an additional process, whereby each day from $t_p + t_s$ onwards there is a daily probability, $p_c$, of becoming fully undetectable, representing clearance of the infection.

*2. Parameterizing the base model*

We parameterized the viral kinetics model using publicly available longitudinal data from the National Basketball Association (NBA) [2]. These data were a convenience sample from daily testing of NBA players, staff, and other affiliates over the course of the pandemic. Clinical samples were combined anterior nares and oropharynx swabs (collected separately from each anatomical site and combined in a single tube). Samples were tested using the Roche Cobas target 1 assay to give Ct values against the ORF1ab gene target. For this analysis, we used only tests from infections where the first positive sample was preceded by a negative test, intended to capture viral kinetics immediately following infection. Furthermore, as our objective was only to simulate a realistic model for the distribution of Ct values over time-since-infection, we did not stratify the data by covariates such as age group, symptom status, vaccination status or variant. Ultimately, we fit our model to 3,627 positive samples and 8,252 negative samples, representing 403 distinct infection episodes with samples taken between day 0 and 51 following a previous negative test.

We used a Markov chain Monte Carlo algorithm [9] to estimate posterior distributions for the model parameters conditional on the NBA dataset using uninformative uniform priors for the model parameters and a likelihood function based on the model described above. We ran 3 chains for 150,000 iterations, discarding the first 50,000 iterations as burn in. High effective sample sizes (>1000) and $\widehat{R}$ values <1.1 were obtained for all estimated parameters. We used the maximum *a posteriori* estimates as point estimates for the simulations. Model fits are shown in **Figure S5**.

*3. Simulated surveillance data*

We simulated Ct values observed under a realistic surveillance system using the following algorithm:

1. Infection times were simulated for *N=2,000,000* individuals (the cumulative incidence of cases in Massachusetts) by drawing infection times from the 7-day rolling mean reported incidence of cases from Massachusetts.
2. Two surveillance strategies were simulated giving each individual two possible sampling times:
    a. Random cross-sectional testing, representing detection of symptomatic infections. Uniformly distributed sampling dates were simulated for each infection. The time-since-infection was given as the difference between the sampling date and infection date (thus, many individuals were sampled before they became infected or long after they cleared their infection). All individuals are assigned one random sampling time.
    b. Symptom-based surveillance was simulated by assuming all infected individuals became symptomatic (note that it is not important to reflect the true symptomatic fraction for SARS-CoV-2, as we are only interested in generating a large number of simulated Ct values). Each symptomatic individual was assigned a randomly generated incubation period drawn from a log-normal distribution with mean = 1.621 and standard deviation = 0.41 on the log scale. For symptom-based surveillance, each symptomatic individual was additionally given a sampling delay drawn from a distribution. An individual’s sampling date was given as their infection date, plus their incubation period, plus their sampling delay.
3. Expected Ct value at their sampling time were calculated using the viral kinetics model described above combined with the MAP estimates from the model fitting.
4. Time to full clearance was simulated for each individual from a negative binomial distribution with success probability $p_c$.

5. Finally, observed Ct values were simulated from a normal distribution with mean given by the expected Ct value given time-since-infection and the time-dependent variance as described above. If the simulated Ct value is greater than the limit of detection or the individual had already fully cleared the infection, then the Ct value was set to 40.

*4. Synthetic data scenarios*

To understand how the relationship between observed Ct values and true growth rate of infection incidence varies across different scenarios, we implemented 6 different scenarios. We start with the "Ideal" scenario, which modifies the simulation parameters to provide an unrealistic scenario where we expect to see a consistent and clear relationship between surveillance Ct values and incidence growth rates. We then return each of these simulation parameters to realistic values one at a time to understand which factors confound our ability to infer growth rates from surveillance Ct values.

Scenarios:

1. Ideal conditions: assuming that viral kinetics are extremely left-skewed (very fast growth phase relative to clearance phase), that there is little variation in observed Ct values given time-since-infection, and that the delay from symptom onset to sampling time is uniformly distributed between 0 and 7 days.
2. Realistic scenario: using the viral kinetics parameter estimates and variance from fitting the model, and assuming that sampling delays are gamma-distributed with a mean of 4 days and variance of 8 days.
3. Realistic viral kinetics: using the viral kinetics parameter estimates from fitting the model, but with half the variation in observed Ct values given time-since-infection and uniformly distributed sampling delays.

4. Realistic variation: using an extremely left-skewed viral kinetics curve and uniformly distributed sampling delays, but with the variance of the Ct value distribution given time-since-infection based on the model estimates.
5. Realistic sampling: using an extremely left-skewed viral kinetics curve and reduced variance in observed Ct values, but assuming that sampling delays are gamma-distributed with a mean of 4 days and variance of 8 days.
6. Symmetric viral kinetics scenario: using the same parameter values as the ideal scenario, but increasing the time between infection and peak viral load to 7 days

**Figure S6** shows the assumed viral kinetics models, and incubation and sampling delay distributions used for each of the scenarios. **Figure S3** shows the assumed epidemic growth rate curve, and the resulting mean Ct values over time through the two surveillance systems.

We also simulated a scenario where variants with different viral kinetics parameters dominated different waves. This scenario was based on the realistic scenario and used the same model parameters and data, but assumed that three successive variants circulated:

**1.** Pre-Delta variant using the "realistic scenario" parameters in **Table S7.**
2. Delta variant assuming a higher peak viral load (peak Ct value of 23.0 vs. 26.1) and earlier peak (delay between infection and peak of 1.50 days vs. 2.41 days).
3. Omicron variant assuming a later peak (4.00 days vs. 2.41 days) and longer initial clearance phase (10.0 days vs. 5.20 days).

We used publicly available data on variant proportions for the Northeast USA from the CDC to assign the proportion of cases to each variant, assigning each lineage into its WHO variant of concern nomenclature and finally into Pre-Delta, Delta and Omicron variants [10].

*5. Analytical convolutions*

We used the *virosolver* R package to generate analytical convolutions between the infection incidence curve and the viral kinetics model to calculate the expected Ct value distribution for each day. The convolution for cross-sectional testing is defined as:

$$P(X = x|\pi_{t-50}, \dots, \pi_t|) = \sum_{a=t-50}^{t} p_a(x)\varphi_a \pi_{t-a}$$

For $x < Ct_{lod}$, and for $x = Ct_{lod}$

$$P(X = Ct_{lod}|\pi_{t-50}, \dots, \pi_t) = 1 - \sum_{a=t-50}^{t} \varphi_a \pi_{t-a}$$

Where $\pi_t$ gives the probability of infection at time *t*; $\varphi_t$ gives the probability of being detectable at *t* days post infection; and $p_a(x)$ gives the probability of observing a Ct value of *x* at *a* days post infection. Note that $x_a \sim N\big(f(a), \sigma(t)\big)_0^{38}$ as described above.

The convolution for symptom-driven testing is similar but implements additional convolutions with the incubation period distribution and distribution of delays between symptom onset and sampling:

$$P(X = x|\pi_{t-50}, \dots, \pi_t|) = \sum_{a=t-50}^{t} \sum_{o=0}^{50} \sum_{d=0}^{50} p_a(x)\gamma_o \beta_d \varphi_a \pi_{t-a}$$

Where $\gamma_o$ gives the probability of an incubation period of *o* days (given by a discretized log-normal distribution); and $\beta_d$ gives the probability of a delay between onset and sampling of *d* days (given by a discretized gamma distribution or uniform distribution). Note that in these convolutions an arbitrary upper bound of 50 days is set for all delays.

### Supplementary Text 2 – model choice

To determine our choice of model, we tested a series of regression models and GAMs, using different predictors (daily Ct mean, standard deviation, and skewness), functional forms (log-linear vs. cubic regression splines), and variant interaction terms. We fitted these models to the baseline synthetic dataset and compared their AIC as well as in-sample and nowcasting performance (see below). There was a clear bias-variance tradeoff between models; more flexible model specifications yielded better AIC and in-sample fit, at the cost of worse out-of-sample or nowcasting performance (see **Table S2** and **Figure S25**). We ultimately selected the final model using mean and skewness with a cubic spline, as the theoretical relationship between cross-sectional Ct values and epidemic growth rates is non-linear and depends on the *distribution* of Ct values observed; short of fitting the growth rate model to the entire distribution of observed values, using mean and skewness provides a parsimonious way to include information about the shape of the distribution in the model.

### Supplementary Text 3 – data sources

*Massachusetts*

Massachusetts Ct value data comes primarily from testing in 16 hospitals in the Mass General Brigham hospital system, with a catchment area largely in eastern Massachusetts. The full dataset comprises 2,671,041 SARS-CoV-2 test results, with specimen collection dates ranging from 3 Mar 2020 to 23 Feb 2023, of which 161,273 were positive. There were 3531 individuals who appeared to experience repeat infections (defined as >60 days between positive results), of which 72 individuals had 2 or more repeat infections. As we could not rule out long COVID or other idiosyncratic viral kinetics, we drop these 72 individuals from the final dataset. Limiting to results reporting Ct values and first reported Ct values for each confirmed case yields the final sample of 104,534 Ct values used in this analysis (**Table S3**), of which the earliest specimens were collected on 31 Mar 2020.

Samples are from a combination of routine outpatient (77,700; 74.3% of samples) and inpatient (7,311; 7.0%) screening and diagnostic tests, as well as ER patient testing (19,523; 18.7% of samples); while not entirely random nor representative, routine screening tests suffer less self-selection bias than symptom-based or voluntary testing. We did not have access to information on patients' vaccination or infection history, infecting variant, or symptom status.

The final sample includes specimens collected from nasal and nasopharyngeal swabs (approx. 2:1 ratio). Specimens were processed using seven different RT-qPCR platform/assay combinations (**Table S3**), variously targeting E/N/N1/N2/ORF1ab genes. For the main analysis here, Ct values were pooled across platforms/assays; where a single result reported Ct values for multiple target genes, the lowest value was used.

Daily confirmed case counts for Massachusetts were obtained from the Massachusetts Department of Public Health COVID-19 dashboard[11].

We also analyzed a secondary dataset of Ct values from Tufts Medical Center in Boston, Massachusetts for comparison. This dataset comprised 84,848 test results with collection dates ranging from 18 Feb 2021 to 31 Oct 2022, of which 10,338 were positive. Filtering the reported test results using the same criteria as used for the MGB data yielded a final sample of 10,214 Ct values used here. **Figure S27** summarizes the reported Ct value distributions over time and compares these to reported COVID-19 incidence.

*Los Angeles County*

LAC Ct value data comes from municipal COVID-19 testing sites operated by the LAC Department of Public Health and Department of Health Services, comprising approximately 10% of all municipal testing conducted in LAC during the sample period. The full dataset comprises 330,034 SARS-CoV-2 positive test results, with specimens collected over two time periods – 21 May 2020 to 27 Jul 2021, and 30 Dec 2021 to 29 Sep 2022. (Note: data were unavailable for the intervening period.) The data contain an infection episode identifier; limiting to the first reported Ct value for each infection episode yields the final sample of 279,492 Ct values used in this analysis.

The final sample includes specimens collected through nasal, nasopharyngeal, and oral swabs, and analyzed by Fulgent Genetics using an in-house platform and ThermoFisher QuantStudio™ 6 and 7 PCR systems. Two RT-qPCR assays were used; before mid-Nov 2020, analyses used exclusively LOINC 94531-1 targeting N1 and N2 genes, while subsequently the majority of analyses used LOINC 94533-7 targeting the N gene. Where a single result reported Ct values for multiple target genes, the lowest value was used. Symptom status was reported for approximately 75% of the sample, of which in turn approximately 75% (56% of the full sample) are reported as symptomatic for COVID-19 (**Table S3**). For symptomatic cases, most specimens were collected 1-10 days after symptom onset (modal delay of 3 days). The sample also included vaccination status, with approximately 24% of results coming from vaccinated (partially, fully, or boosted) individuals (**Table S3**).

Daily confirmed case counts were obtained from the LAC DPH COVID-19 dashboard[12].

### Supplementary Text 4 – sample size & outlier removal sensitivity analyses

As a sensitivity analysis, we repeated the rolling nowcast analyses using versions of the MGB dataset downsampled in two ways: 1) by randomly drawing 10/25/50/75% of the total test results available, or 2) by limiting the maximum number of positive test results for each day to 25/50/100, discarding any additional tests. We then reassessed nowcasting performance on each of these downsampled datasets. We repeated this analysis with 100 random downsampling draws for each size, taking the mean of model performance metrics over the 100 draws. We also compared nowcasting performance with a similar analysis using a third, smaller dataset from Tufts Medical Center, which uses the same response variable data as the MGB dataset (i.e., log incidence growth rates for Massachusetts) but has approximately 10% of the total sample size.

The downsampling process can result in some days being excluded from the downsampled dataset model's nowcast. Nowcasting performance can vary considerably from day to day, with outlier days having disproportionate impact. To ensure fair comparison of the impact of downsampling on model accuracy, rather than the impact of certain days being excluded as an indirect result of the downsampling process, we recalculated performance metrics for the baseline model's nowcasts based on just the days included in any given downsampled model's nowcasts, again taking the mean of model performance metrics over the 100 different baseline subsets included for each sample size.

To assess the impact of outliers on nowcasting performance, we trimmed daily observed Ct value distributions by 2.5/5/10% (yielding 95/90/80% ranges) before calculating Ct value distribution statistics, using the trimmed data for both training and nowcasting. Repeat draws were not required as the trimming is deterministic. As with the downsampling analysis, we recalculated baseline model performance metrics for only days included at each trim level.

We also repeated these analyses using the models incorporating only within-dataset case count growth rate, as well as the combined models incorporating within-dataset case count growth rate along with Ct value mean and skewness, as described in the previous section.
